# Risk factors for falls in community-dwelling older adults: An umbrella review

**DOI:** 10.1101/2025.04.04.25325029

**Authors:** Stephanie Saunders, Cassandra D’Amore, Quikui Hao, Nabil Abdelmoneim, Julie Richardson, Ayse Kuspinar, Marla Beauchamp

**Affiliations:** School of Rehabilitation Sciences, Faculty of Health Sciences, McMaster University

**Author notes:** **Corresponding Author:** Marla Beauchamp School of Rehabilitation Science McMaster University Institute of Applied Health Sciences, Room 403 1400 Main Street West Hamilton, ON, Canada L8S 1C7.

## Abstract

**Background:** Falls are a key public health concern, resulting in disability and increased mortality risk. An extensive body of literature has examined risk factors for falls, however results vary across different studies and populations. We aimed to synthesize systematic reviews on fall risk factors in community-dwelling older adults.

**Methods:** A systematic review of systematic reviews. Searches were executed in six databases (Medline, Embase, CINAHL, Cochrane Library, PsychINFO, Ageline) from inception until June 13, 2023. Eligible studies included systematic reviews of prospective cohort studies that included a population of community dwelling older adults (≥60 years) and reported fall risk factors. Three reviewers (SS, CD, QH) screened 8173 records. Summary data were extracted and the units of analyses were the relationships between risk factors and falls. Descriptive results are reported in counts and frequencies. The study was registered in Prospero (CRD42022335392).

**Findings:** Fifty-seven reviews were included. Mobility-related measures (balance, gait, physical function, physical activity, dual task ability, strength, range of motion) accounted for 40% of all relationships. Of these, clinical tests of balance and physical function were consistently predictive of falls. Other consistent predictors were cognition, specifically executive function (76% significant) and processing speed (100%), medications (58%), frailty (100%), and chronic conditions (83%). There was a paucity of evidence for psychosocial, environmental, and sociodemographic factors. The majority of reviews (54%) were rated as low risk of bias.

**Conclusions:** Mobility-related risk factors for falls are well established and can be addressed through interventions. Findings highlight the limited examination of psychosocial, sociodemographic, and environmental risk factors for falls, indicating areas for future research.

**Highlights:**

- This study synthesized all systematic and scoping reviews from six databases that examined 29 fall risk factors in community-dwelling older adults (≥60 years) from over 300 prospective studies
- The results highlight the extensive evaluation of mobility-related measures and identifies promising factors for fall risk assessment (i.e., clinical tests of balance and physical function, executive function, processing speed, frailty, chronic conditions)
- The study synthesizes the magnitude of relationships between risk factors with meta-analysis results
- The evidence from this study will help researchers refine those risk factors most important for identifying fall risk. Psychosocial, sociodemographic, and environmental factors could be valuable targets for future research on fall prevention.

## Introduction

Falls in older adults are a critical and growing global health problem.^1^ Approximately 27% of older adults fall each year around the world, with many falls resulting in serious disability and death. By 2050, one in four individuals in North America and Europe will be over the age of 65,^2^ which means the incidence of falls and their burden will only continue to rise.^3^

Despite their devastating impact, falls can be prevented by as much as 25-40% in community-dwelling older adults when targeted fall prevention interventions are provided to those at risk.^3^ However, providing these interventions at scale and identifying individuals with the highest risk remains difficult. Contributing factors are often complex and multifactorial,^7^ and the literature on fall risk factors, while vast, is often inconsistent with varying results depending on study design, context, population, and the type of fall outcome examined. In particular, much of the literature on fall risk in community-dwelling older adults is combined with data from institutionalized older adults.^4-6^ Besides the change in environmental context, institutionalization can also significantly affect physical function and mobility,^7^ and there is a need to clarify risk factors specifically relevant to community-dwelling older adults.

The aim of this umbrella review was to synthesize the evidence on fall risk factors in community-dwelling older adults from systematic reviews of prospective studies. These data are urgently needed to inform effective fall risk assessment practices in the community given the scarcity of resources for fall prevention interventions.

## Methods

### Search Strategy and Selection Criteria

This umbrella review followed the methodology reported by Aromataris et al., ^8^ and Cochrane overview of reviews guidelines^9^ and the protocol was registered on Prospero (CRD42022335392). We conducted searches in six databases: Cochrane Database for Systematic Reviews, CINAHL, Medline (Ovid), Embase (Ovid), Ageline, Web of Science on June 13, 2023. The search strategy was co-constructed with a librarian with three themes: older adults, falls, and systematic reviews, with no restrictions on dates, publication types, or language. Complete search strategies for all databases are available in the supplementary material (Appendix 1). References of included articles and clinical practice guidelines were screened for eligible studies.

Reviews were eligible if they were systematic or scoping in nature, were published in English or Chinese, had an explicit aim to examine fall risk, and synthesized results where primary papers: 1) had a prospective study design, 2) included participants with a mean age of >60 years (>80% of primary papers), and 3) included participants who were community-dwelling (≥80% of primary papers). To be as inclusive as possible, we included reviews where there was any subset of synthesized results that met the criteria. Conference abstracts, grey literature, and narrative reviews were excluded. Titles and abstracts of search results were uploaded into Covidence software for deduplication and screening.

### Data extraction and quality assessment

Reviewers screened all abstracts and full text articles (SS and CD or QH), with discrepancies addressed through discussion. Data were abstracted by multiple reviewers (SS and CD) into an Excel table developed a priori. In the event the data was not available, SS emailed review authors. Two reviewers (SS and NA) assessed the risk of bias of each review using the *ROBIS* tool.^10^ The tool assesses four bias domains: study eligibility, identification and selection of studies, data collection and study appraisal, and synthesis and findings, and provides an overall summary rating. We report our results identifying the domains at greatest risk of bias. As recommended by Aromataris et al., ^8^, we did not exclude reviews based on quality.

### Data analysis

Results were synthesized at two levels. At the level of the review, we summarized review methods (e.g., years conducted, review aims, populations examined, setting definitions, fall definitions) and the characteristics of included primary papers (e.g., geographic locations, years conducted). At the level of the primary papers, we summarized information about the relationships between risk factors and falls. Relationships have been summarized in one of two ways based on the presentation of results in reviews. When meta-analyses results were available, we reported estimates and 95% confidence intervals. Due to the heterogeneity in statistical methods (odds ratios (OR), hazard ratios (HR), mean differences (MD), standard mean differences (SMD)) we did not synthesize results further for the magnitude of relationships. When meta-analyses results were not available, we recorded the frequency of significant results for each relationship (i.e., fall type and risk factor). Results have been organized according to the risk factors assessed and summarized in tables. Finally, we counted how many times each relationship was reported, either as an individual or meta-analysis relationship, and noted the percentage of relationships that were significant. This has been compiled into a table that includes all identified risk factors providing a comprehensive overview of the literature. For clarity, the included systematic and scoping reviews are referred to as reviews and individual studies within these reviews are referred to as primary studies.

### Corrected covered area (CCA)

As a function of including reviews on the same topic, there is the chance that the same articles appear multiple times. This may result in a bias that more heavily weights the multi-cited primary studies.^11^ The Corrected Covered Area (CCA), calculated as: (# of primary papers and how often they occur – # of unique primary papers) / (# of unique primary papers – # of reviews), assesses the extent of overlap between multiple systematic reviews^11^ providing a score that ranges from 0-100%. Classifications for overlap are slight= 0-5%, moderate =6-10%, high = 11-15%, or very high = >15%. The CCA measure was determined for the overall umbrella review as well as for the five most frequently identified predictors of fall risk. This score provides context for the reporting of each risk factor, highlighting where primary research overlaps.

## Results

A total of 8173 records were examined, see PRISMA diagram for full details (Figure 1). After de-duplication a total of 4105 abstracts and 477 full texts were screened for eligibility. Fifty-seven reviews were included, 53 reviews examined any fall outcome (i.e., did not differentiate between single falls, recurrent falls, or injurious falls) and 4 reviews examined only recurrent falls. All excluded full-text articles and reasons for exclusion can be found in Appendix 1, Table S1.1.

**Figure 1.**
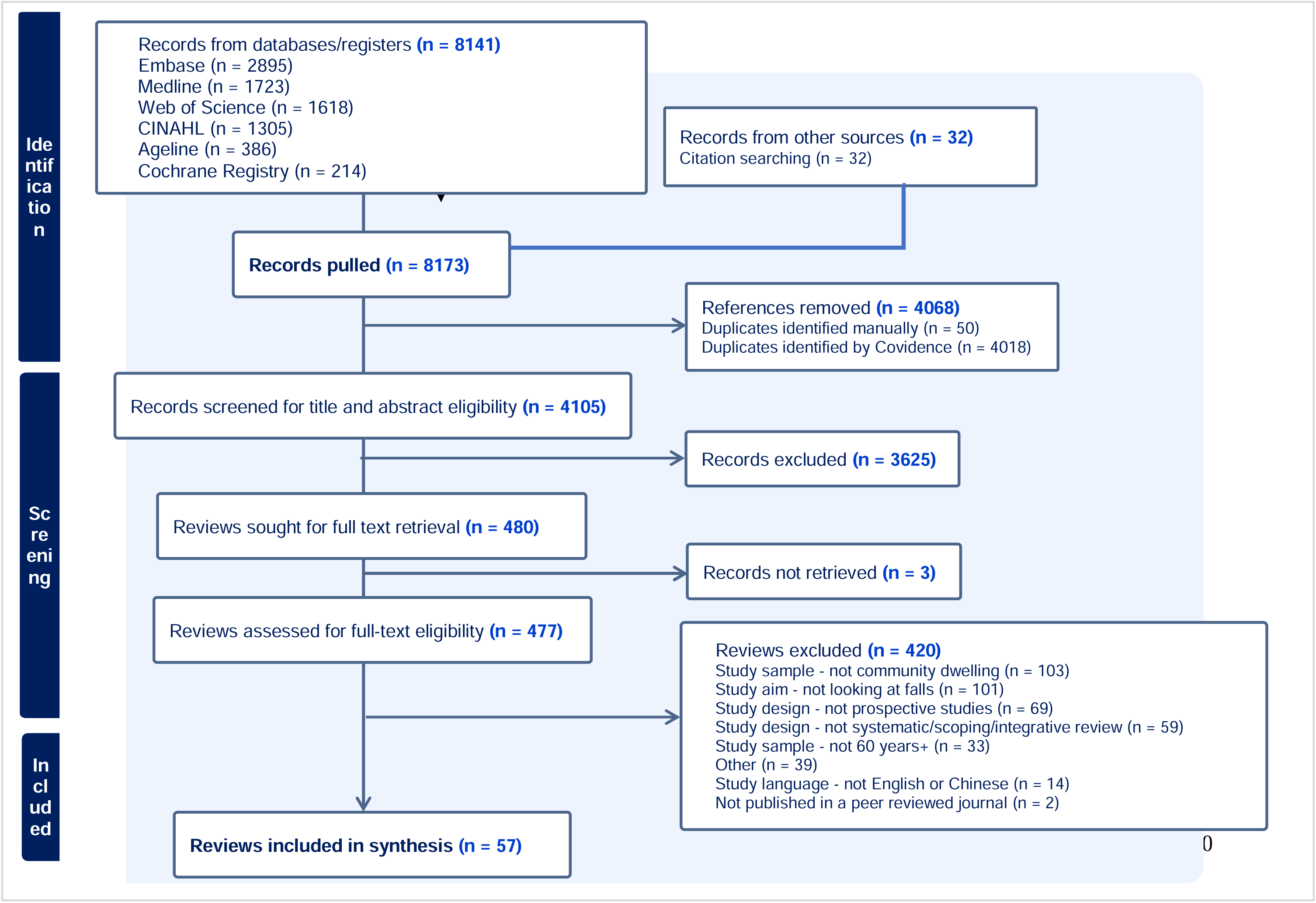
PRISMA Flow diagram.

The included reviews were conducted between 2004 and 2023, with primary studies conducted between 1981-2022. In total, 963 relationships were synthesized across 29 different risk factors; 660 relationships predicting any falls, 239 predicting recurrent falls, and 64 predicting injurious falls (Table 1). Thirty-eight reviews included studies that mostly took place in high income countries (67%), with only 6 reviews including studies that took place in lower middle-income countries and no reviews including studies from low-income countries. Sixteen reviews reported results that included male only samples, 34 reviews reported results that were female only and seven reviews reported only mixed samples. Clinical populations were explicitly examined in 7 reviews; three in dementia/cognitive impairment, one in patients with diabetes, one in patients with stroke, one in patients with depression, and one in patients with pain. Of the reviews that reported mean ages (n=52), they ranged from 68 to 84 years. Primary studies were synthesized both via meta-analyses (n=196 relationships; see Figure 2 for overview), qualitative narrative synthesis (n=660), and predictive validity testing (n=107). Most reviews reported that primary studies did adjust for confounders in their analyses, however few reported what these confounders were (n=518 did not report the nature of confounders). Of those reported, age, gender, and education were most common.

**Figure 2.**
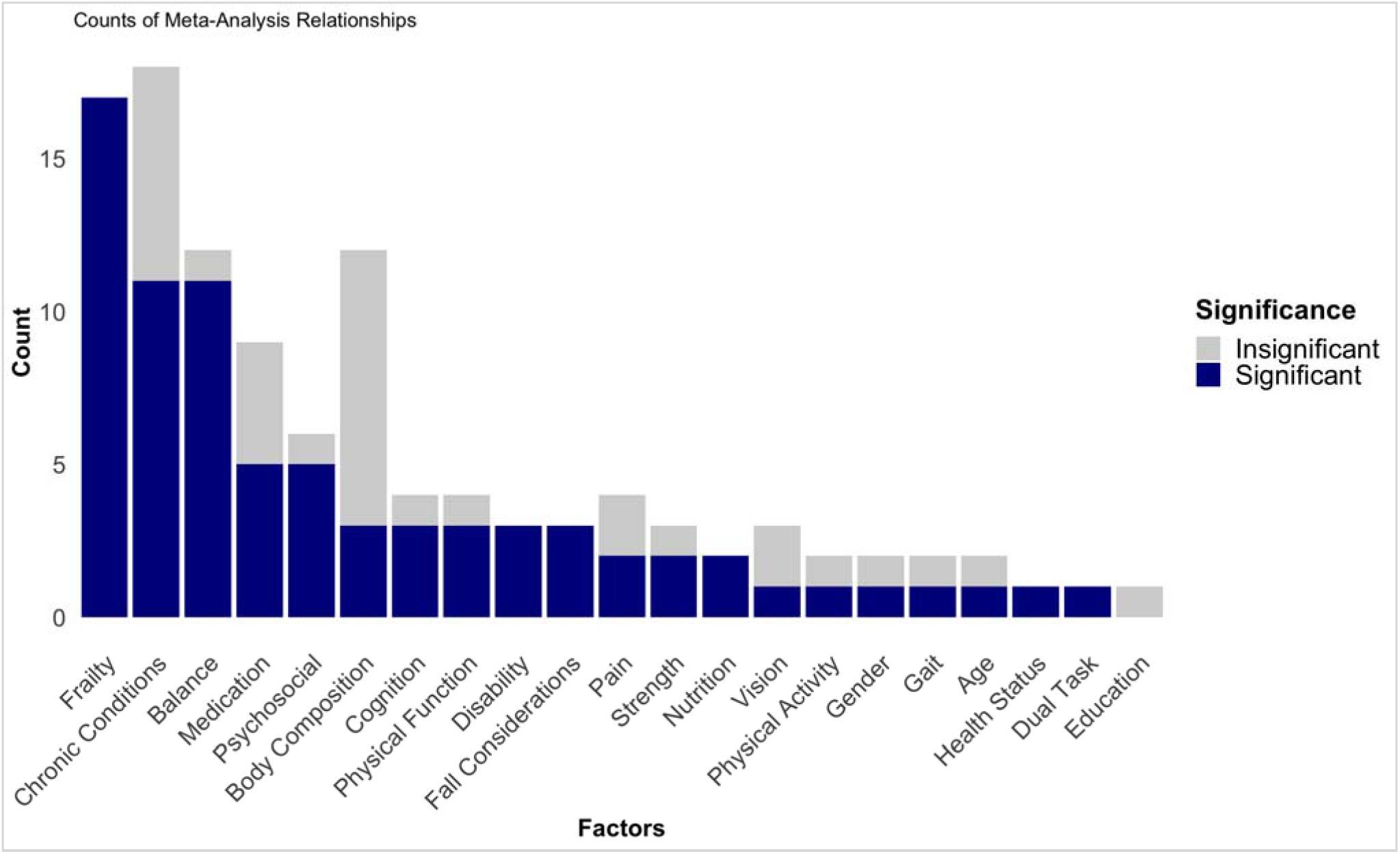
Counts of Meta-Analysis Relationships Predicting Any Falls. **Notes.** Counts of each meta-analysis relationship. Dark blue indicates a significant relationship, grey indicates an insignificant relationship, with the figure demonstrating the proportion of each predictor that is significant.

**Table 1.** Characteristics of Included Reviews.

| **Review characteristics** |  |
| --- | --- |
| **Years reviews were published** | 2004-2023 |
| Years primary studies were published | 1981-2022 |
| **Clinical populations examined by review (n=reviews)** |  |
| Dementia/cognitive impairment, | N=3 |
| Diabetes | N=1 |
| Stroke | N=1 |
| Depression | N=1 |
| Knee Osteoarthritis | N=1 |
| **Countries where primary studies were published (n=reviews)\*** |  |
| High income | N=38 |
| Upper mid income | N=21 |
| Lower mid income | N=6 |
| Lower income | N=0 |
| **Gender (n=reviews)** |  |
| Reviews including primary studies examining only men | N=16 |
| Reviews including primary studies examining only women | N=34 |
| **Relationships examining any predictors of any fall (i.e., $\geq 1$)** | N=660 |
| Predictive validity testing | N=82 |
| Meta-analyses | N=112 |
| Narrative single relationships | N=466 |
| **Relationships examining any predictors of recurrent falls (i.e., $\geq 2$)** | N=239 |
| Predictive validity testing | N=25 |
| Meta-analyses | N=74 |
| Narrative single relationships | N=140 |
| **Relationships examining any predictors of injurious falls** | N=64 |
| Predictive validity testing | N=0 |
| Meta-analyses | N=10 |
| Narrative single relationships | N=54 |
| **Total number of participants across all reviews** | N=9,582,108<sup>†</sup> |
| **Mean age ranges across all reviews, years** | 68-84<sup>‡</sup> |
| **Total number of primary papers included** | N=356 |
| Notes:<br>* 17 reviews did not report countries of primary studies<br><sup>†</sup> 34 reviews did not report a total review-wide sample size; some participants were not included in the final reported results; some participants may have been included more than once due to overlap in primary studies<br><sup>‡</sup> 5 reviews did not report a review-wide mean age |  |

Across reviews, falls were categorized in various ways: any falls (n=53), recurrent falls (n=48), injurious falls (n=19), falls that took place outdoors (n=1), and falls that took place indoors (n=1). Thirty-three reviews (58%) published between 2004-2023 did not report a definition of falls. Of those that did, the most common definitions were from Lamb et al., ^12^(n=5) and Gibson et al., ^13^(n=3). With the exception of one review, all falls were self-reported, and the most common means of recording falls were via fall calendars or diaries (reported daily, weekly, bi-monthly, monthly) and phone call interviews (reported weekly, monthly). Follow-up time periods ranged from 1 month to 252 months, with an average of 18 months.

The most commonly examined fall risk factor was mobility (n=307 relationships), consisting of measures of balance (n=142), gait (n=57), physical function (n=38), physical activity (n=32), dual-task ability (n=19), strength (n=15), and range of motion (n=4). The other most commonly examined risk factors included cognition (n=80), chronic conditions (n=42), medications (n=26), and frailty (n=17). Of these, only half of them reported ≥60% of their relationships as significant (gait, chronic conditions, physical function, dual task ability, frailty; Table S1.3). Other predictors where: 1) more than 5 relationships were examined, and 2) ≥60% were significant included: fall-specific factors (i.e., fall history, falls self-efficacy), gender, oral health, psychosocial factors, pain, disability, and sensorimotor factors (Table S1.3). Fewer relationships were reported for sociodemographic predictors such as age or education. Of these, older age as a risk factor for falls was inconclusive, with ORs from meta-analyses ranging from 1.02-1.12, and 50% of them reaching significance (Table 2, Table S1·4). Similarly, of the 6 narrative relationships examining age as a risk factor, only 3 were significant (Table 3, Table S1·5). Female gender also had only 50% of studies demonstrating an increased risk, with ORs from meta-analyses ranging from 1·01-1·30. Only one meta-analysis examined education as a predictor for falls and it was insignificant (OR=1·01). For details see Table 2. Results for recurrent falls and injurious falls were similar, with a greater number of significant relationships and a greater range of psychosocial outcomes (e.g., social connectedness, depression, psychological wellbeing), and fall-specific factors, such as a fall self-efficacy (Appendix 2 and 3).

**Table 2.**
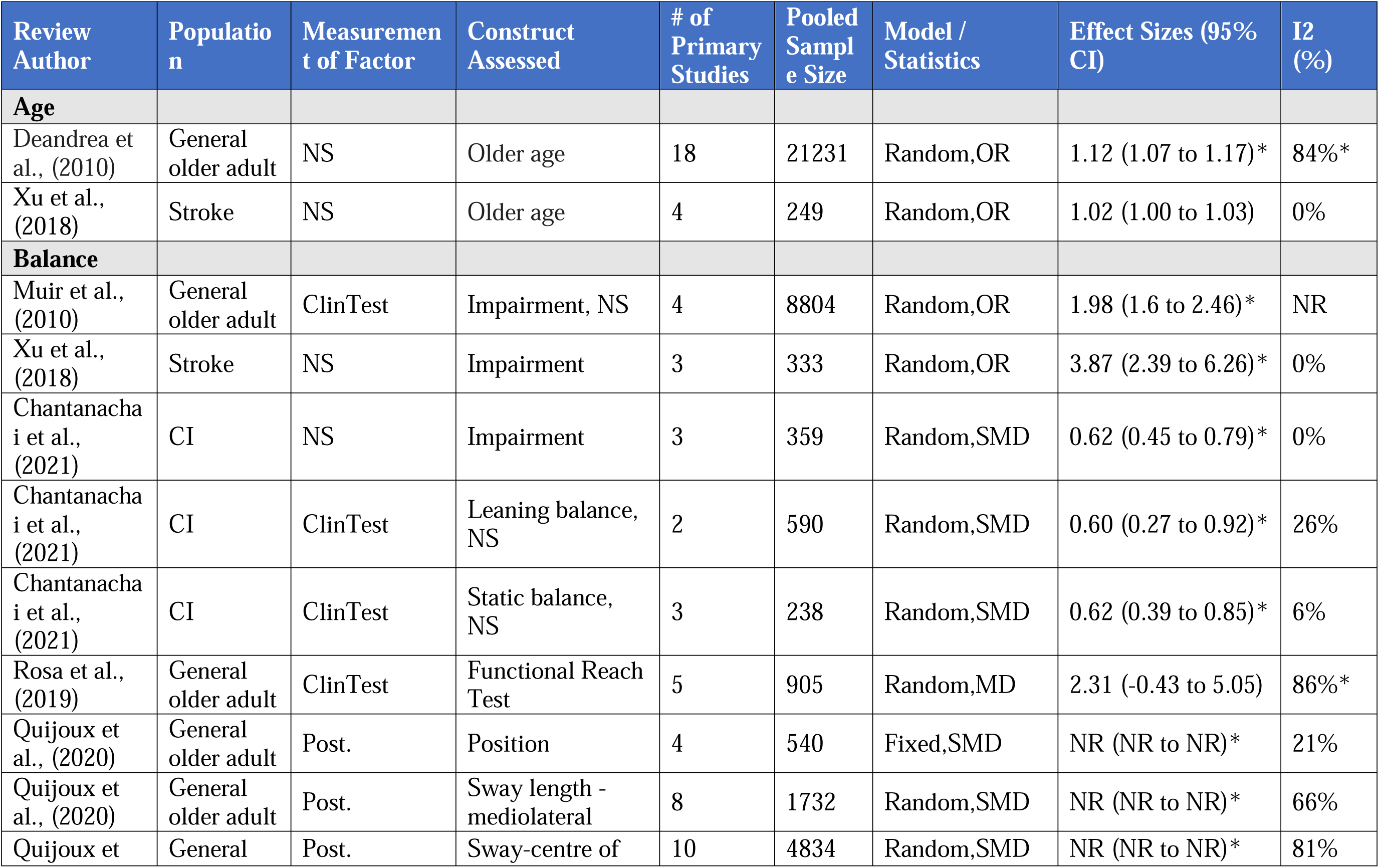
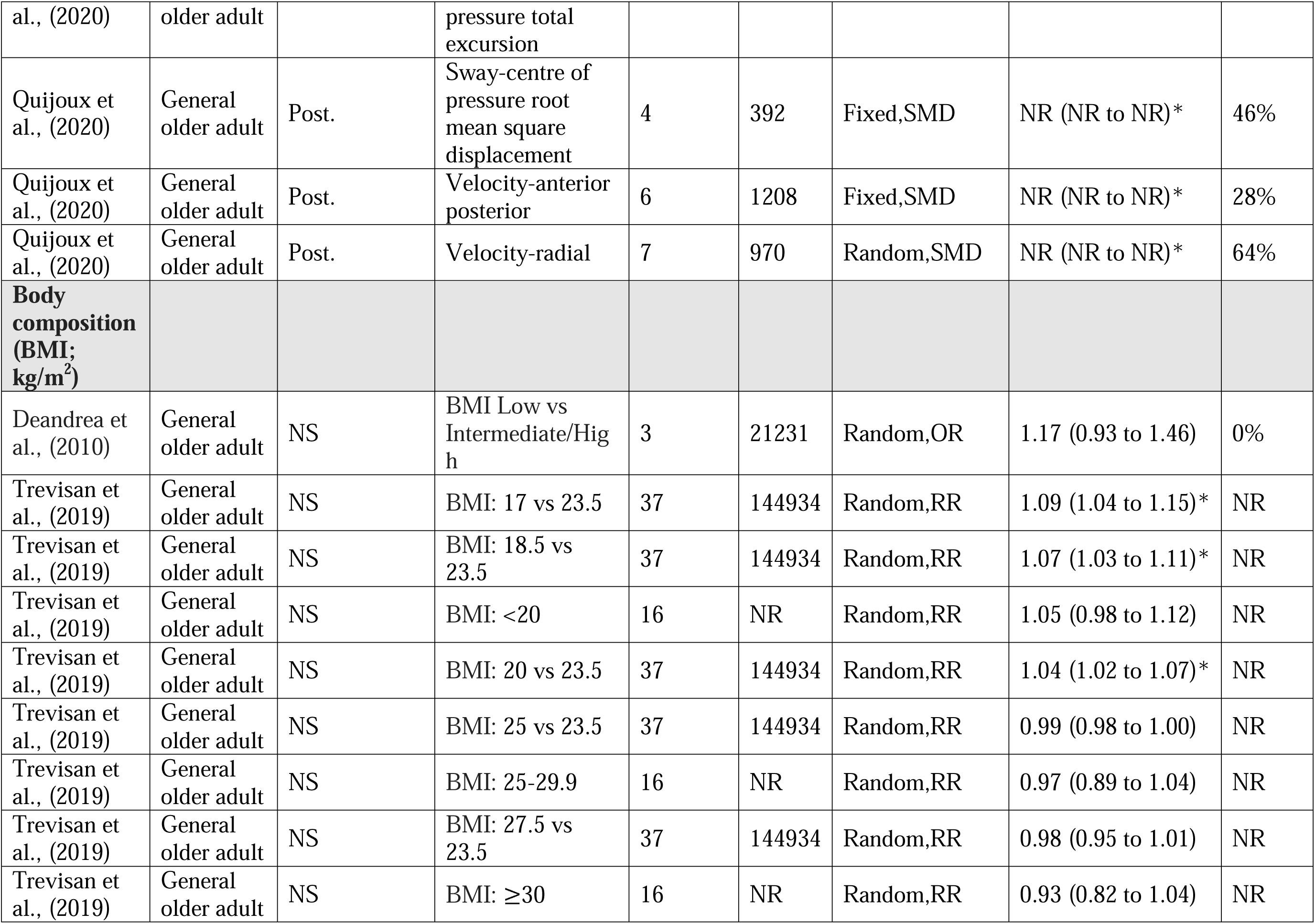
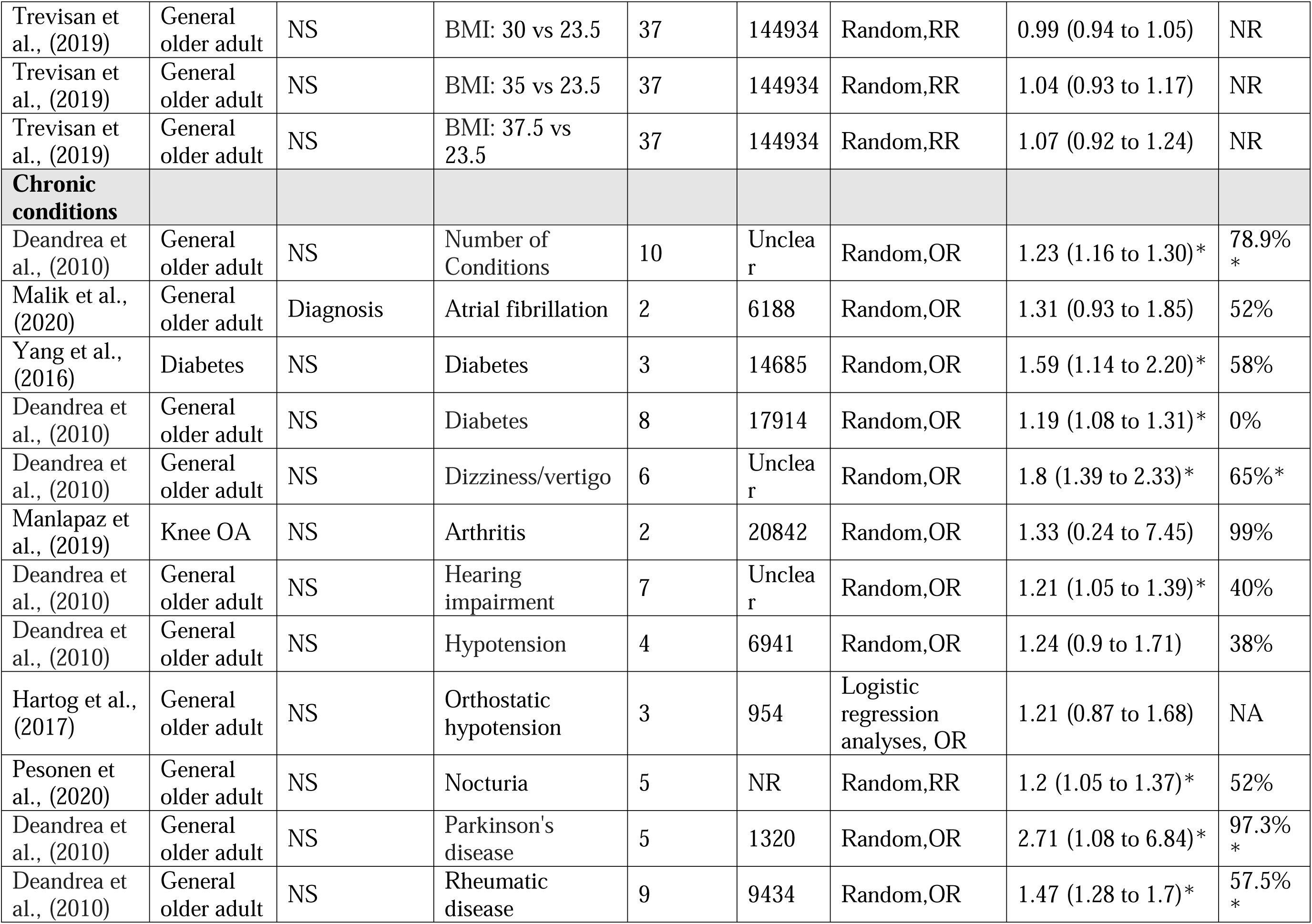
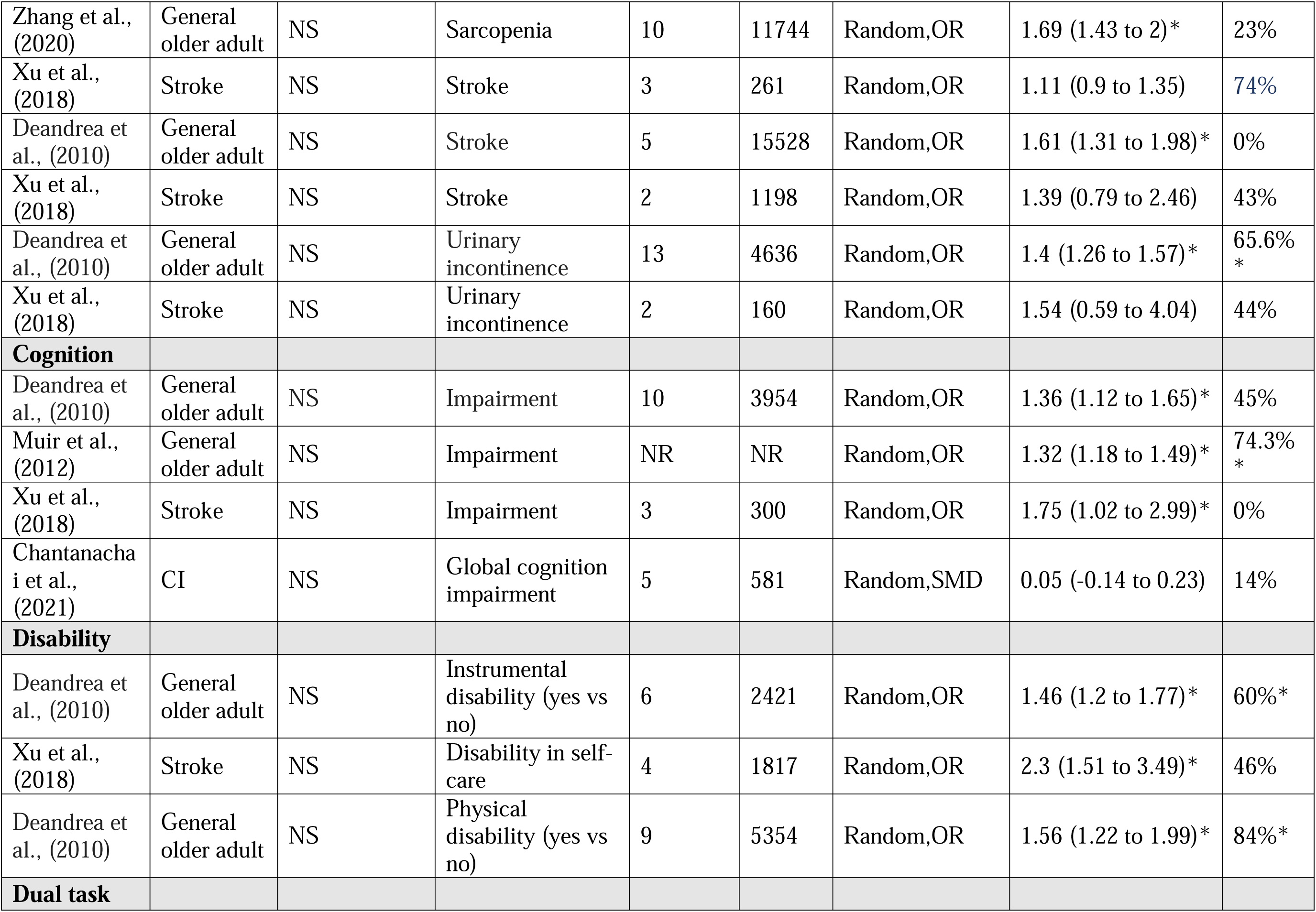
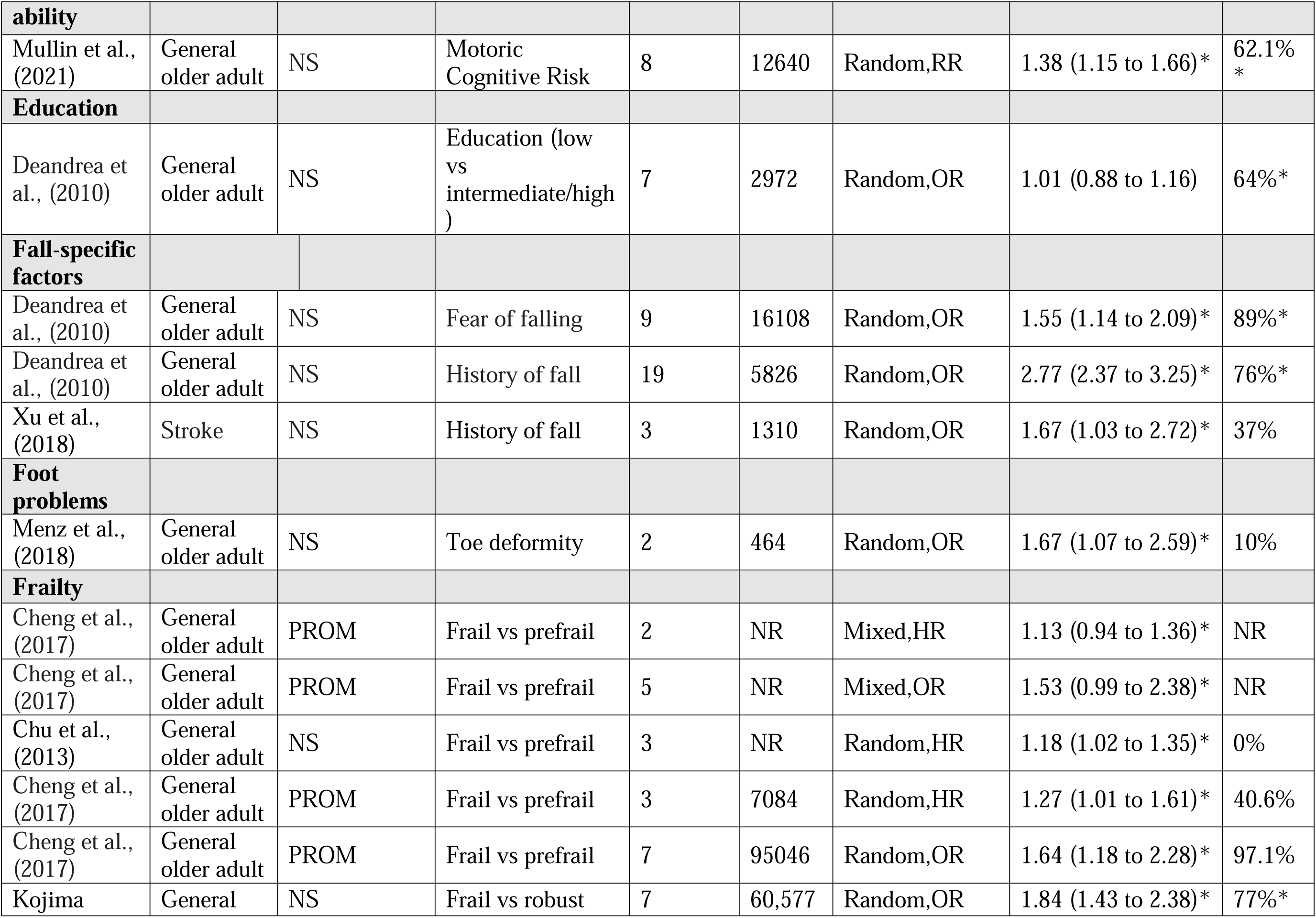
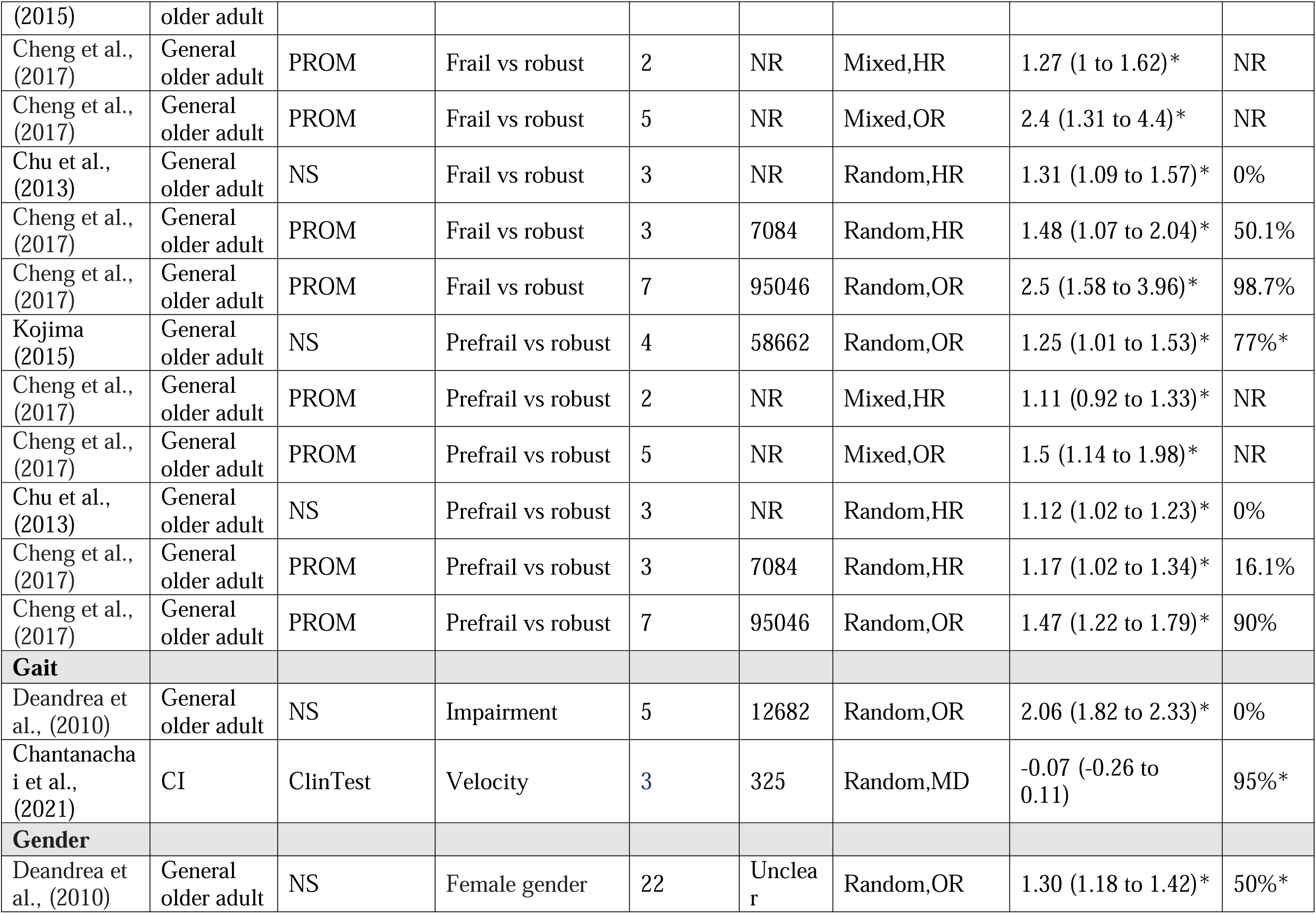
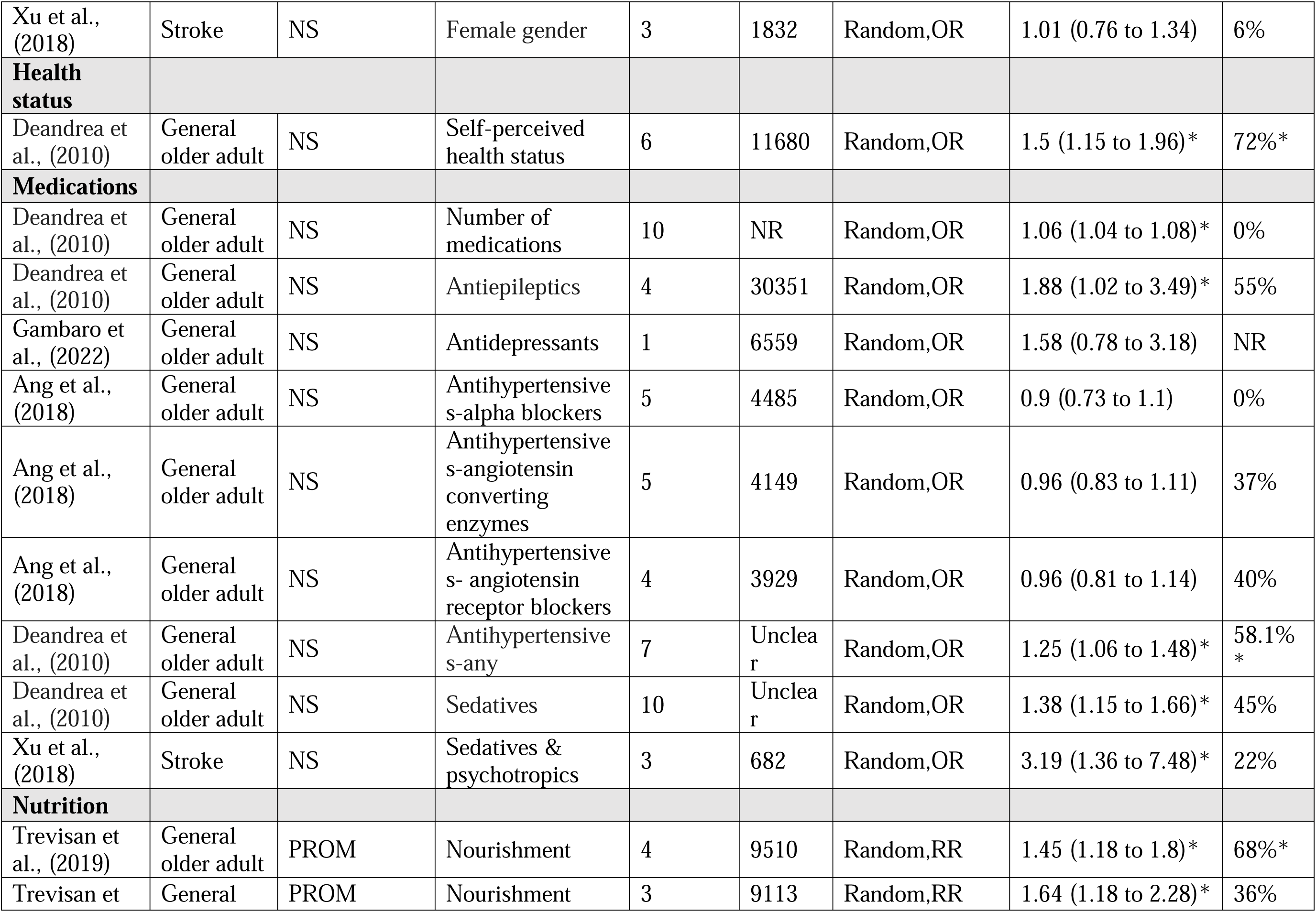
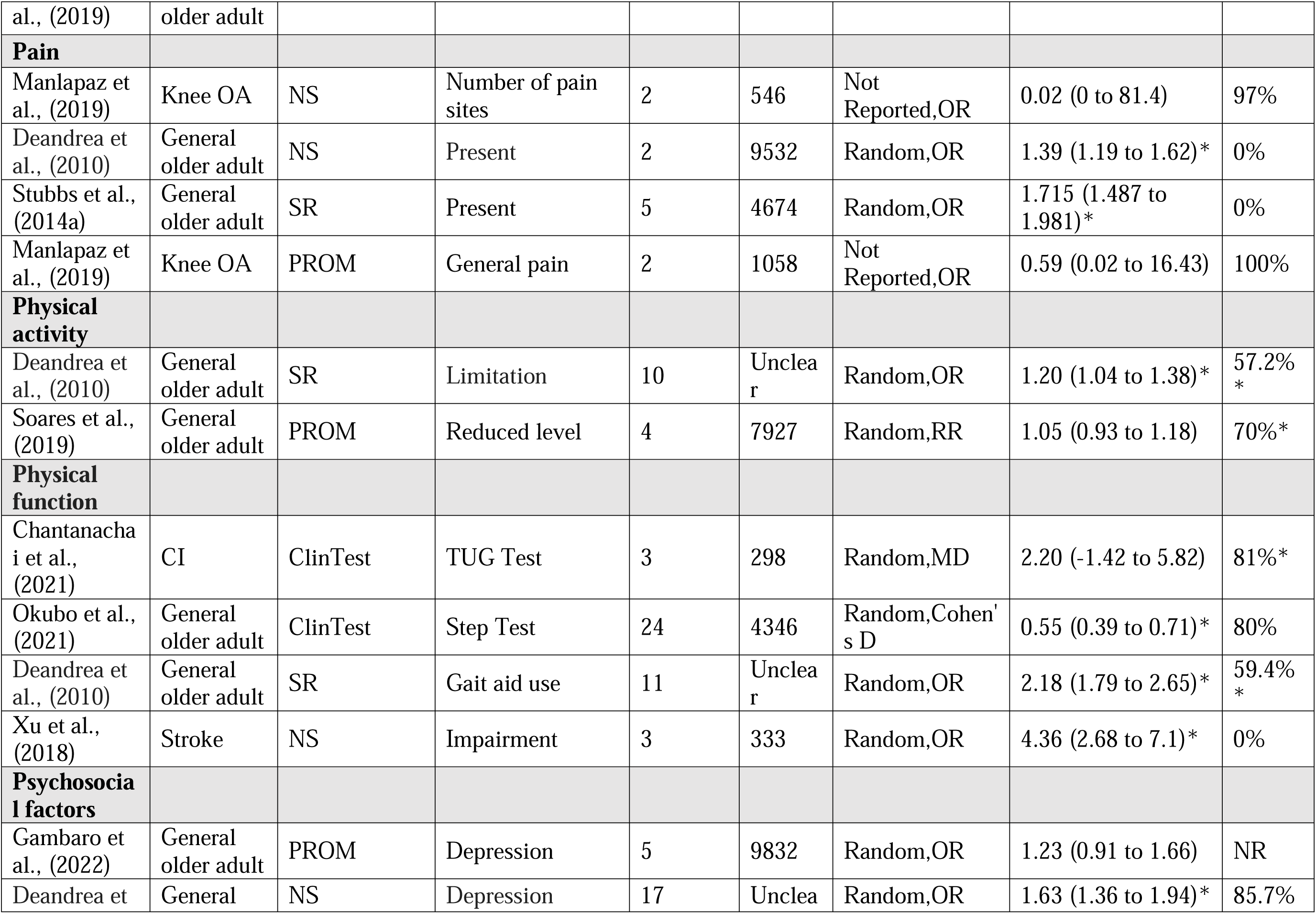
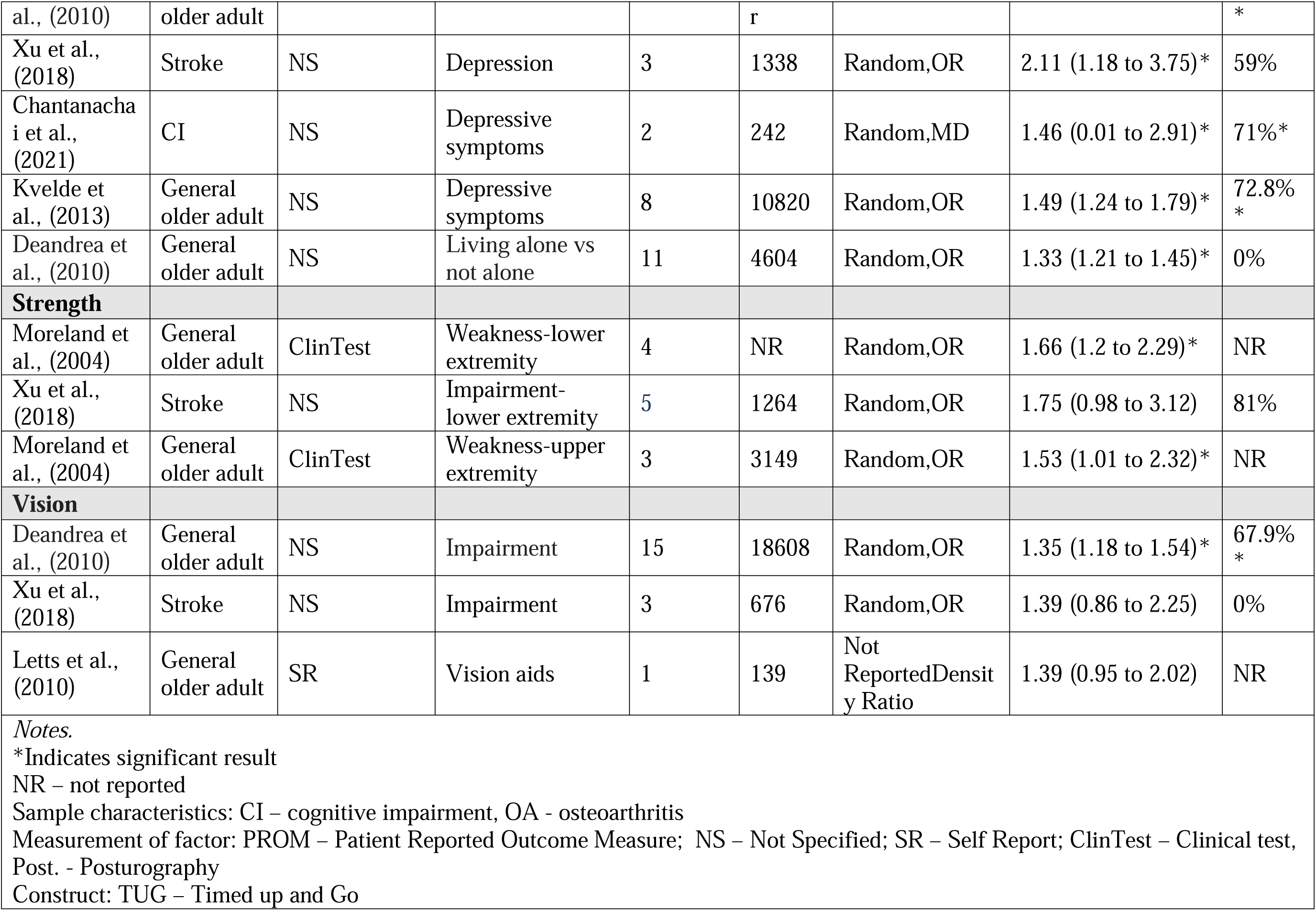
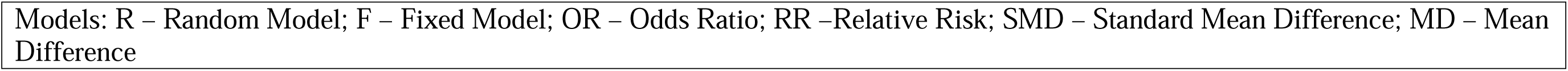
Description of Meta-Analysis Relationships Predicting Any Fall.

**Table 3.**
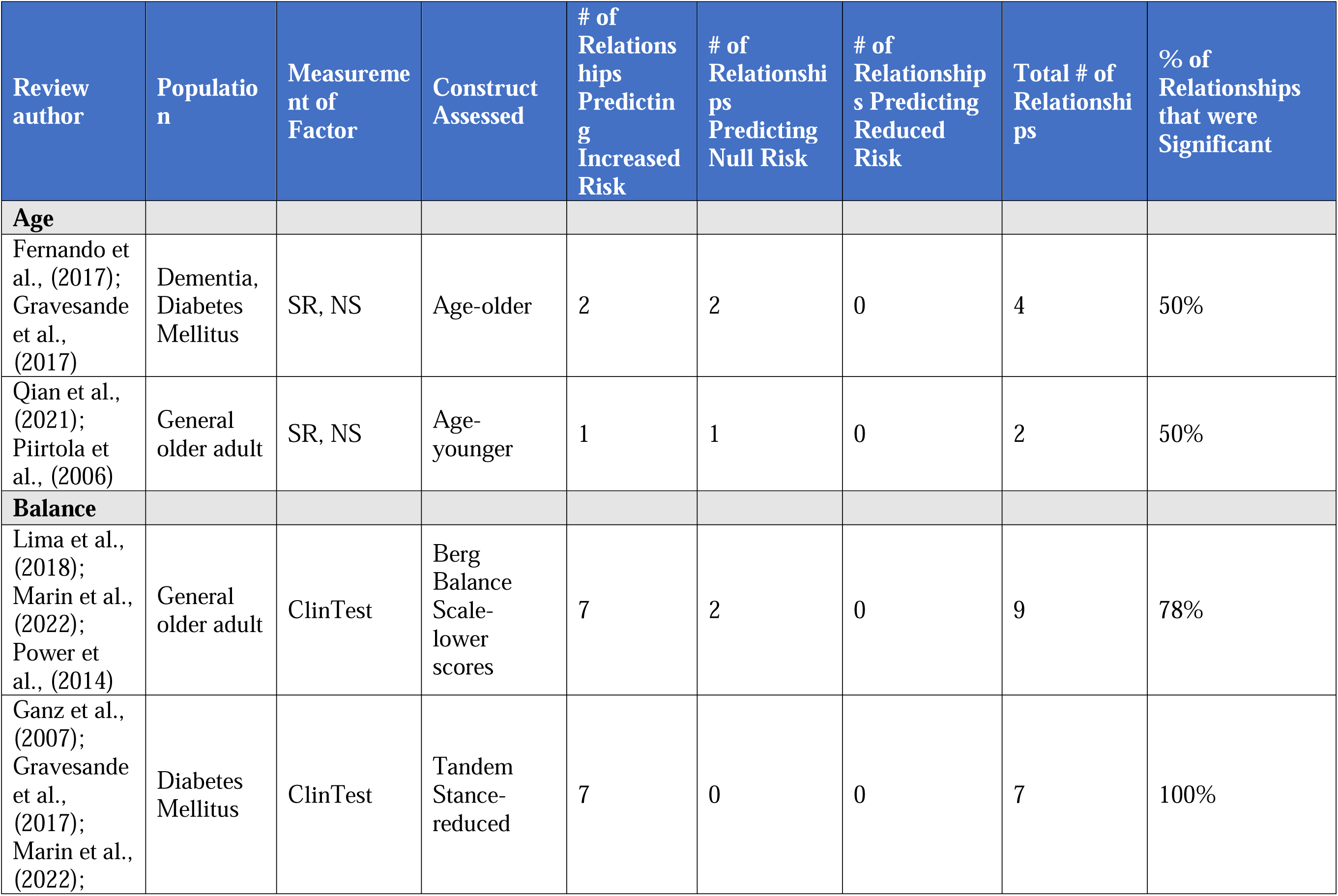
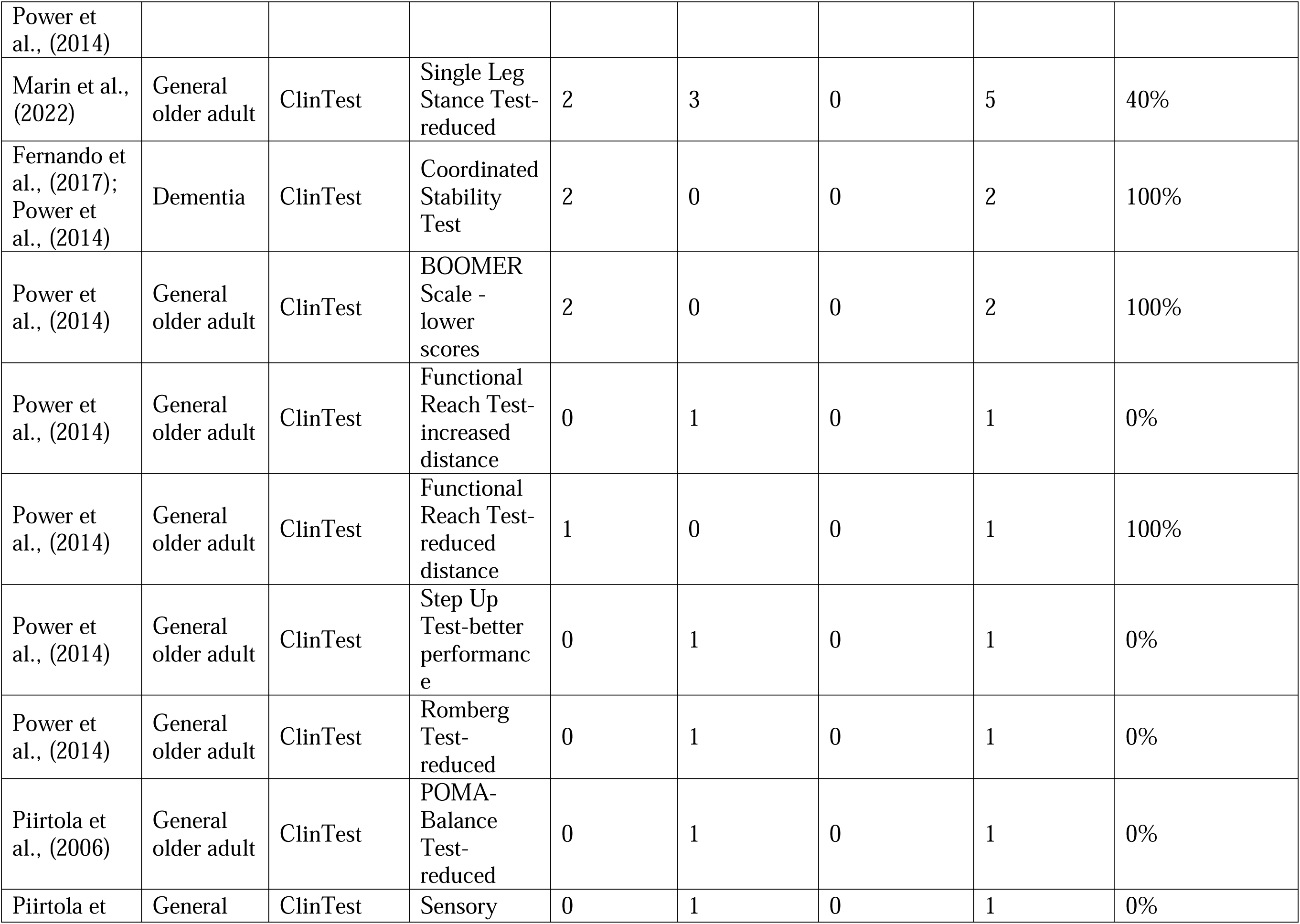
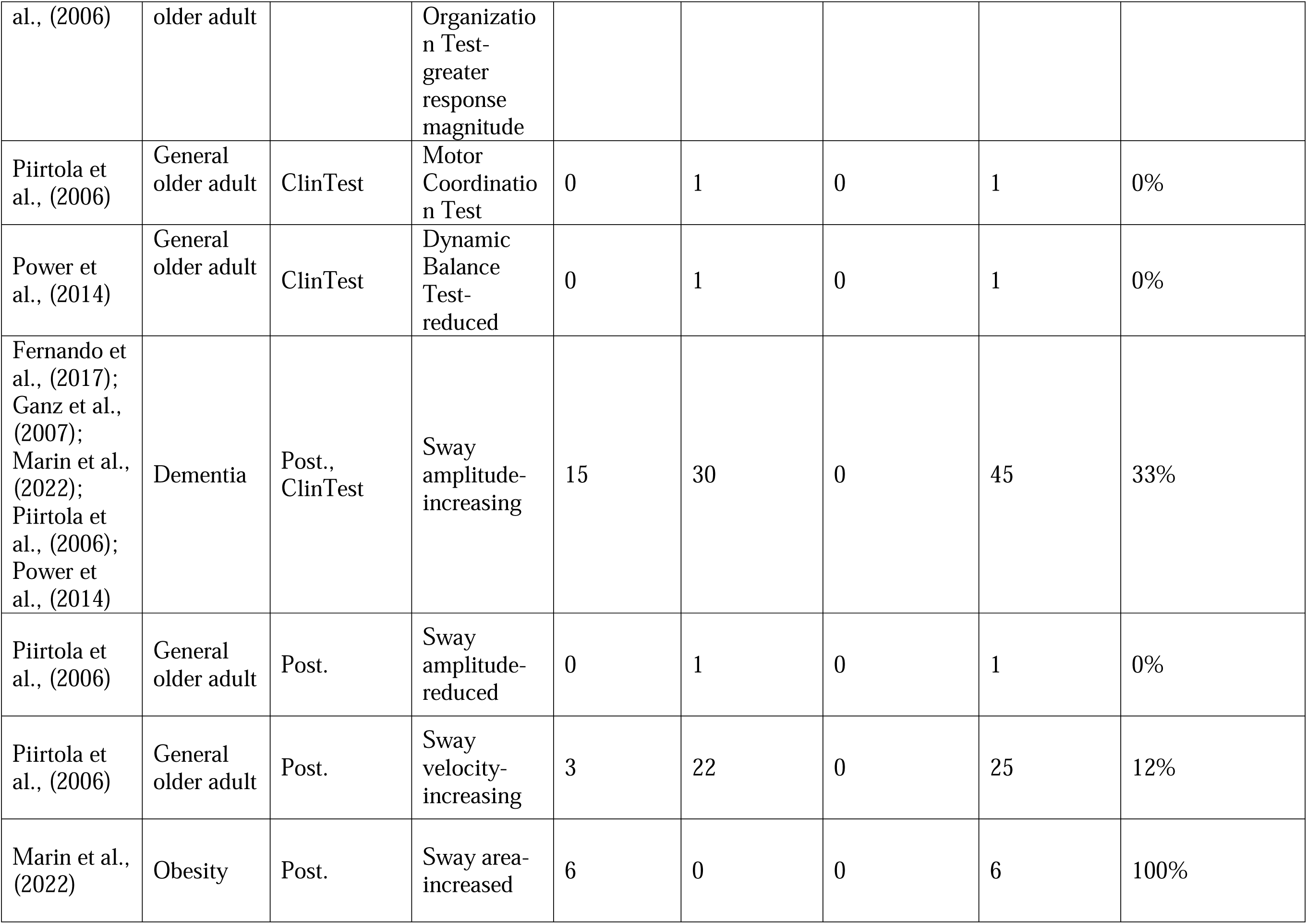
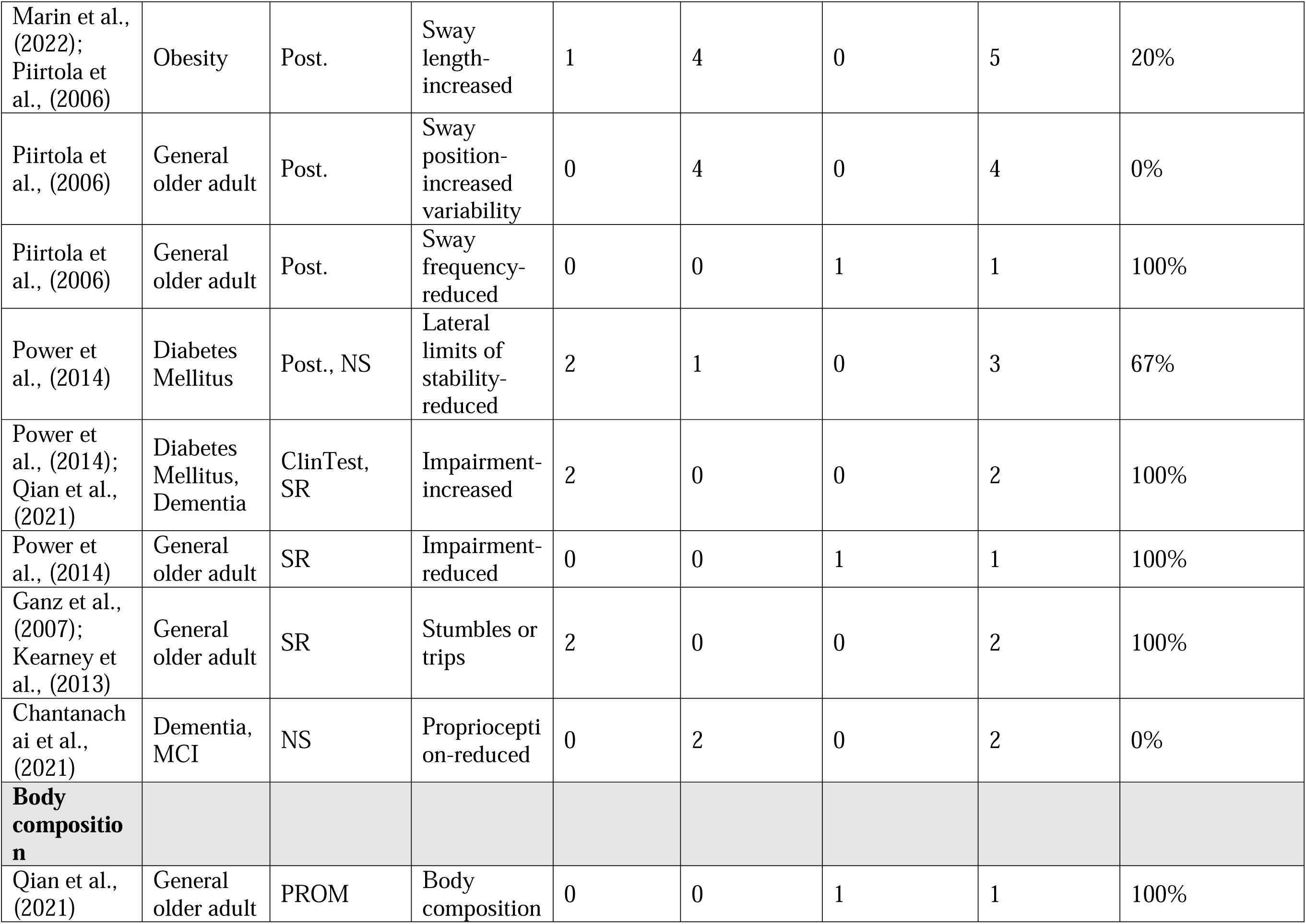
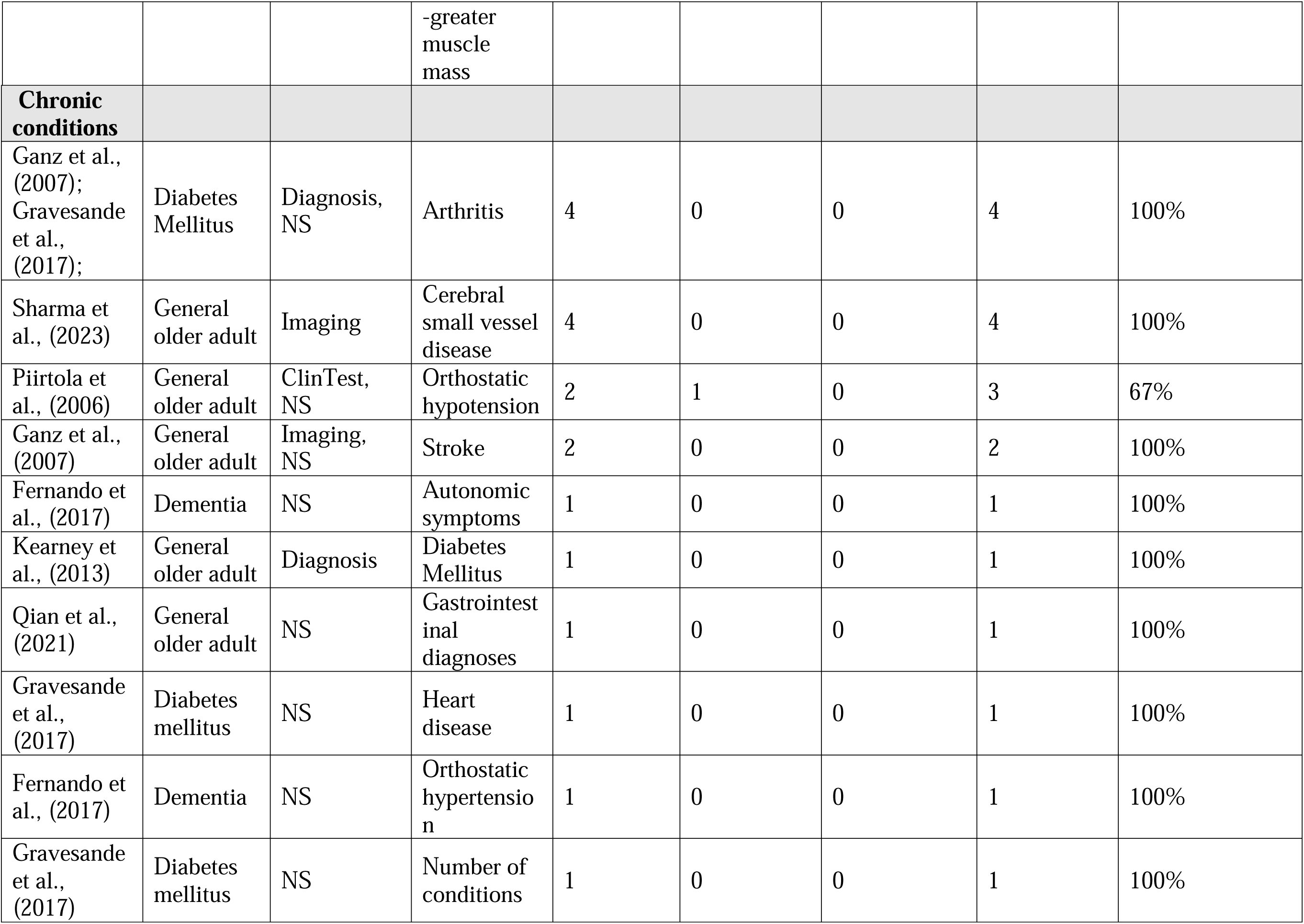
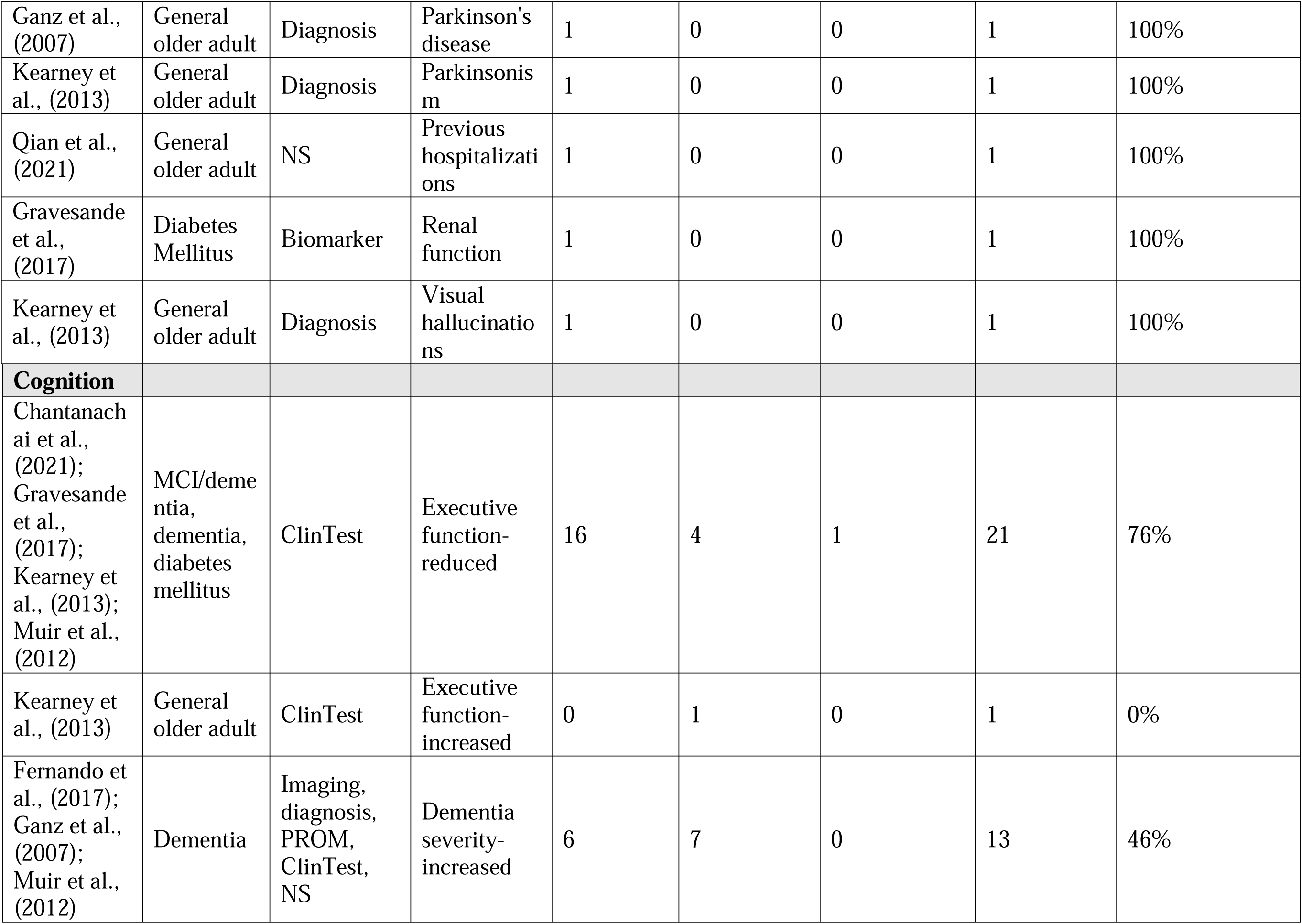
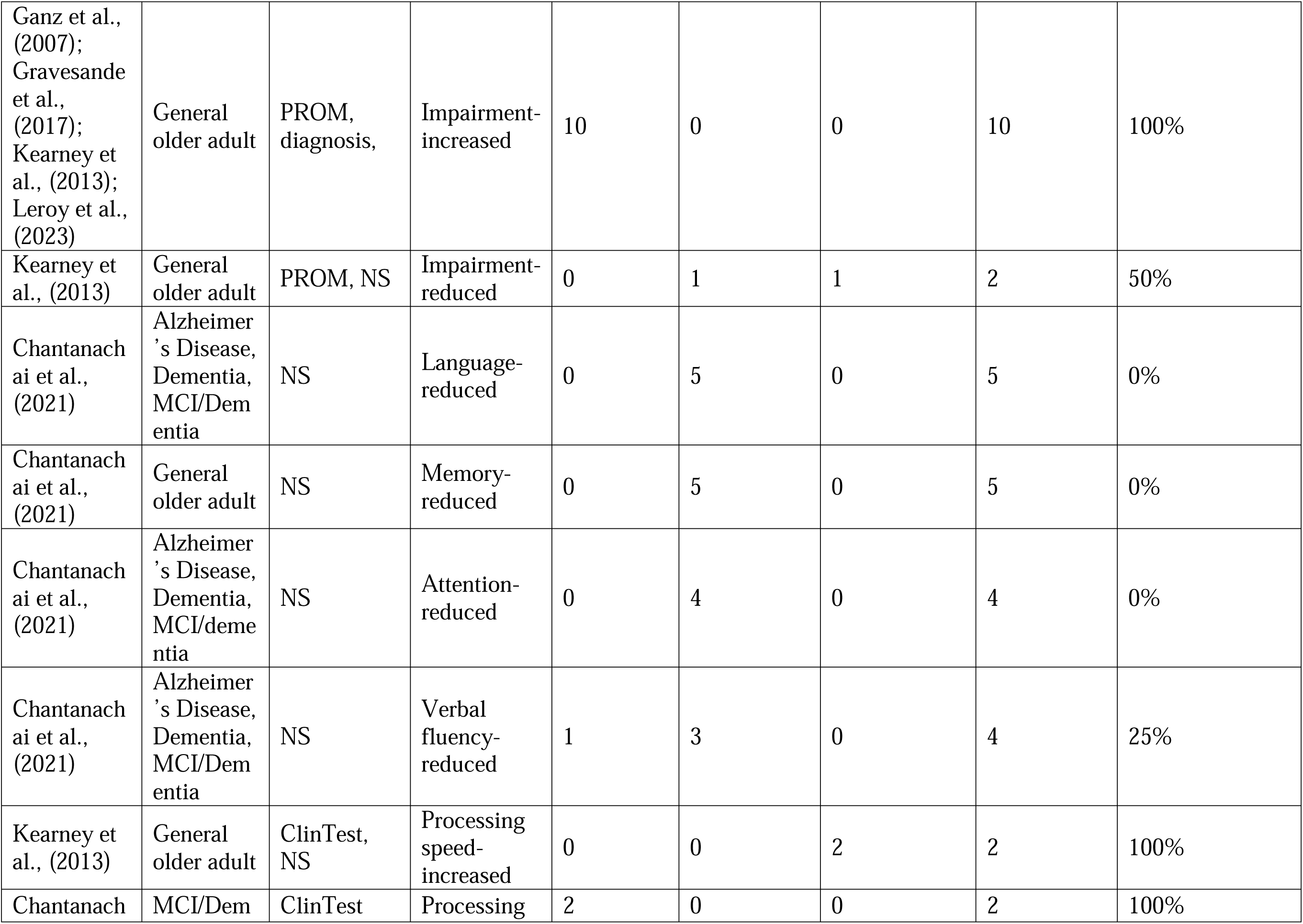
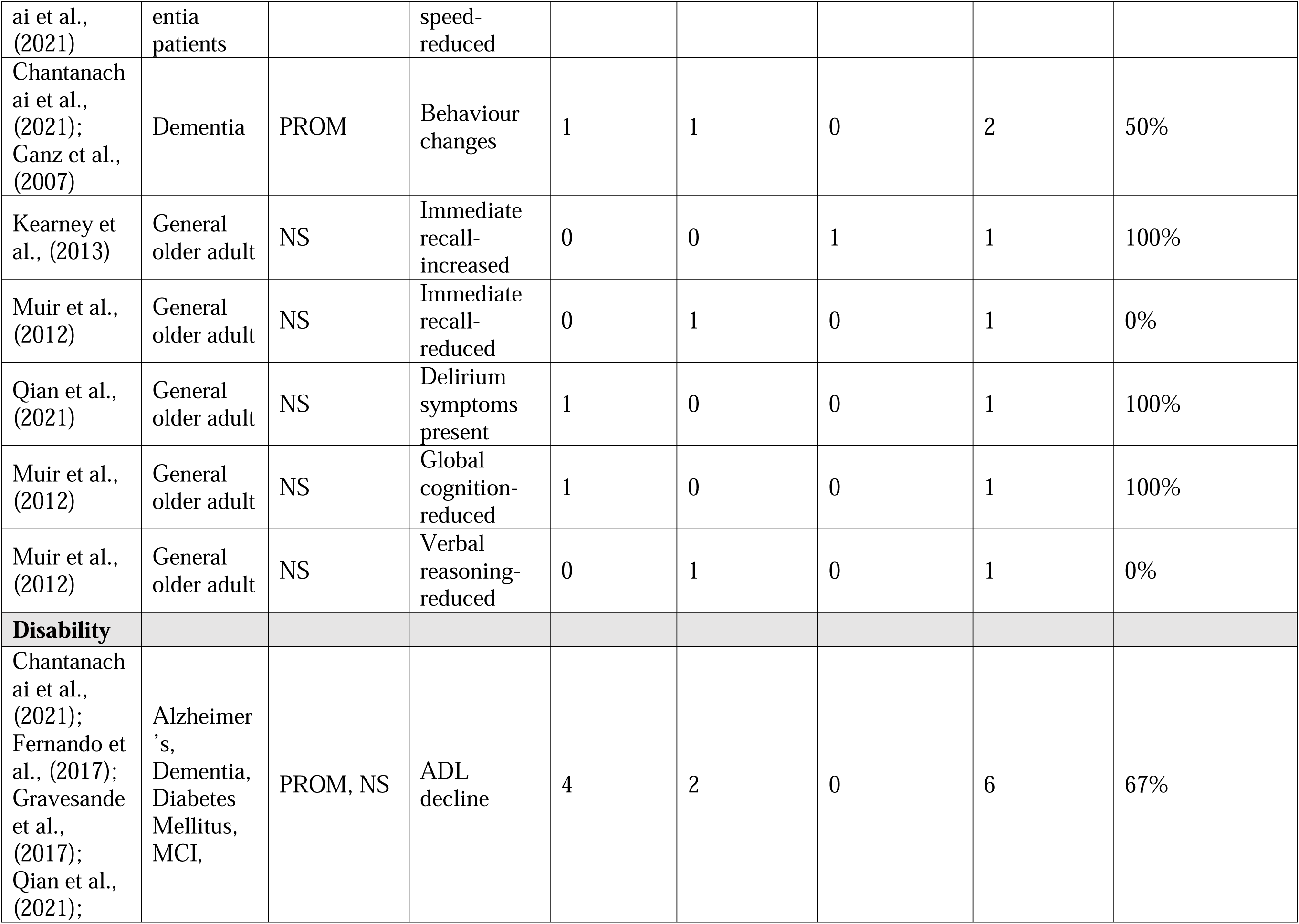
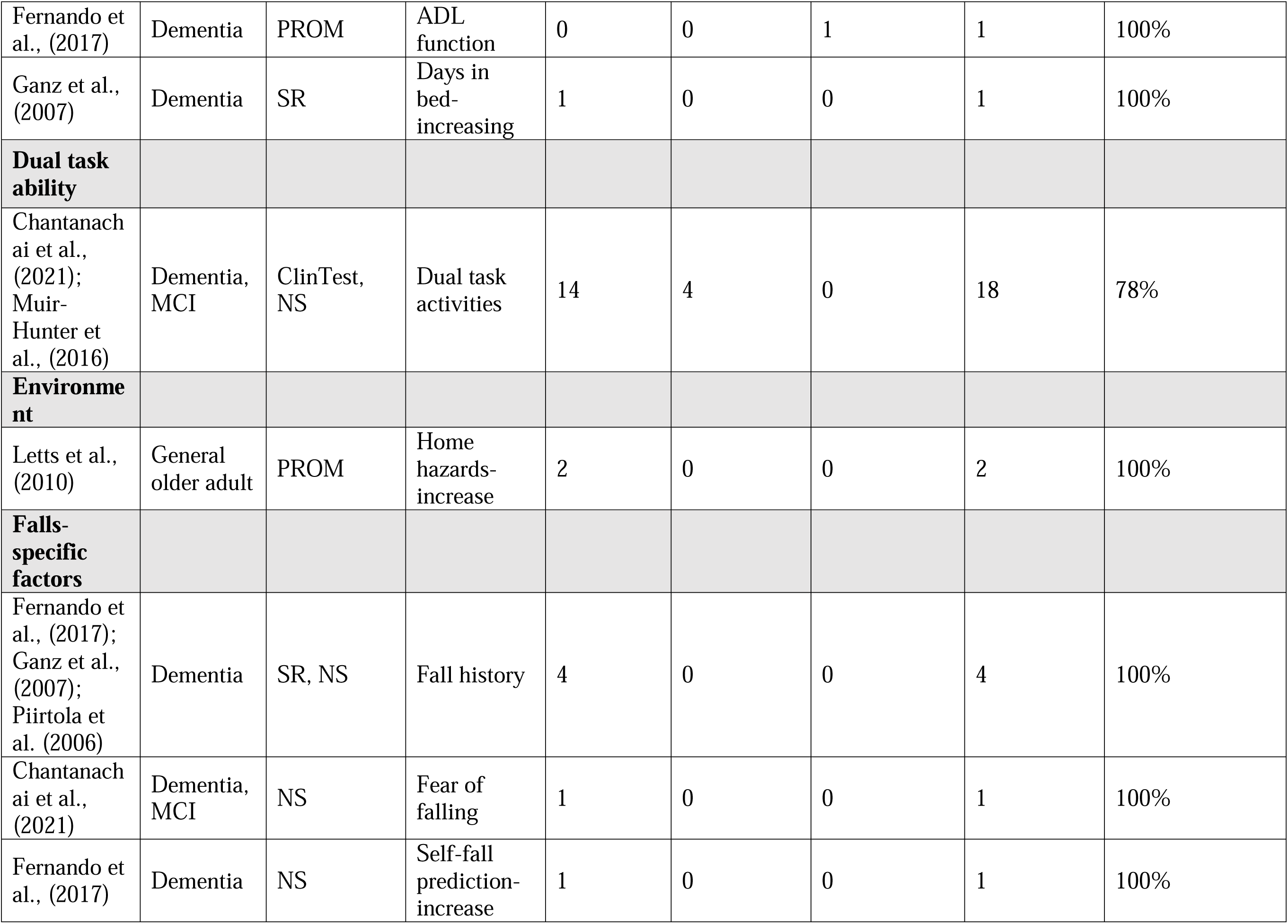
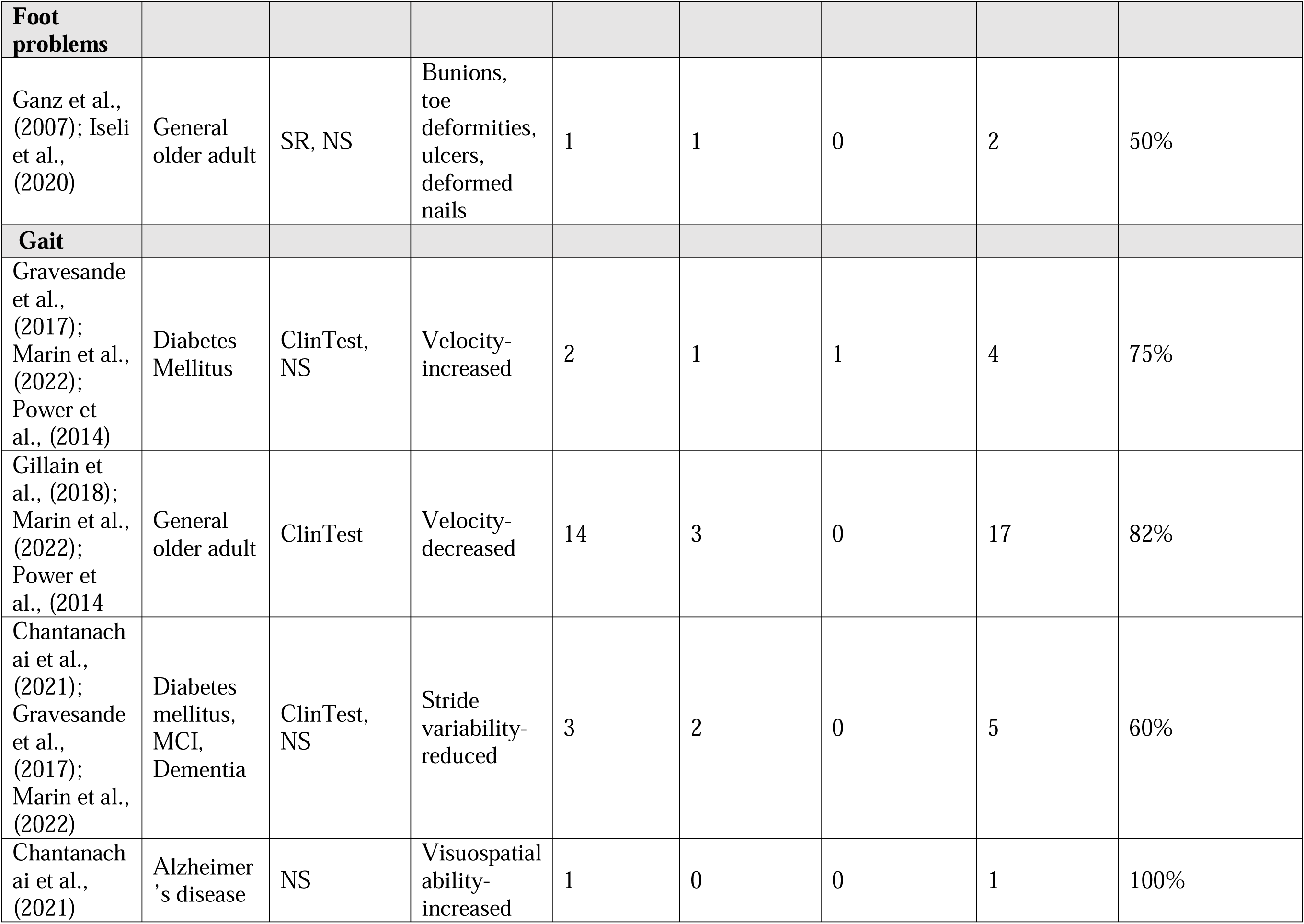
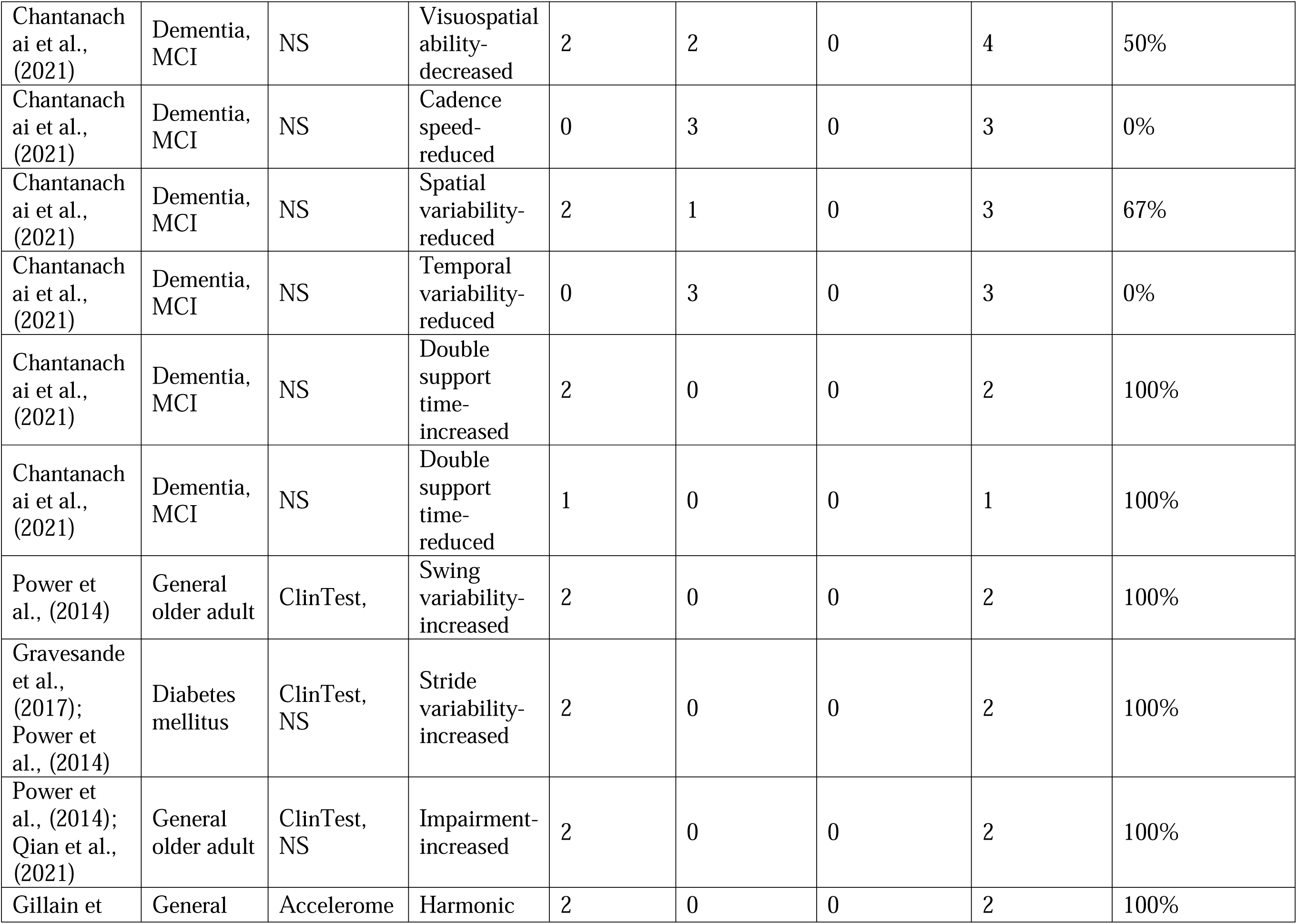
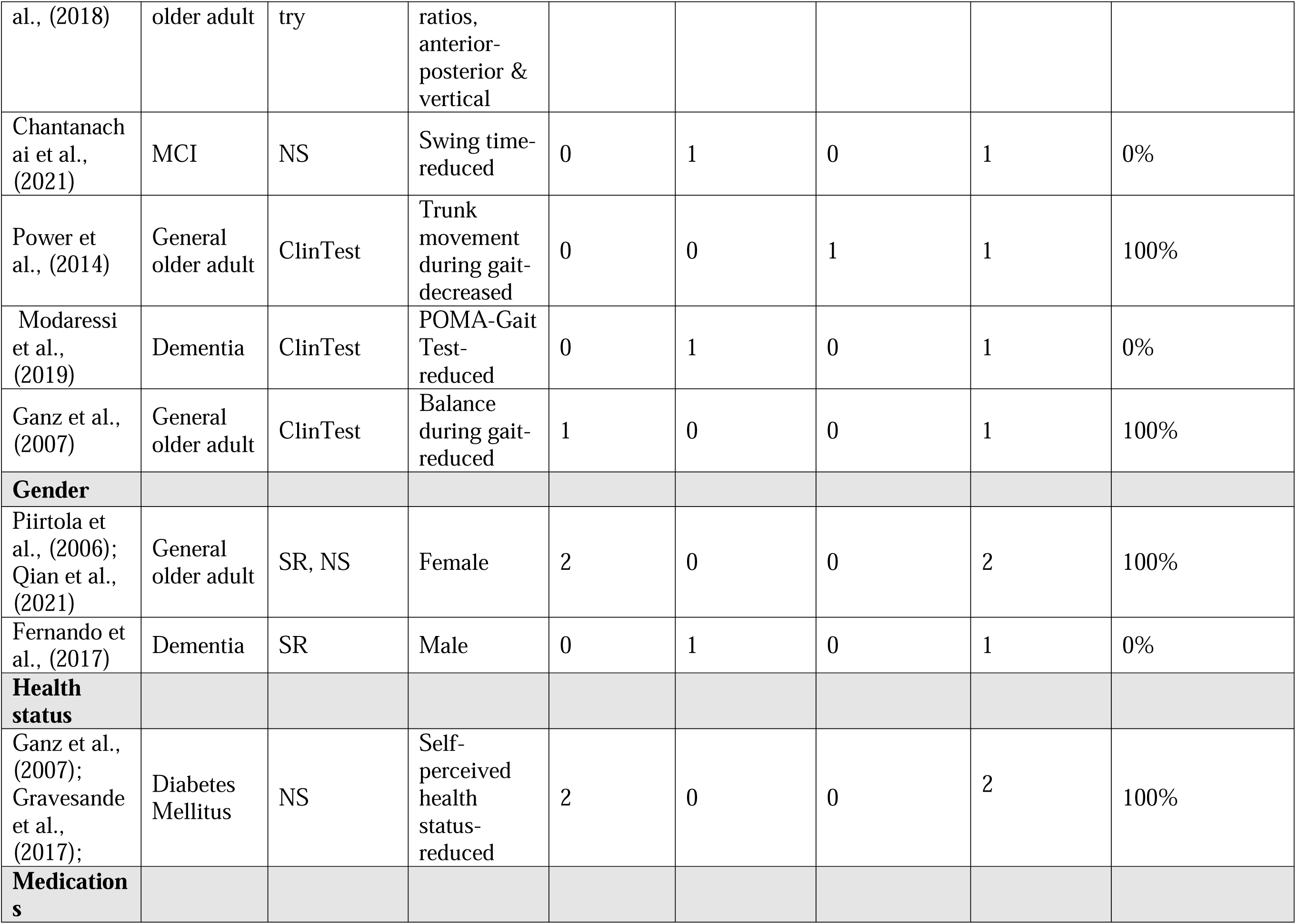
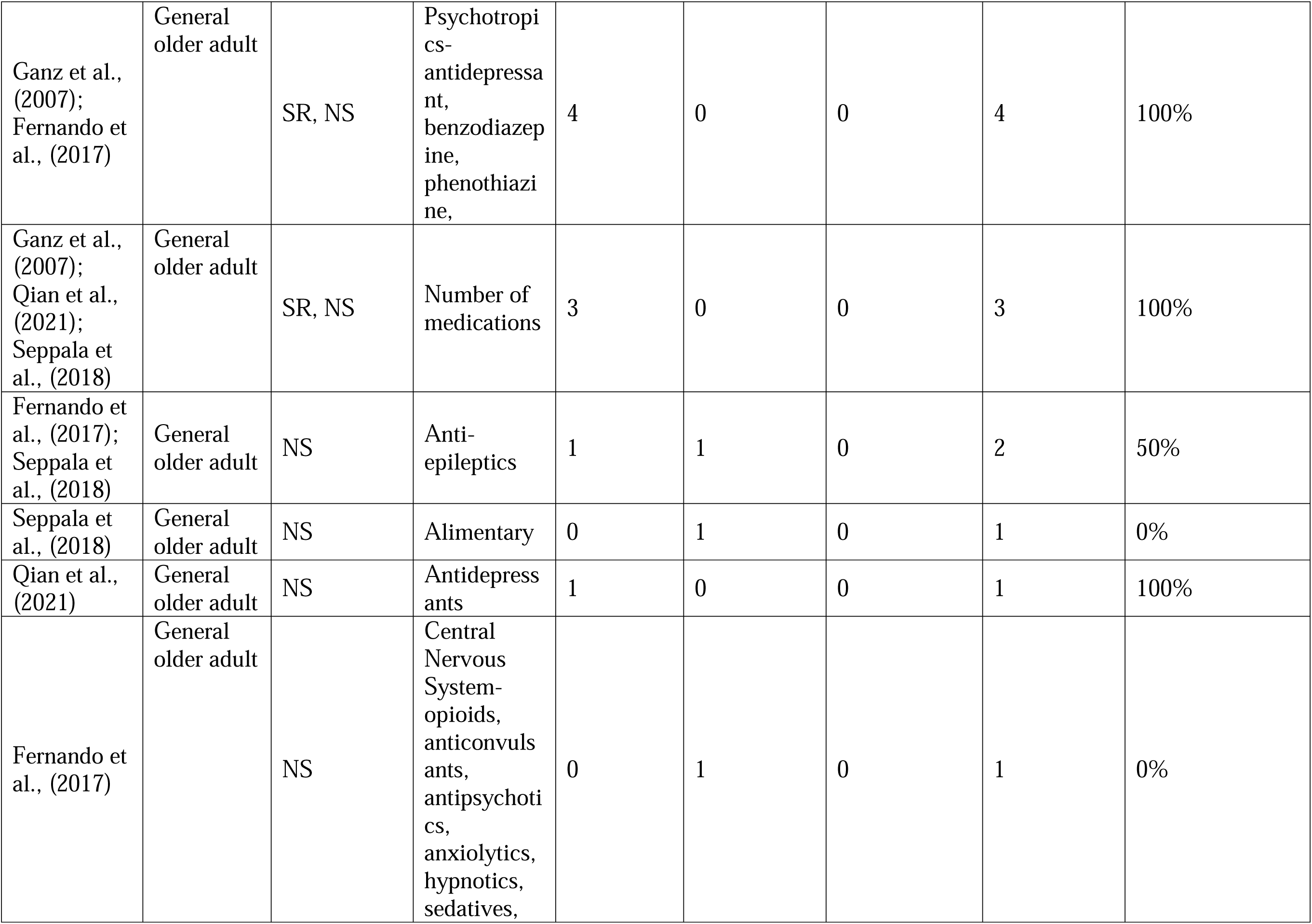
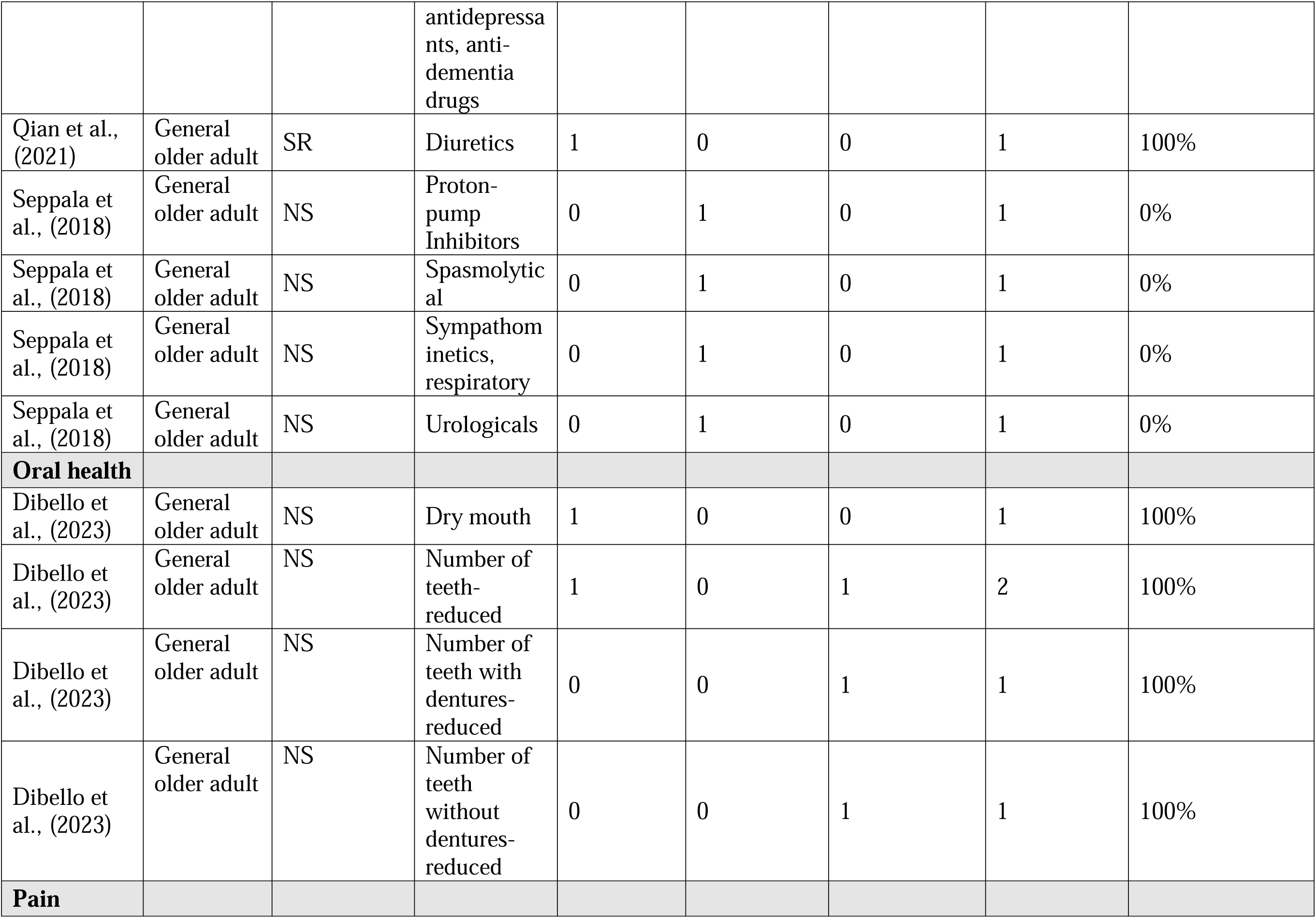
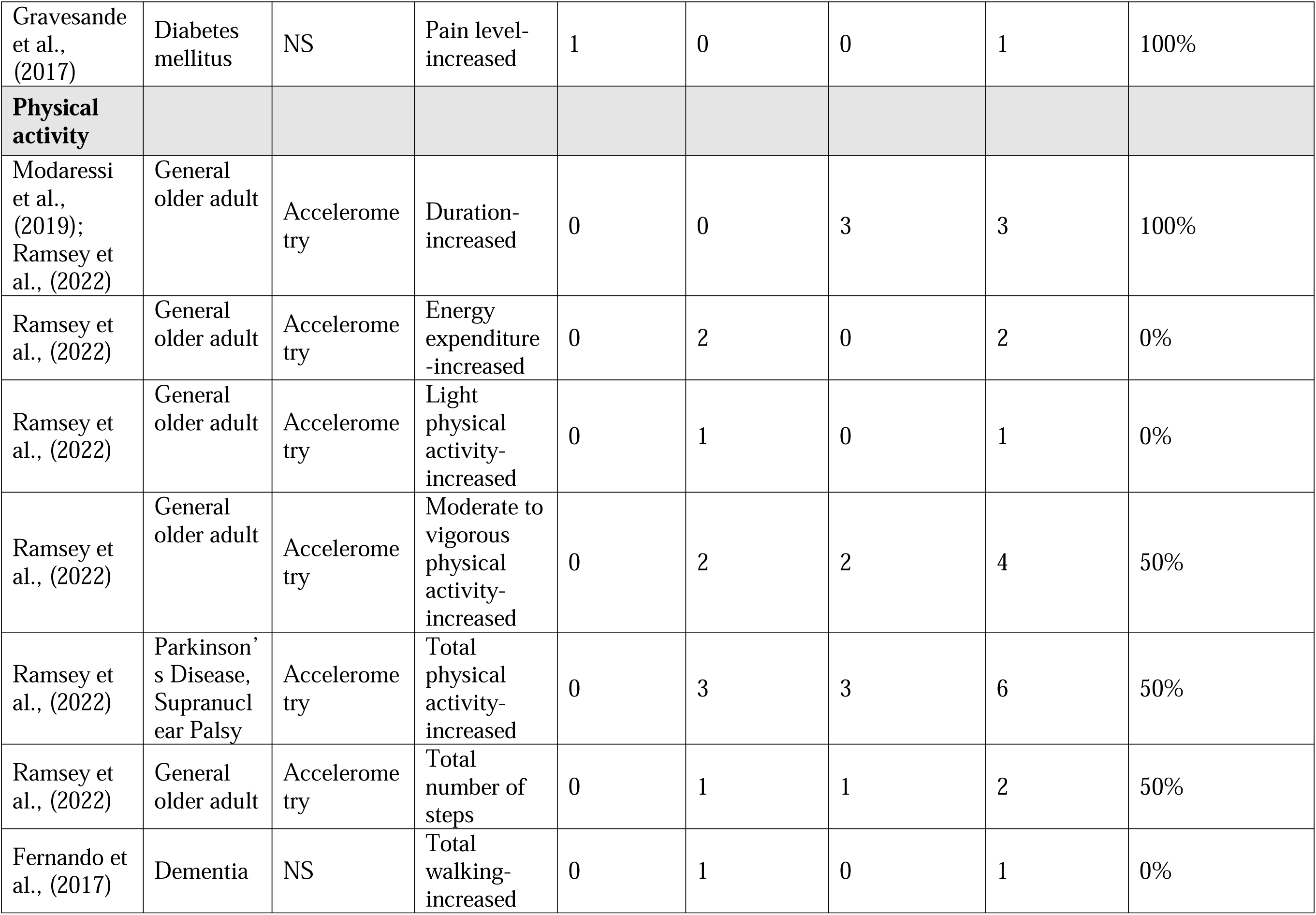
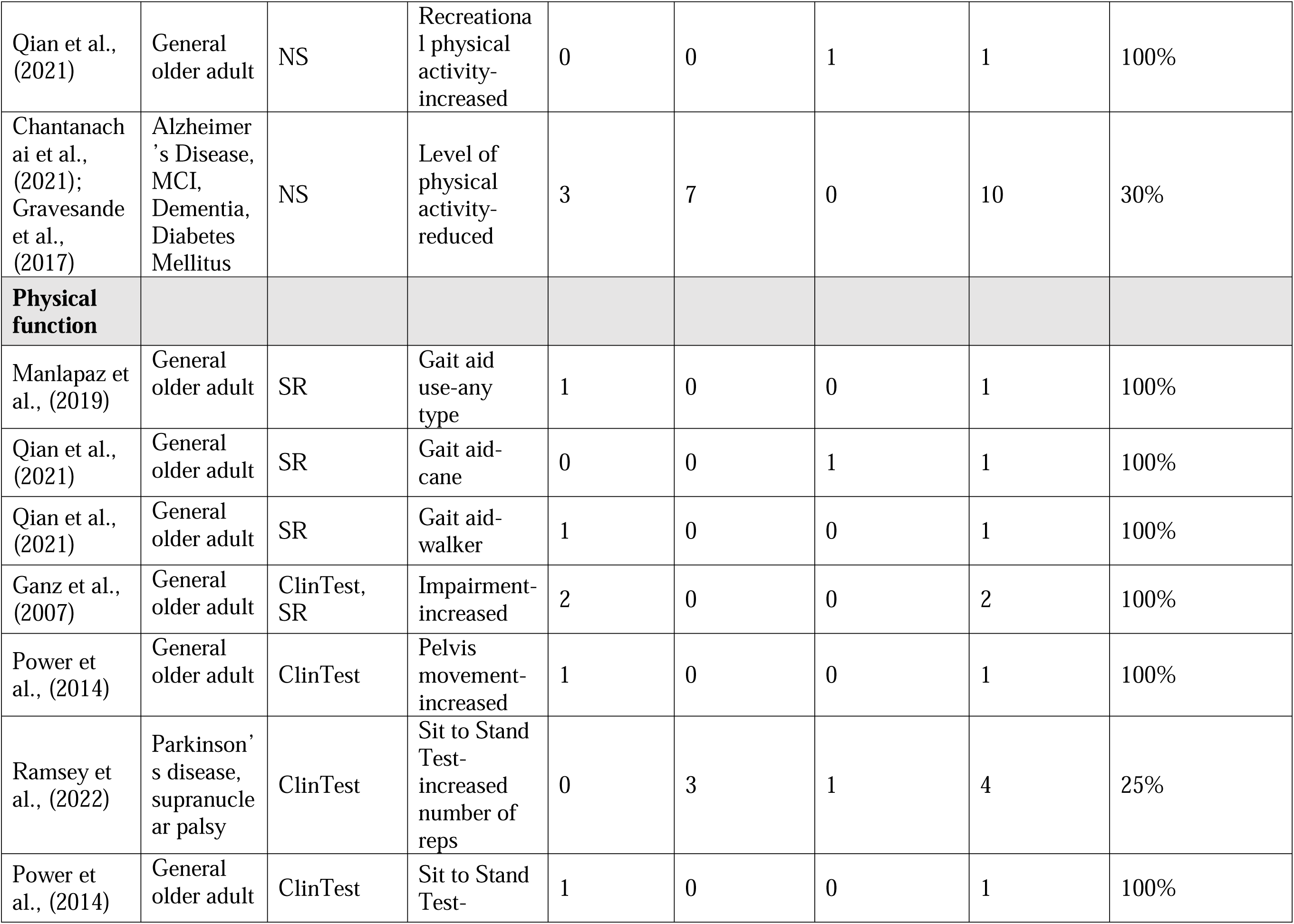
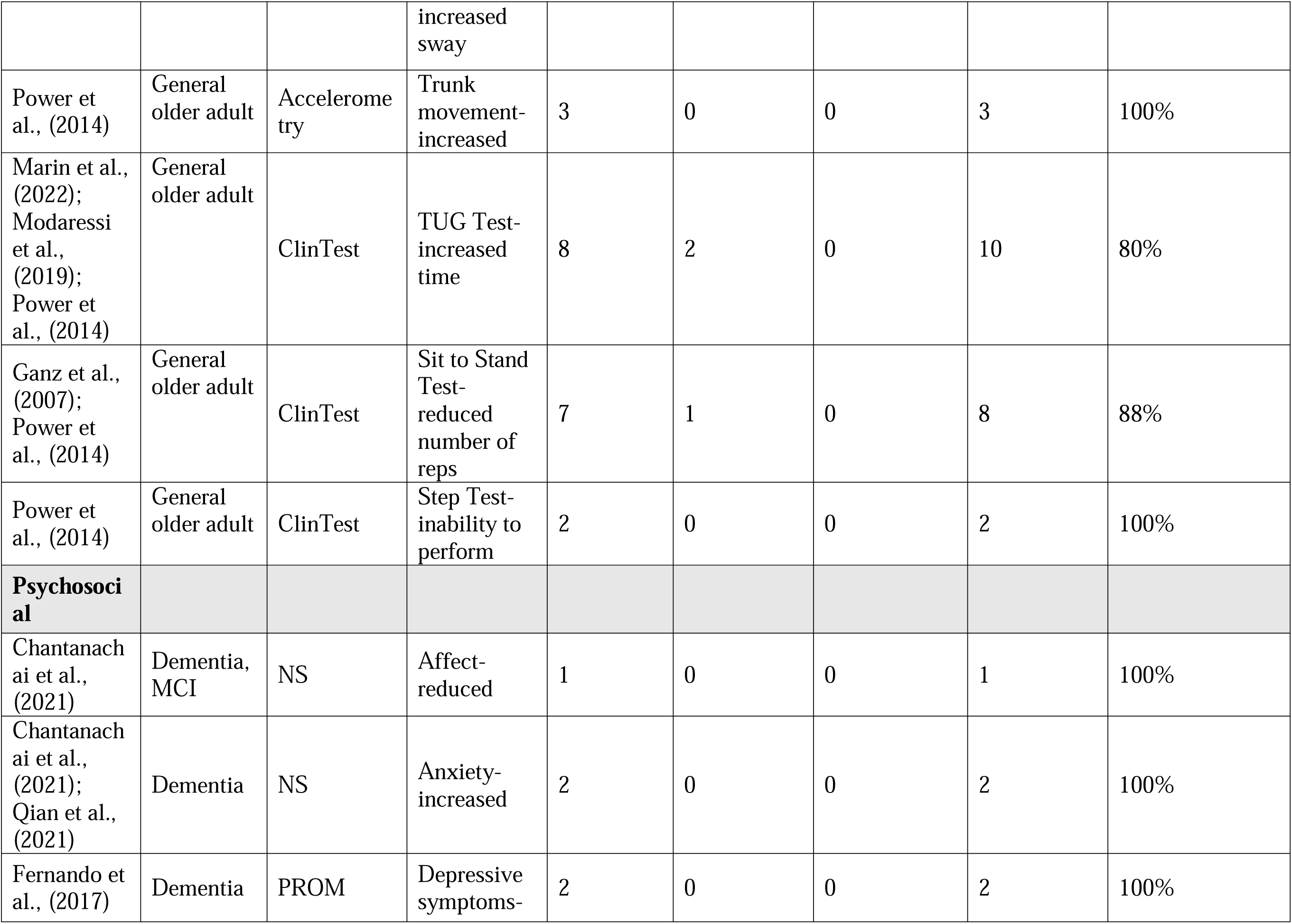
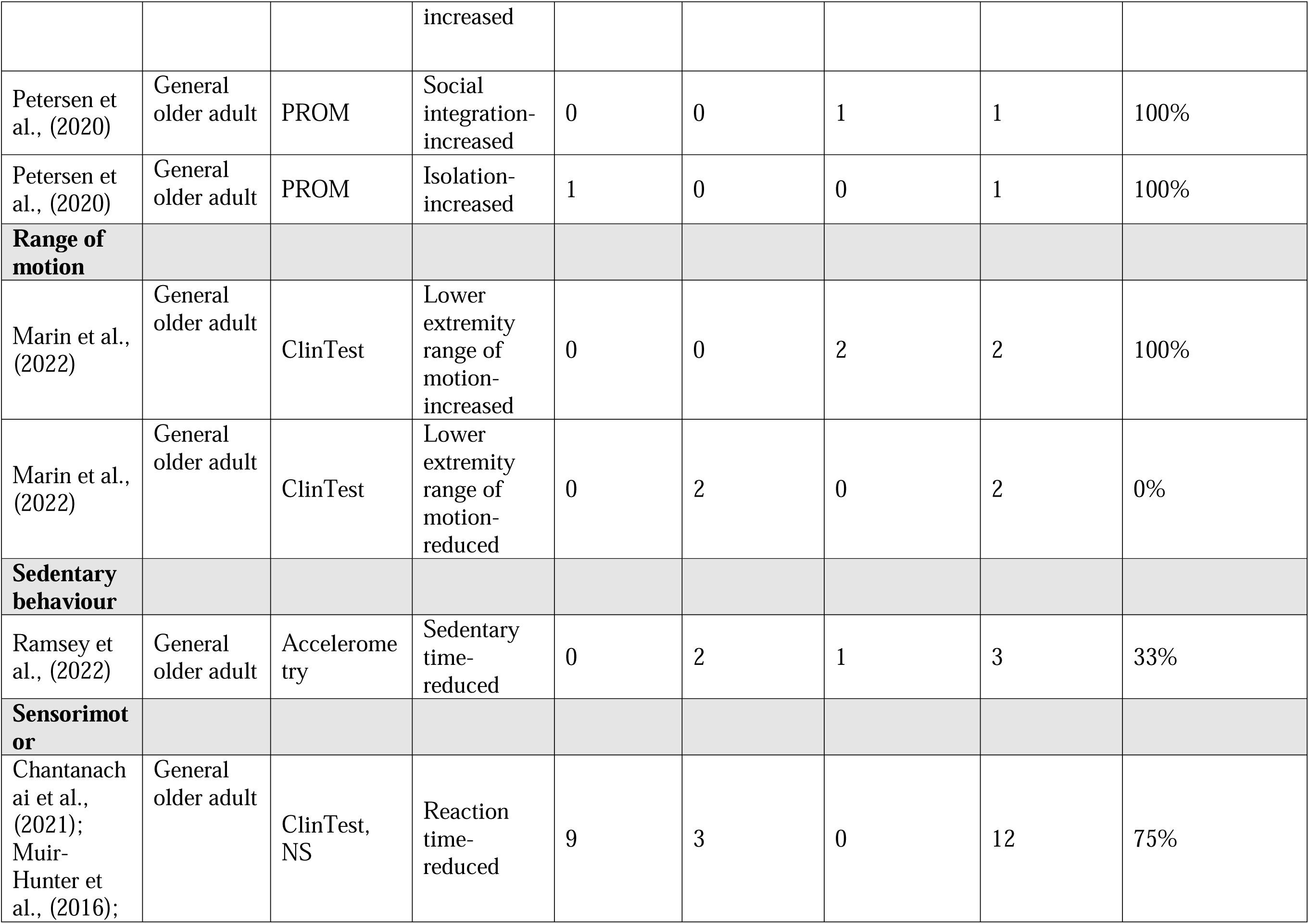
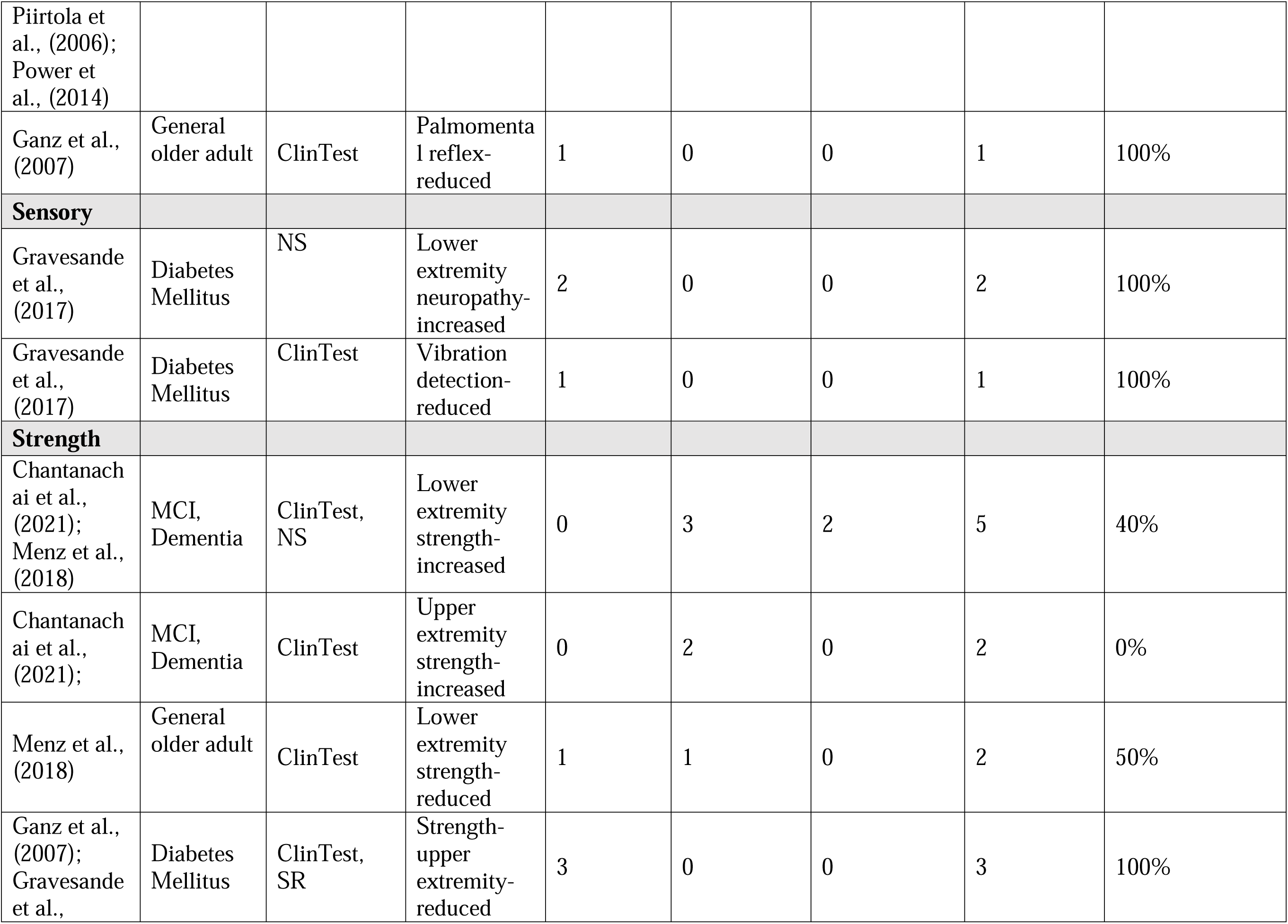
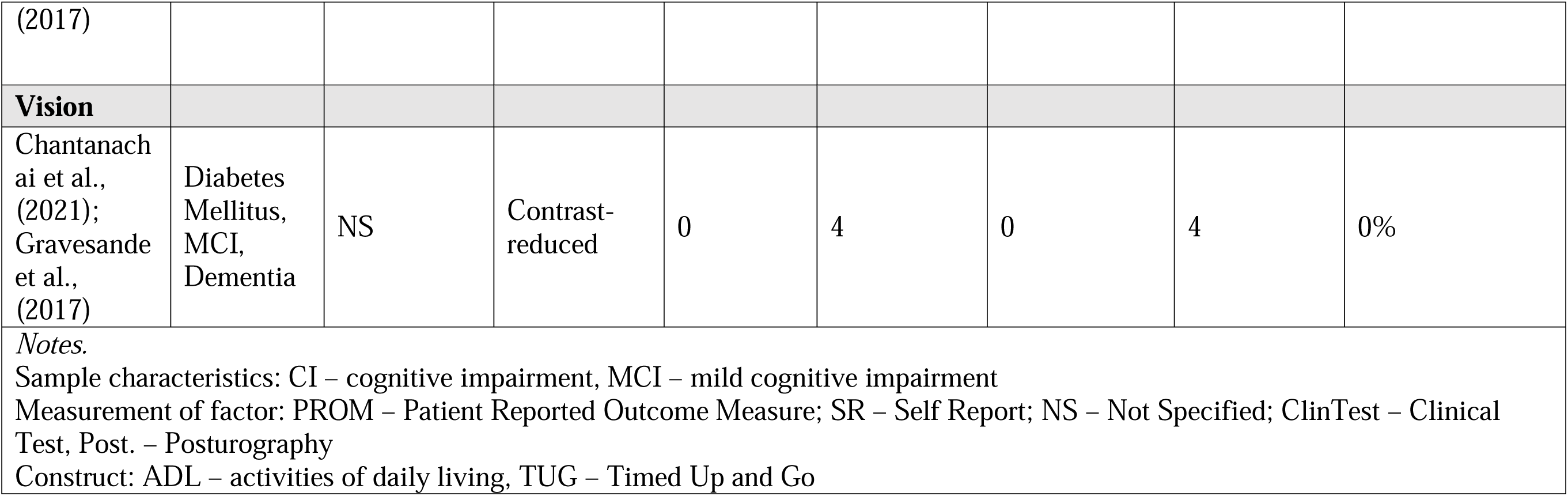
Description of Single Relationships Predicting Any Fall.

A total of 142 relationships examined balance, 12 from meta-analysis and 130 from narratively reported relationships. Global balance impairments were a significant predictor of falls in both meta-analysis relationships (ORs ranging from 1·98-3·87, SMD=0·62, Table 2) and single narrative relationships (n=3/3, 100%, Table 3). Five meta-analysis relationships from the review by Quijoux et al., ^14^ examining posturography measures of balance, including sway, position, and velocity, found that they were all significant predictors of falls. However, effect sizes were not reported and all relationships showed high heterogeneity (Table 2). The narrative relationships assessing balance via posturography also predominantly assessed increased sway amplitude and increased sway velocity (Table 3) with few relationships that predicted falls. For sway amplitude, only 33% of relationships (n=15/45) predicted an increased risk of falls and with sway velocity, only 12% of relationships (n=3/25) predicted falls. Regarding clinical tests of balance, two meta-analysis results, leaning and static balance, were predictors of falls with SMDs ranging from 0·60 to 0·62 (Table 2). The narrative results spanned 12 different balance-related clinical tests as predictors of falls. The most common tests were the Berg Balance Scale, where 78% of relationships predicted falls (n=7/9), Tandem stance, where 100% of relationships predicted falls (n=7/7), and Single Leg Stance, where 40% of relationships predict falls (n=2/5) (Table 3).

Multiple gait parameters were examined with gait velocity being the most common (n=1 meta-analysis relationship, n=21 narrative relationships). Regarding decreased gait velocity, the meta-analysis relationship did not demonstrate significantly increased risk (Table 2), yet 82% of narrative relationships were predictive of falls (Table 3). For faster gait velocity, 50% of relationships showed an increased risk of falls, while 25% showed reduced risk of falls, suggesting a U-shaped relationship within the narrative results. While other gait parameters, including variations in stride, swing, spacing, and balance, were all predictive of falls, there were few relationships (n£2) examining these outcomes (Table 3).

Measures of physical function predicted falls, however few had more than four relationships examining a single predictor. The most common measure was the Timed up and Go, with 80% of relationships showing a slower time predicted falls (n=8/10; Table 3). Similarly, performing fewer sit to stands was also predictive of falls, with 88% of relationships found to be significant (n=7/8). In contrast, completing more sit to stands was not protective for falls. Other measures of physical function included gait aid use (n=3), greater trunk movement (n=3), general impairment (n=2), and step tests (n=2), all of which had 100% of relationships significantly predictive of falls (Table 3).

Investigations of physical activity as a risk factor for falls largely explored exertion-based classifications, where only physical activity duration emerged as protective against falls (n=3, 100%, Table 3). Greater energy expenditure, light physical activity, and total walking had few relationships reported and none showed a protective effect (0%). For moderate to vigorous physical activity and greater total physical activity both had 50% of relationships with a protective effect against falls.

In examining participants’ ability to perform dual task activities (i.e., combined motor and cognitive tasks), 79% of relationships significantly predicted falls (n=15/19). Based on the available information, it was not feasible to distinguish whether the motor or the cognitive task was considered the primary task (Table S1·3).

The results for strength-related risk factors depended on the direction. Reduced strength in the upper extremities predicted falls in both meta-analysis (OR= 1·53; Table 2) and narrative relationships (n=3/3, 100%; Table 3). Meanwhile greater strength in the upper and lower extremities examined in narrative relationships was only protective against falls in 0/2 (0%) and 2/5 (40%) relationships, respectively (Table 3)

In terms of cognition, 57% of relationships were significant (Table S1·3), with 100% of relationships examining greater general cognitive impairment showing an increased risk of falls in both meta-analysis (ORs ranging from 1·32-1·75) and single relationships (n=10/10, 100%). However, certain cognitive functions had a higher proportion of significant relationships, specifically, executive function and processing speed were both predictive of falls (Table 2, Table 3). Executive function examined within MCI/dementia populations had 76% of relationships significant for predicting falls (n=16/21). Processing speed was also predictive of falls both as a protective factor (i.e., increased processing speed; n=2/2) and as a risk factor (i.e., reduced processing speed; n=2/2) with 100% of relationships reaching significance. Cognitive factors that reported more than 4 relationships and were not predictive of falls included poor language capacity (n=0/5, 0%), poorer memory (n=0/5, 0%), reduced attention (n=0/4, 0%), and poorer verbal fluency (n=1/4, 25%).

There was a wide range of chronic conditions found to predict falls. Conditions where more than one relationship was reported included diabetes (n=2, ORs ranging from 1·19 to 1·59), stroke (n=3, ORs ranging from 1·11 to 1·61) and urinary incontinence (n=2, ORs ranging from 1·4 to 1·54). Of these conditions, diabetes was a consistent predictor of falls, with both relationships found to be significant (Table 2).

Medications were grouped according to how each review classified them. Antihypertensives were most common and were insignificant in all but one relationship (n=4; ORs ranging from 0·9 to 1·38, Table 2). Sedatives as predictors for falls were examined in 2 relationships and while both relationships were significant (ORs ranging from 1·38 to 3·19, Table 2) with moderate-low heterogeneity, one of these relationships also included psychotropics more broadly. Psychotropic medications had the highest number of reported relationships (n=4) and 100% of these were predictive for falls (Table 3). Qian et al., ^15^ reported antidepressant medications as a standalone category, despite being classified within psychotropics, with 100% of relationships significantly predictive of falls (Table 3). That said, central nervous system medications, which also consist of antidepressant medications, were not predictive of falls (n=1, Table 3).

All relationships examining frailty via meta-analysis were significant. The relationships examined different frailty classifications, including frail vs prefrail, frail vs robust, prefrail vs robust. The comparison between frail versus robust had the largest range of effect sizes (1·27 to 2·5, mix of HR and ORs, Table 2).

Predictive validity of outcome measures was also evaluated as part of this review. Of the 22 measures reported, which included both physical performance tests and patient reported outcomes (e.g., Berg Balance Scale, Falls Efficacy Scale International) few tests reported AUC values and none demonstrated AUCs >0·7 for any falls (Table S1·6). For recurrent falls, Functional Gait Assessments, POMA, and TUG had AUCs of 0·74, 0·76, 0·73, respectively (Appendix 2, Table S2·5).

Reviews were mixed in terms of risk of bias. The majority of reviews (n=31, 54%) were rated as low risk of bias, while 26 (46%) reviews were rated as high risk of bias. High risk of bias was most commonly found in identification of studies, largely due to weak search strategies. Study eligibility and data collection and appraisal criteria had the lowest risk of bias, See Table S1·2 for more details.

Across the entire review, there was relatively low overlap between studies, with a calculated CCA of 1%. The CCAs of the five most common risk factors were as follows: balance=2%, cognition=2%, gait=2%, chronic conditions=1%, and physical function=2%.

## Discussion

This umbrella review represents the most comprehensive summary of predictors of fall risk among community-dwelling older adults. We summarized the results from 57 reviews reporting on 356 prospective studies that examined 29 different fall risk factors, including 963 relationships. The ten most common predictors of falls were balance, cognition, gait, physical function, physical activity, medications, chronic conditions, dual task ability, frailty, and strength. Of these, frailty, chronic conditions, physical function, dual task ability, and gait had the greatest proportion of relationships reaching significance (i.e., ≥60%). Over half of the fall risk predictors were mobility-related constructs (e.g., balance, physical function), and there was a paucity of evidence examining psychosocial, sociodemographic, and environmental factors.

Mobility limitation is a well-established risk factor for falls and is a key component of the World Falls Guidelines algorithm for fall risk screening,^6^ which explains why over half of the risk factors examined in this review were mobility-related measures. Of these, balance was the most examined predictor of falls in this review, accounting for 25% of all relationships. Yet, the type of balance testing seems important for predicting falls. Results were most consistent for general balance impairment and clinical balance tests (i.e., Berg Balance Scale) across both meta-analyses and narrative relationships. However, measurement of balance via posturography had mixed results, which is likely due to the number of different parameters reported, limiting our ability to synthesize the results into meaningful conclusions. Other mobility-related measures included gait, where variability (i.e., stride, swing, visuospatial ability) and slower gait velocity were consistently predictive of falls, as well as physical function both when assessed via clinical tests and as general impairments. Reduced strength in the upper extremities also predicted falls in both meta-analyses and narrative relationships. Notably, the evidence for physical activity as a protective factor against falls was insufficient, underscoring the complex relationships between physical activity and falls.^3^ In contrast to the overall results for mobility, the predictive accuracy for any of the specific balance and physical function tests was low (AUCs < 0·70) for any falls. These results highlight that while mobility-related measures are undoubtedly important risk factors for falls, multifactorial screening tools are likely necessary to accurately classify future fall risk.

Other risk factors that were frequently significant (i.e., ≥ 60% of reported relationships) included cognition, medications, frailty, and chronic conditions. Previous research on cognition in both community-dwelling and institutional settings aligns with our findings that the cognitive function being examined is an important consideration for determining fall risk, with executive function and processing speed most predictive of falls.^16^ The World Fall Guidelines advocate for executive function assessments as part of fall risk screening and, based on our findings, an assessment of processing speed may also be warranted. However, functions such as memory, language, and attention, may not predict falls. In terms of medications, the meta-analysis results and the narrative results align, suggesting that sedatives, psychotropics, and/or anti-depressants are likely fall risk factors (over for example, antihypertensives). Of note, the medication types had varying degrees of classifications, making it challenging to draw conclusions since some categories were embedded within each other. Notably, frailty and a number of the chronic conditions reported as significant predictors of falls in this review are often included in geriatric syndrome classifications.^17^ These conditions largely affect motor function, namely stroke, Parkinson’s disease, and arthritis. The impact of these conditions likely predisposes older adults to falling and these individuals should be considered high priority for fall prevention interventions.

Despite the breadth of predictors for falls, this umbrella review highlights the limited examination of psychosocial, sociodemographic, and environmental risk factors. While psychosocial factors showed a number of significant relationships (n= 12/13, 92% significant), the results focused predominantly on depression with few other predictors examined. Given the importance of psychosocial wellbeing for healthy ageing, and the modifiable nature of many psychosocial outcomes, additional attention should be given to this important topic. Similarly, findings on non-modifiable sociodemographic factors were scant. Previous research suggests both age and gender predict fall outcomes, while ethnicity, income, and education may limit access to fall prevention strategies.^18^ Further, these characteristics have been shown to increase risk for falls in institutionally dwelling populations^16, 18^ and are relatively easy to screen for, thus helping to triage those with greater fall prevention needs. Future research to identify the degree to which these factors predict fall risk in community-dwelling populations and how to best address them is warranted.

Throughout this umbrella review, risk factors were not classified by any particular system or framework. This was intentional since multiple systems already exist; classifying factors as intrinsic or extrinsic to the person,^19^ the 7 factor model suggested by Lord et al., ^5^ (socio-demographic; balance and mobility; sensory and neuromuscular; psychological; medical; medication; and environmental), and the 4 domains posited by the WHO’s Risk Factor Model for Falls^20^ (biological, socioeconomic, behavioral, and environmental). Additional fall-specific frameworks have been suggested but are not as common.^21^ Across these models, there is an emphasis on factors affecting one’s physical self, which perhaps has led to the high number of fall risk studies done to date in this area. Within our own results, physical factors were most commonly examined, yet factors such as depression, health status, and nutrition, while not frequently studied, show promise for future investigation. This umbrella review provides a rationale for examining risk factors from a deductive approach, as existing frameworks may have unintentionally missed relevant fall risk factors.

The heterogeneity of the literature in this area meant that we could only qualitatively summarize the results, as calculating risk magnitudes between studies would be inappropriate. There are several limitations with this methodology. First, the quality of primary studies was not assessed in this umbrella review, and so we cannot speak to the quality of the evidence, only the risk of bias in the synthesis. Next, the lack of reporting of confounders makes it hard to draw definitive conclusions. For instance, some reviews only selected adjusted models when extracting data from primary studies and others only selected unadjusted models. For those that reported adjusted models, not all reported the confounders. Some reviews also used approximations of effect estimates, for example, using HRs and ORs to approximate RR’s, thus overstating the relationships. In examining the degree of crossover of primary studies, the overall CCA was low (1%). The comprehensive and inclusive nature of our review ensures that as many relationships as possible were included, providing the largest synthesis to date of predictors of falls in community-dwelling older adults.

## Conclusion

Overall, this is the most comprehensive synthesis of fall risk factors in community-dwelling older adults. Our results identified 29 risk factors, with the ten most common being balance, cognition, gait, chronic conditions, physical function, physical activity, medications, dual task ability, frailty, and strength. Among these, dual task ability, frailty, chronic conditions, physical function, and gait factors had the highest number of significant results for predicting falls. The current literature has sufficiently examined physical aspects of fall risk yet has overlooked psychosocial and sociodemographic factors that could be valuable targets for future research.

## Contributions

Stephanie Saunders: co-conceptualized the study; curated the data by developing and executing the search strategy, screening abstracts and full texts, and reviewing the final studies; conducted the analysis; drafted the original manuscripts and completed final edits

Cassandra D’Amore: curated the data by screening abstracts and full texts, and reviewing the final studies; supported the analysis; critically revised the manuscript

Quikui Hao: curated the data by screening abstracts and full texts, and reviewing the final studies; supported the analysis; critically revised the manuscript

Nabil Abd El-Moneim: supported the analysis; critically revised the manuscript

Julie Richardson: co-conceptualized the study; supported the analysis; critically revised the manuscript

Ayse Kuspinar: co-conceptualized the study; supported the analysis; critically revised the manuscript

Marla Beauchamp: co-conceptualized the study; supported the analysis; critically revised the manuscript; supervised the first author

## Data Sharing

The datasets used and/or analysed during the current study are available from the corresponding author on reasonable request.

## Supporting information

All supplementary files

## Data Availability

All data produced in the present study are available upon reasonable request to the authors

## Acknowledgements

We would like to thank Laura Banfield for their help executing the search strategy.

## Supplementary Material

Appendix 1 – Search Details & Any Falls Results

- Search Strategies
- Table S·1. Excluded Full Text Studies
- Table S1·2. Risk of Bias – ROBIS Assessment
- Table S1·3. Summary of All Relationships Predicting Any Fall Risk
- Table S1·4. Summary of Meta-Analysis Relationships Predicting Any Fall Risk
- Table S1·5. Summary of Single Relationships Predicting Any Fall
- Table S1·6. Predictive Validity of Tests for Any Falls

Appendix 2 – Recurrent Falls Results

- Table S2·1. Summary of Meta-Analysis Relationships Predicting Recurrent Falls
- Table S2·2. Description of Meta-Analysis Relationships Predicting Recurrent Falls
- Table S2·3. Summary of Single Relationships Predicting Recurrent Falls
- Table S2·4. Description of Single Relationships Predicting Recurrent Falls
- Table S2·5. Predictive Validity of Tests for Recurrent Falls

Appendix 3 – Injurious Falls Results

- Table S3·1. Summary of Meta-Analysis Relationships Predicting Injurious Falls
- Table S3·2. Description of Meta-Analysis Relationships Predicting Injurious Falls
- Table S3·3. Summary of Single Relationships Predicting Injurious Falls
- Table S3·4. Description of Single Relationships Predicting Injurious Falls

Appendix 4 – Reference List of Included Studies

