## Supplementary material for "Risk factors for falls in community-dwelling older adults: An umbrella review": All supplementary files

***Appendices***

### ***APPENDICES***

### ***Appendix 1 – Search Details & Any Falls Results***

#### Search Strategies

**MEDLINE**

| 1. | **exp Accidental Falls/** |
| --- | --- |
| **2.** | **fall*.mp.** |
| **3.** | **1 or 2** |
| **4.** | **exp Aged/** |
| **5.** | **((old* or aged or frail) adj2 (person* or adult* or people* or patient*)).mp.** |
| **6.** | **(elder* or geriatric*).mp.** |
| **7.** | **retiree*.mp.** |
| **8.** | **pensioner*.mp.** |
| **9.** | **(senior* adj2 citizen*).mp.** |
| **10.** | **(senior* adj2 adult*).mp.** |
| **11.** | **or/4-10** |
| **12.** | **exp systematic review/** |
| **13.** | **(systematic adj1 review*).mp.** |
| **14.** | **meta?analys*.mp.** |
| **15.** | **exp meta-analysis/** |
| **16.** | **(scoping adj1 review*).mp.** |
| **17.** | **12 or 13 or 14 or 15 or 16** |
| **18.** | **3 and 11 and 17** |

**EMBASE**

| **1.** | **exp falling/** |
| --- | --- |
| **2.** | **exp fall risk assessment/ or fall*.mp.** |
| **3.** | **1 or 2** |
| **4.** | **exp aged/** |
| **5.** | **aging/ or cognitive aging/ or healthy aging/** |
| **6.** | **((old* or aged or frail) adj2 (person* or adult* or people* or patient*)).mp.** |
| **7.** | **(elder* or geriatric*).mp.** |
| **8.** | **retiree*.mp.** |
| **9.** | **pensioner*.mp. or exp pensioner/** |
| **10.** | **senior*.mp.** |
| **11.** | **4 or 5 or 6 or 7 or 8 or 9 or 10** |
| **12.** | **exp "systematic review"/** |
| **13.** | **(systematic adj1 review*).mp. [mp=title, abstract, heading word, drug trade name, original title, device manufacturer, drug manufacturer, device trade name, keyword, floating subheading word, candidate term word]** |
| **14.** | **exp meta analysis/** |
| **15.** | **meta?analys*.mp.** |
| **16.** | **(scoping adj1 review*).mp.** |
| **17.** | **12 or 13 or 14 or 15 or 16** |
| **18.** | **3 and 11 and 17** |

**CINAHL**

**(((MH "Aged+") OR (MH "Rehabilitation, Geriatric")) OR ((MH "Frail Elderly") OR old* OR frail OR (old* N2 adult*) OR (old* N2 person*) OR (old* N2 people*) OR (old* N2 patient*) OR elder* OR geriatric* OR retiree* OR pensioner* OR senior*)) AND ((MH "Accidental Falls") OR fall*) AND ((MH "Systematic Review") OR (MH "Scoping Review") OR (systematic N1 review*) OR (scoping review*) OR meta?analys*)**

**Ageline**

**((old* OR frail OR (old* N2 adult*) OR (old* N2 person*) OR (old* N2 people*) OR (old* N2 patient*) OR elder* OR geriatric* OR retiree* OR pensioner* OR senior*) OR (DE "60 " OR DE "65 " OR DE "70 " OR DE "75 " OR DE "80 " OR DE "85 " OR DE "90 " OR DE "95 " OR DE "Centenarians" OR DE "Old Old" OR DE "Young Old")) AND (((DE "Falls") OR fall*)) AND ((DE "Literature Reviews" OR DE "Meta Analyses") OR ((systematic N1 review*) OR ("scoping review*") OR meta?analys*))**

**COCHRANE**

1. **(Fall*):ti,ab,kw**
2. **(frail OR (old* NEAR /2 person*) OR (old* NEAR /2 adult*) OR (old* NEAR/2 patient*) OR eld* OR geriatric OR retiree* OR pensioner* OR senior):ti,ab,kw**
3. **(((systematic NEAR /2 review*) OR (meta analys*))):ti,ab,kw**
4. **MeSH descriptor: [Accidental Falls] explode all trees**
5. **MeSH descriptor: [Aged] in all MeSH products**
6. **#1 OR #4**
7. **#2 OR #5**
8. **#6 AND #7 AND #3**

**Web of Science**

**#1 TS=(Fall*)**

**#2 TS=(frail OR (old* NEAR /2 person*) OR (old* NEAR /2 adult*) OR (old* NEAR/2 patient*) OR eld* OR geriatric OR retiree* OR pensioner* OR senior)**

**#3 TS=(((systematic NEAR /2 review*) OR (meta?analys*)))**

**#4 #3 AND #2 AND #1**

#### Table S1.1 Excluded Full Text Studies

|  | **Study** | **Exclusion Reason** |
| --- | --- | --- |
| 1 | 廖英; 高静; 包新茹; 余静雅; 肖青青; 柏丁兮; 赵霞 血管紧张素转化酶抑制剂与老年人跌倒 风险关系的Meta 分析. Chinese Nursing Research,2019;33(14):2381-2386 | Samples not community dwelling |
| 2 | Abdelbasset, W. K.; Nambi, G.; Elsayed, S. H.; Osailan, A. M.; Eid, M. M. Falls and potential therapeutic interventions among elderly and older adult patients with cancer: a systematic review African Health Sciences,2021;21(4):1776-1783 | Review design |
| 3 | Abdelhafiz, Ahmed H.; Austin, Christopher A. Visual factors should be assessed in older people presenting with falls or hip fracture Age and Ageing,2003;32(1):26-30 | Review design |
| 4 | Abellan van Kan, G; Rolland, Y; Andrieu, S; BaueRandom, J; Beauchet, O; Bonnefoy, M; Cesari, M; Donini, L M; Gillette Guyonnet, S; Inzitari, M; Nourhashemi, F; OndeRandom, G; Ritz, P; Salva, A; VisseRandom, M; Vellas, B Gait speed at usual pace as a predictor of adverse outcomes in community-dwelling older people an International Academy on Nutrition and Aging (IANA) Task Force. The journal of nutrition, health & aging,2009;13(10):881-9 | Age not mean of 60 years + |
| 5 | Aboutorabi, Atefeh; Bahramizadeh, Mahmood; ArazpouRandom, Mokhtar; Fadayevatan, Reza; Farahmand, Farzam; Curran, Sarah; Hutchins, Stephen W A systematic review of the effect of foot orthoses and shoe characteristics on balance in healthy older subjects. Prosthetics and orthotics international,2016;40(2):170-81 | Review purpose not looking at falls |
| 6 | Abrahamsen, Bjarke; Hansen, Rikke N; Rossing, Charlotte For which patient subgroups are there positive outcomes from a medication review? A systematic review. Pharmacy practice,2020;18(4):1976 | Review purpose not looking at falls |
| 7 | Afifi, M.; Al-Hussein, M.; Bouferguene, A. Geriatric bathroom design to minimize risk of falling for older adults-a systematic review European Geriatric Medicine,2015;6(6):598-603 | Review purpose not looking at falls |
| 8 | Al-Musawe, Labib; Martins, Ana Paula; Raposo, Joao Filipe; Torre, Carla The association between polypharmacy and adverse health consequences in elderly type 2 diabetes mellitus patients; a systematic review and meta-analysis. Diabetes research and clinical practice,2019;155(ebi, 8508335):107804 | Review purpose not looking at falls |
| 9 | Alarcon, Teresa; Gonzalez-Montalvo, Juan Ignacio; Otero Puime, Angel [Assessing patients with fear of falling. Does the method use change the results? A systematic review]. Atencion primaria,2009;41(5):262-8 | Language |
| 10 | Albasri, Ali; Hattle, Miriam; Koshiaris, Constantinos; Dunnigan, Anna; Paxton, Ben; Fox, Sarah Emma; Smith, Margaret; ArcheRandom, Lucinda; Levis, Brooke; Payne, Rupert A.; Riley, Richard D.; Roberts, Nia; Snell, Kym I. E.; Lay-Flurrie, Sarah; Usher-Smith, Juliet; Stevens, Richard; Hobbs, F. D. Richard; McManus, Richard J.; Sheppard, James P.; Stratify investigators Association between antihypertensive treatment and adverse events: systematic review and meta-analysis BMJ (Clinical research ed.),2021;372():n189 | Other |
| 11 | Alenazi, A.; Alqahtani, B.; HooveRandom, J.; Alshehri, M. Alarming High Prevalence of Falls Among Older Adults in The Arabian Gulf Cooperation Council Countries Archives of Physical Medicine and Rehabilitation,2019;100(10):e148-e149 | Study designs not prospective |
| 12 | Algase, Donna L. What's new about wandering behaviour? An assessment of recent studies International Journal of Older People Nursing,2006;1(4):226-234 | Review purpose not looking at falls |
| 13 | Allen, Jennifer; Koziak, Adriana; Buddingh, Sarah; Liang, Jieyun; Buckingham, Jeanette; Beaupre, Lauren A Rehabilitation in patients with dementia following hip fracture: a systematic review. Physiotherapy Canada. Physiotherapie Canada,2012;64(2):190-201 | Review purpose not looking at falls |
| 14 | Allen, Natalie E; Schwarzel, Allison K; Canning, Colleen G Recurrent falls in Parkinson‚Äôs disease: a systematic review Parkinson‚Äôs disease,2013;2013(): | Age not mean of 60 years + |
| 15 | Allet, L.; Armand, S.; Golay, A.; Monnin, D.; de Bie, R. A.; de Bruin, E. D. Gait characteristics of diabetic patients: a systematic review Diabetes-Metabolism Research and Reviews,2008;24(3):173-191 | Age not mean of 60 years + |
| 16 | Alqahtani, Bader A; Alshehri, Mohammed M; HooveRandom, Jeffrey C; Alenazi, Aqeel M Prevalence of falls among older adults in the Gulf Cooperation Council countries: A systematic review and meta-analysis. Archives of gerontology and geriatrics,2019;83(8214379, 7ax):169-174 | Study designs not prospective |
| 17 | Ambrose, Anne Felicia; Paul, Geet; Hausdorff, Jeffrey M Risk factors for falls among older adults: a review of the literature Maturitas,2013;75(1):51-61 | Review design |
| 18 | American Geriatrics Society Workgroup on Vitamin D Supplementation for Older Adults Recommendations abstracted from the American Geriatrics Society Consensus Statement on vitamin D for Prevention of Falls and Their Consequences. Journal of the American Geriatrics Society,2014;62(1):147-52 | Review design |
| 19 | Amirpourabasi, Arezoo; Lamb, Sallie E.; Chow, Jia Yi; Williams, Genevieve K. R. Nonlinear Dynamic Measures of Walking in Healthy Older Adults: A Systematic Scoping Review Sensors (Basel, Switzerland),2022;22(12): | Review purpose not looking at falls |
| 20 | Angelousi, Anna; Girerd, Nicolas; Benetos, Athanase; Frimat, Luc; GautieRandom, Sylvie; Weryha, Georges; Boivin, Jean-Marc Association between orthostatic hypotension and cardiovascular risk, cerebrovascular risk, cognitive decline and falls as well as overall mortality: a systematic review and meta-analysis. Journal of hypertension,2014;32(8):1562-1571 | Samples not community dwelling |
| 21 | AnnweileRandom, C; Beauchet, O Questioning vitamin D status of elderly fallers and nonfallers: a meta-analysis to address a 'forgotten step'. Journal of internal medicine,2015;277(1):16-44 | Review purpose not looking at falls |
| 22 | AnnweileRandom, C; Schott, A M; Berrut, G; Fantino, B; Beauchet, O Vitamin D-related changes in physical performance: a systematic review. The journal of nutrition, health & aging,2009;13(10):893-8 | Review purpose not looking at falls |
| 23 | Avin, Keith G.; Hanke, Timothy A.; Kirk-Sanche, Neva; McDonough, Christine M.; Shubert, Tiffany E.; Hardage, Jason; Hartley, Greg Management of Falls in Community- Dwelling Older Adults: Clinical Guidance Statement From the Academy of Geriatric Physical Therapy of the American Physical Therapy Association. Physical Therapy,2015;95(6):815-834 | Review design |
| 24 | Bai, Xue; Han, Bing; Zhang, Man; Liu, Jinfeng; Cui, Yi; Jiang, Hong The association between diuretics and falls in older adults: A systematic review and meta-analysis Geriatric nursing (New York, N.Y.),2023;52():106-114 | Samples not community dwelling |
| 25 | BalzeRandom, Katrin; BremeRandom, Martina; Schramm, Susanne; Luhmann, Dagmar; Raspe, Heiner Falls prevention for the elderly. GMS health technology assessment,2012;8(101276045):Doc01 | Review purpose not looking at falls |
| 26 | Banda, K. J.; Chu, H.; Chen, R.; Kang, X. L.; Jen, H. J.; Liu, D.; Shen Hsiao, S. T.; Chou, K. R. Prevalence of Oropharyngeal Dysphagia and Risk of Pneumonia, Malnutrition, and Mortality in Adults Aged 60 Years and Older: A Meta-Analysis Gerontology,2022;68(8):841-853 | Review purpose not looking at falls |
| 27 | Barrett, R S; Mills, P M; Begg, R K A systematic review of the effect of ageing and falls history on minimum foot clearance characteristics during level walking. Gait & posture,2010;32(4):429-35 | Review purpose not looking at falls |
| 28 | Bauman, Adrian; Merom, Dafna; Bull, Fiona C.; BuchneRandom, David M.; Singh, Maria A. Fiatarone Updating the Evidence for Physical Activity: Summative Reviews of the Epidemiological Evidence, Prevalence, and Interventions to Promote ‚ÄúActive Aging‚Äù. Gerontologist,2016;56():S268-S280 | Review design |
| 29 | Bayot, Madli; Dujardin, Kathy; Dissaux, Lucile; Tard, Celine; Defebvre, Luc; Bonnet, Cedrick T; Allart, Etienne; Allali, Gilles; Delval, Arnaud Can dual-task paradigms predict Falls better than single task? - A systematic literature review. Neurophysiologie clinique = Clinical neurophysiology,2020;50(6):401-440 | Study designs not prospective |
| 30 | Beauchet, O; Fantino, B; Allali, G; MuiRandom, S W; Montero-Odasso, M; AnnweileRandom, C Timed Up and Go test and risk of falls in older adults: a systematic review. The journal of nutrition, health & aging,2011;15(10):933-8 | Samples not community dwelling |
| 31 | Beauchet, Olivier; Dubost, V; Revel Delhom, C; Berrut, G; Belmin, J; French Society of Geriatrics and Gerontology How to manage recurrent falls in clinical practice: guidelines of the French Society of Geriatrics and Gerontology. The journal of nutrition, health & aging,2011;15(1):79-84 | Review purpose not looking at falls |
| 32 | Beaudart, Charlotte; Zaaria, Myriam; Pasleau, Francoise; ReginsteRandom, Jean-Yves; Bruyere, Olivier Health Outcomes of Sarcopenia: A Systematic Review and Meta-Analysis. PloS one,2017;12(1):e0169548 | Review purpose not looking at falls |
| 33 | Beauplet, B.; Loggia, G.; Lequesne, J.; SolemLaviec, H.; Le Bon, P.; Beuscart, J.; Rambeau, A.; Girault, G. A systematic review on fall prevention in older patients with cancer and cancer survivors Supportive Care in CanceRandom,2021;29(SUPPL 1):S74 | Other |
| 34 | Beghe, C.; Wilson, A.; ErshleRandom, W. B. Prevalence and outcomes of anemia in geriatrics: A systematic review of the literature American Journal of Medicine,2004;116():45361 | Review purpose not looking at falls |
| 35 | Beghe, Claudia; Wilson, Alisa; ErshleRandom, William B Prevalence and outcomes of anemia in geriatrics: a systematic review of the literature. The American journal of medicine,2004;116 Suppl 7A(0267200, 3ju):3S-10S | Review purpose not looking at falls |
| 36 | Binotto, Maria Angelica; Lenardt, Maria Helena; Rodriguez-Martinez, Maria Del Carmen Physical frailty and gait speed in community elderly: a systematic review. Revista da Escola de Enfermagem da U S P,2018;52(rss, 0242726):e03392 | Language |
| 37 | Bird, Marie-Louise; Cheney, Michael J; Williams, Andrew D Accidental Fall Rates in Community-Dwelling Adults Compared to Cancer Survivors During and Post-Treatment: A Systematic Review With Meta-Analysis. Oncology nursing forum,2016;43(2):E64-72 | Age not mean of 60 years + |
| 38 | Biswas, I.; Adebusoye, B.; Chattopadhyay, K. Risk factors for falls among older adults in India: A systematic review and meta-analysis Health Sci Rep,2022;5(4):e637 | Samples not community dwelling |
| 39 | Blain, H.; Rolland, Y.; Beauchet, O.; AnnweileRandom, C.; Benhamou, C. L.; Benetos, A.; Berrut, G.; Audran, M.; Bendavid, S.; Bousson, V.; Briot, K.; BrazieRandom, M.; Breuil, V.; Chapuis, L.; Chapurlat, R.; Cohen-Solal, M.; Cortet, B.; Dargent, P.; Fardellone, P.; Feron, J. M.; Gauvain, J. B.; Guggenbuhl, P.; Hanon, O.; Laroche, M.; Kolta, S.; Lespessailles, E.; Letombe, B.; Mallet, E.; Marcelli, C.; Orcel, P.; Puisieux, F.; Seret, P.; Souberbielle, J. C.; SutteRandom, B.; Tremollieres, F.; Weryha, G.; Roux, C.; Thomas, T.; Grp, Rech; Information, Osteoporose Usefulness of bone density measurement in fallers Joint Bone Spine,2014;81(5):403-408 | Review design |
| 40 | Bloch, F; Thibaud, M; Dugue, B; Breque, C; Rigaud, A S; Kemoun, G Episodes of falling among elderly people: a systematic review and meta-analysis of social and demographic pre-disposing characteristics. Clinics (Sao Paulo, Brazil),2010;65(9):895-903 | Samples not community dwelling |
| 41 | Bloch, F.; Thibaud, M.; Tournoux, C.; Rigaud, A. S.; Kemoun, G. A meta-analysis-based study on risk factors for falls in the medical histories of elderly subjects: The importance of neurological disorders Parkinsonism and Related Disorders,2010;16(SUPPL 1):S19 | Other |
| 42 | Bloch, Frederic; Thibaud, Marie; Dugue, Benoit; Breque, Cyril; Rigaud, Anne-Sophie; Kemoun, Gilles Laxatives as a risk factor for iatrogenic falls in elderly subjects: myth or reality?. Drugs & aging,2010;27(11):895-901 | Study designs not prospective |
| 43 | Bloch, Frederic; Thibaud, Marie; Dugue, Benoit; Breque, Cyril; Rigaud, Anne-Sophie; Kemoun, Gilles Psychotropic drugs and falls in the elderly people: updated literature review and meta-analysis. Journal of aging and health,2011;23(2):329-46 | Samples not community dwelling |
| 44 | Bloch, Frederic; Thibaud, Marie; Tournoux-Facon, Caroline; Breque, Cyril; Rigaud, Anne-Sophie; Dugue, Benoit; Kemoun, Gilles Estimation of the risk factors for falls in the elderly: can meta-analysis provide a valid answer?. Geriatrics & gerontology international,2013;13(2):250-63 | Samples not community dwelling |
| 45 | Blum, Lisa; Korner-Bitensky, Nicol Usefulness of the Berg Balance Scale in stroke rehabilitation: a systematic review. Physical therapy,2008;88(5):559-66 | Review purpose not looking at falls |
| 46 | Boehm, Jackie; Franklin, Richard C.; King, Jemma C. Falls in rural and remote community dwelling older adults: A review of the literature. Australian Journal of Rural Health,2014;22(4):146-155 | Review purpose not looking at falls |
| 47 | Bouaziz, Walid; Vogel, Thomas; Schmitt, Elise; Kaltenbach, Georges; Geny, Bernard; Lang, Pierre Olivier [Health benefits of aerobic training programs in adults aged 70 or over: A systematic review]. Presse medicale (Paris, France : 1983),2017;46(9):794-807 | Review purpose not looking at falls |
| 48 | Bourke, R.; Doody, P.; Perez, S.; Moloney, D.; Lipsitz, L.; Kenny, R. A. Cardiovascular abnormalities and falls among older adults: A systematic review for the task force on global guidelines for falls in older adults European Geriatric Medicine,2022;13(Supplement 1):S28-S29 | Other |
| 49 | Bowen, L.; Griffioen, M. A Scoping Review of Falls and Physical Activity among Community-Dwelling Older Adults with Pain Archives of Physical Medicine and Rehabilitation,2019;100(12):e213 | Other |
| 50 | Brenton-Rule, Angela; Dalbeth, Nicola; Bassett, Sandra; Menz, Hylton B; Rome, Keith The incidence and risk factors for falls in adults with rheumatoid arthritis: a systematic review. Seminars in arthritis and rheumatism,2015;44(4):389-98 | Age not mean of 60 years + |
| 51 | Buck, Jackie; Hill, Julia Fromings; Martin, Alison; Springate, Cassandra; Ghosh, Bikramaditya; Ashton, Rachel; Lee, Gerry; Orlowski, Andrzei Reasons for discontinuing oral anticoagulation therapy for atrial fibrillation: a systematic review Age and Ageing,2021;50(4):1108-1117 | Review purpose not looking at falls |
| 52 | Butera, K. A.; Roff, S. R.; Buford, T. W.; Cruz-Almeida, Y. The impact of multisite pain on functional outcomes in older adults: Biopsychosocial considerations Journal of Pain Research,2019;12():1115-1125 | Review design |
| 53 | Butt, D. A.; Harvey, P. J. Benefits and risks of antihypertensive medications in the elderly. Journal of Internal Medicine,2015;278(6):599-626 | Review purpose not looking at falls |
| 54 | Cao Wenzhu; Huang Youyi; Xi Shuxin Meta - analysis of risk factors for fall in Chinese elderly. Chinese Nursing Research,2018;32(20):3222-3228 | Samples not community dwelling |
| 55 | Capiau, A.; Huys, L.; van Poelgeest, E.; van der Velde, N.; Petrovic, M.; Somers, A. Therapeutic dilemmas with benzodiazepines and Z-drugs: insomnia and anxiety disorders versus increased fall risk: a clinical review European Geriatric Medicine,2022;(): | Review design |
| 56 | Cardwell, Karen; Hughes, Carmel M; Ryan, Cristin The Association Between Anticholinergic Medication Burden and Health Related Outcomes in the 'Oldest Old': A Systematic Review of the Literature. Drugs & aging,2015;32(10):835-48 | Samples not community dwelling |
| 57 | Carlson, John E.; OstiRandom, Glenn V.; Black, Sandra A.; Markides, Kyriakos S.; Rudkin, Laura; Goodwin, James S. Disability in older adults 2: physical activity as prevention Behavioral Medicine,1999;24(4):157-168 | Review design |
| 58 | CarpenteRandom, Christopher R; Avidan, Michael S; Wildes, Tanya; Stark, Susan; FowleRandom, Susan A; Lo, Alexander X Predicting geriatric falls following an episode of emergency department care: a systematic review. Academic emergency medicine : official journal of the Society for Academic Emergency Medicine,2014;21(10):1069-82 | Other |
| 59 | CarpenteRandom, Christopher R.; Avidan, Michael S.; Wildes, Tanya; Stark, Susan; FowleRandom, Susan A.; Lo, Alexander X.; Gerson, Lowell Predicting Geriatric Falls Following an Episode of Emergency Department Care: A Systematic Review La Predicci√≥n de Ca√≠das Geri√°tricas tras un Episodio en el Servicio de Urgencias: Una Revisi√≥n Sistem√°tica. Academic Emergency Medicine,2014;21(10):1069-1082 | Samples not community dwelling |
| 60 | Cavanaugh, Ellen J; Richardson, Jenna; McCallum, Christine A; Wilhelm, Mark The Predictive Validity of Physical Performance Measures in Determining Markers of Preclinical Disability in Community-Dwelling Middle-Aged and Older Adults: A Systematic Review. Physical therapy,2018;98(12):1010-1021 | Review purpose not looking at falls |
| 61 | Chan, D. K. Y.; Gibian, T. Medications and falls in the elderly Australian Journal on Ageing,1994;13(1):22-26 | Review design |
| 62 | ChandleRandom, Julie McClure Understanding the relationship between strength and mobility in frail older persons: a review of the literature Topics in Geriatric Rehabilitation,1996;11(3):20-37 | Review design |
| 63 | Chang, Chee-Tao; Ang, Ju-Ying; Islam, Md Asiful; Chan, Huan-Keat; Cheah, Wee-Kooi; Gan, Siew Hua Prevalence of Drug-Related Problems and Complementary and Alternative Medicine Use in Malaysia: A Systematic Review and Meta-Analysis of 37,249 Older Adults Pharmaceuticals (Basel, Switzerland),2021;14(3): | Study designs not prospective |
| 64 | Chen-Ju, Fu; Wen-Chien, Chen; Meng-Ling, Lu; Chi-Chien, Niu; Yi-Hsuan, Lee; Chih-Hsiu, Cheng Equipment-Free Fall-Risk Assessments for the Functionally Independent Elderly: A Systematic Review and Meta-Analysis International Journal of Gerontology,2021;15(4):301-308 | Samples not community dwelling |
| 65 | Chen, B.; Wang, M.; He, Q.; Wang, Y.; Lai, X.; Chen, H.; Li, M. Impact of frailty, mild cognitive impairment and cognitive frailty on adverse health outcomes among community-dwelling older adults: A systematic review and meta-analysis Frontiers in Medicine,2022;9():1009794 | Review purpose not looking at falls |
| 66 | Chen, Manting; Wang, Hailiang; Yu, Lisha; Yeung, Eric Hiu Kwong; Luo, Jiajia; Tsui, Kwok-Leung; Zhao, Yang A Systematic Review of Wearable Sensor-Based Technologies for Fall Risk Assessment in Older Adults Sensors (Basel, Switzerland),2022;22(18): | Study designs not prospective |
| 67 | Chen, Serena Kuangyi; VoaklandeRandom, Don; Perry, Danielle; Jones, C Allyson Falls and fear of falling in older adults with total joint arthroplasty: a scoping review. BMC musculoskeletal disorders,2019;20(1):599 | Samples not community dwelling |
| 68 | Chen, Y.; Zhu, L. L.; Zhou, Q. Effects of drug pharmacokinetic/pharmacodynamic properties, characteristics of medication use, and relevant pharmacological interventions on fall risk in elderly patients Therapeutics and Clinical Risk Management,2014;10(1):437-448 | Review design |
| 69 | Chiarelli, Pauline E; Mackenzie, Lynette A; Osmotherly, Peter G Urinary incontinence is associated with an increase in falls: a systematic review. The Australian journal of physiotherapy,2009;55(2):89-95 | Study designs not prospective |
| 70 | Cho, H; Myung, J; Suh, H S; Kang, H-Y Antihistamine use and the risk of injurious falls or fracture in elderly patients: a systematic review and meta-analysis. Osteoporosis international : a journal established as result of cooperation between the European Foundation for Osteoporosis and the National Osteoporosis Foundation of the USA,2018;29(10):2163-2170 | Samples not community dwelling |
| 71 | Chou, Roger; Dana, Tracy; Bougatsos, Christina ,2009;(): | Review design |
| 72 | Chou, Roger; Dana, Tracy; Bougatsos, Christina ,2009;(): | Review design |
| 73 | Chow, K. P.; Fong, D. Y. T.; Wang, M. P.; Wong, J. Y. H.; Chau, P. H. Meteorological factors to fall: a systematic review International Journal of Biometeorology,2018;62(12):2073-2088 | Samples not community dwelling |
| 74 | Chowdhury, R.; Peel, N. M.; Krosch, M.; Hubbard, R. E. Frailty and chronic kidney disease: A systematic review Archives of Gerontology and Geriatrics,2017;68():135-142 | Review purpose not looking at falls |
| 75 | Chu, Yu-Hsiu; Tang, Pei-Fang; Peng, Ya-Chi; Chen, Hui-Ya Meta-analysis of type and complexity of a secondary task during walking on the prediction of elderly falls. Geriatrics & gerontology international,2013;13(2):289-97 | Samples not community dwelling |
| 76 | Church, Sophie; Rogers, Emily; Rockwood, Kenneth; Theou, Olga A scoping review of the Clinical Frailty Scale. BMC geriatrics,2020;20(1):393 | Review purpose not looking at falls |
| 77 | Ciorba, Andrea DIZZINESS AND THE RISK OF FALLING IN THE ELDERLY: A LITERATURE REVIEW. Journal of Hearing Science,2015;5(1):45548 | Review design |
| 78 | Clark, E.; Podschun, L.; Church, K.; Fleagle, A.; Hull, P.; Ohree, S.; Springfield, M.; Wood, S. Use of accelerometers in determining risk of falls in individuals post-stroke: A systematic review Clinical Rehabilitation,2023;():2692155231168300 | Age not mean of 60 years + |
| 79 | Coote, Susan; CombeRandom, Laura; Quinn, Gillian; Santoyo-Medina, Carme; Kalron, Alon; Gunn, Hilary Falls in People with Multiple Sclerosis: Risk Identification, Intervention, and Future Directions. International Journal of MS Care,2020;22(6):247-255 | Review design |
| 80 | Corona, Giovanni; Norello, Dario; Parenti, Gabriele; Sforza, Alessandra; Maggi, Mario; Peri, Alessandro Hyponatremia, falls and bone fractures: A systematic review and meta-analysis. Clinical endocrinology,2018;89(4):505-513 | Samples not community dwelling |
| 81 | Costa, Alice Gabrielle de Sousa; Oliveira, Ana Railka de Souza; de Sousa, Vanessa Emille Carvalho; de Araujo, Thelma Leite; Cardoso, Maria Vera L√∫cia Moreira Leit√£o; da Silva, Viviane Martins Instruments used in brazil for evaluation of physical mobility as predictor factor of falls in adults. Ciencia, Cuidado e Saude,2011;10(2):401-407 | Language |
| 82 | Coyne, K S; Wein, A; Nicholson, S; Kvasz, M; Chen, C-I; Milsom, I Comorbidities and personal burden of urgency urinary incontinence: a systematic review. International journal of clinical practice,2013;67(10):1015-33 | Review purpose not looking at falls |
| 83 | Crabtree, Thomas; Ogendo, Jael-Joy; Vinogradova, Yana; Gordon, Jason; Idris, Iskandar Intensive glycemic control and macrovasculaRandom, microvasculaRandom, hypoglycemia complications and mortality in older (age >=60years) or frail adults with type 2 diabetes: a systematic review and meta-analysis from randomized controlled trial and observation studies Expert review of endocrinology & metabolism,2022;17(3):255-267 | Samples not community dwelling |
| 84 | Crandall, Marie; Duncan, Thomas; Mallat, Ali; Greene, Wendy; Violano, Pina; Christmas, A Britton; Barraco, Robert Prevention of fall-related injuries in the elderly: An Eastern Association for the Surgery of Trauma practice management guideline. The journal of trauma and acute care surgery,2016;81(1):196-206 | Review design |
| 85 | Cranney, A.; WeileRandom, H. A.; O'Donnell, S.; Puil, L. Summary of evidence-based review on vitamin D efficacy and safety in relation to bone health American Journal of Clinical Nutrition,2008;88(2):513S-519S | Study designs not prospective |
| 86 | Cranney, Ann; Horsley, Tanya; O'Donnell, Siobhan; WeileRandom, Hope; Puil, Lorri; Ooi, Daylily; Atkinson, Stephanie; Ward, Leanne; MoheRandom, David; Hanley, David; Fang, Manchung; Yazdi, Fatemeh; Garritty, Chantelle; Sampson, Margaret; Barrowman, Nick; Tsertsvadze, Alex; Mamaladze, Vasil Effectiveness and safety of vitamin D in relation to bone health. Evidence report/technology assessment,2007;(158):1-235 | Review design |
| 87 | Creaby, M. W.; Cole, M. H. Gait characteristics and falls in Parkinson's disease: A systematic review and meta-analysis Parkinsonism & Related Disorders,2018;57():45299 | Samples not community dwelling |
| 88 | Cui, M.; Yu, K.; Li, C.; Li, R. Impact of sarcopenia on the risk of falls, osteoporosis, fractures, and all causes of death among elderly people: A Meta-analysis of prospective cohort studies Chinese Journal of Clinical Nutrition,2018;26(5):299-308 | Samples not community dwelling |
| 89 | Cui, M.; Yu, K.; Li, C.; Li, R. Relationship between sarcopenia and the incidence of fall in elderly-a systematic review of advanced nutritional and physical interventions studies Chinese Journal of Clinical Nutrition,2017;25(5):278-285 | Samples not community dwelling |
| 90 | Darowski, Adam; Whiting, Robert Cardiovascular medication and falls. Reviews in Clinical Gerontology,2011;21(2):170-179 | Review design |
| 91 | Davis, A.; Haines, T.; Williams, C. Do footwear styles cause falls or increase falls risk in healthy older adults? A systematic review Footwear Science,2019;11(1):13-23 | Samples not community dwelling |
| 92 | de Clercq, Hendrika; Naud√©, Alida; Bornman, Juan Factors included in adult fall risk assessment tools (FRATs): a systematic review Ageing & Society,2021;41(11):2558-2582 | Review purpose not looking at falls |
| 93 | de Rezende, Leandro Fornias Machado; Rey-Lopez, Juan Pablo; Matsudo, Victor Keihan Rodrigues; do Carmo Luiz, Olinda Sedentary behavior and health outcomes among older adults: a systematic review. BMC public health,2014;14(100968562):333 | Review purpose not looking at falls |
| 94 | de Vries, Max; Seppala, Lotta J; Daams, Joost G; van de Glind, Esther M M; Masud, Tahir; van der Velde, Nathalie; EUGMS Task and Finish Group on Fall-Risk-Increasing Drugs Fall-Risk-Increasing Drugs: A Systematic Review and Meta-Analysis: I. Cardiovascular Drugs. Journal of the American Medical Directors Association,2018;19(4):371.e1-371.e9 | Samples not community dwelling |
| 95 | Desapriya, Ediriweera; Subzwari, Sayed; Scime-Beltrano, Giulia; Samayawardhena, Lionel A; Pike, Ian Vision improvement and reduction in falls after expedited cataract surgery Systematic review and metaanalysis. Journal of cataract and refractive surgery,2010;36(1):45548 | Samples not community dwelling |
| 96 | Dhital, A; Pey, T; Stanford, Miles R Visual loss and falls: a review Eye,2010;24(9):1437-1446 | Samples not community dwelling |
| 97 | di Laura Frattura, G.; Filardo, G.; Giunchi, D.; Fusco, A.; Zaffagnini, S.; Candrian, C. Risk of falls in patients with knee osteoarthritis undergoing total knee arthroplasty: A systematic review and best evidence synthesis J Orthop,2018;15(3):903-908 | Samples not community dwelling |
| 98 | di Laura Frattura, Giorgio; Filardo, Giuseppe; Giunchi, Dario; Fusco, Augusto; Zaffagnini, Stefano; Candrian, Christian Risk of falls in patients with knee osteoarthritis undergoing total knee arthroplasty: A systematic review and best evidence synthesis. Journal of orthopaedics,2018;15(3):903-908 | Samples not community dwelling |
| 99 | Diaz-Gutierrez, M Jose; Martinez-Cengotitabengoa, Monica; Saez de Adana, Estibaliz; Cano, Ana Isabel; Martinez-Cengotitabengoa, Maria Teresa; Besga, Ariadna; Segarra, Rafael; Gonzalez-Pinto, Ana Relationship between the use of benzodiazepines and falls in older adults: A systematic review. Maturitas,2017;101(mwn, 7807333):17-22 | Samples not community dwelling |
| 100 | Dolatabadi, Elham; Van Ooteghem, Karen; Taati, Babak; Iaboni, Andrea Quantitative Mobility Assessment for Fall Risk Prediction in Dementia: A Systematic Review. Dementia and geriatric cognitive disorders,2018;45(45418):353-367 | Samples not community dwelling |
| 101 | Dubbeldam, Rosemary; Lee, Yu Yuan; Pennone, Juliana; Mochizuki, Luis; Le Mouel, Charlotte Systematic review of candidate prognostic factors for falling in older adults identified from motion analysis of challenging walking tasks European review of aging and physical activity : official journal of the European Group for Research into Elderly and Physical Activity,2023;20(1):2 | Age not mean of 60 years + |
| 102 | Duong, Mai H.; Gnjidic, Danijela; McLachlan, Andrew J.; Sakiris, Marissa A.; Goyal, Parag; HilmeRandom, Sarah N. The Prevalence of Adverse Drug Reactions and Adverse Drug Events from Heart Failure Medications in Frail Older Adults: A Systematic Review Drugs & Aging,2022;39(8):631-643 | Review purpose not looking at falls |
| 103 | DyeRandom, Suzanne M.; Suen, Jenni; Williams, Helena; Inacio, Maria C.; Harvey, Gillian; RodeRandom, David; Wesselingh, Steve; Kellie, Andrew; Crotty, Maria; Caughey, Gillian E. Impact of relational continuity of primary care in aged care: a systematic review BMC Geriatrics,2022;22(1):45309 | Samples not community dwelling |
| 104 | Eades, C.; Haddow, L. Falls and falls-related fracture in HIV+ adults over 45 years: A systematic review HIV Medicine,2014;15(SUPPL. 3):74 | Other |
| 105 | Eagles, Debra; Yadav, Krishan; Perry, Jeffrey J; Sirois, Marie Josee; Emond, Marcel Mobility assessments of geriatric emergency department patients: A systematic review. CJEM,2018;20(3):353-361 | Samples not community dwelling |
| 106 | Edwards, Nancy; Dulai, Joshun; Rahman, Alvi A Scoping Review of Epidemiological, Ergonomic, and Longitudinal Cohort Studies Examining the Links between Stair and Bathroom Falls and the Built Environment. International journal of environmental research and public health,2019;16(9): | Samples not community dwelling |
| 107 | Ehrenpreis, E. D.; Doniparthi, M.; DeMent, M.; Pulido, C.; Bartl, M. B. Systematic Review of Anticholinergic Drugs Used in the Treatment of Gastrointestinal Conditions American Journal of Gastroenterology,2020;115(SUPPL):S245 | Other |
| 108 | El-Saifi, N; Moyle, W; Jones, C; Tuffaha, H Quetiapine safety in older adults: a systematic literature review. Journal of clinical pharmacy and therapeutics,2016;41(1):45491 | Review purpose not looking at falls |
| 109 | Elias Filho, Jose; Borel, Wyngrid Porfirio; Diz, Juliano Bergamaschine Mata; Barbosa, Alexandre Wesley Carvalho; Britto, Raquel Rodrigues; Felicio, Diogo Carvalho Prevalence of falls and associated factors in community-dwelling older Brazilians: a systematic review and meta-analysis. Cadernos de saude publica,2019;35(8):e00115718 | Study designs not prospective |
| 110 | Ellen, F.; Toby, E.; Chris, T.; Lisa, M.; David, H.; Rixt, Z. G. A.; Lim, M. L.; Manuel, M. O.; Kim, D.; Ryota, S. Global guidelines for falls in older adults: Working group 12: Fear of falling European Geriatric Medicine,2022;13(Supplement 1):S239 | Other |
| 111 | Ergin, Emine; Akin, Belgin; Kocoglu-TanyeRandom, Deniz Effect of Home Visits by Nurses on the Physical and Psychosocial Health of Older Adults: A Systematic Review and Meta-Analysis Iranian journal of public health,2022;51(4):733-745 | Study designs not prospective |
| 112 | Falck, Ryan S; Davis, Jennifer C; Best, John R; Crockett, Rachel A; Liu-Ambrose, Teresa Impact of exercise training on physical and cognitive function among older adults: a systematic review and meta-analysis Neurobiology of aging,2019;79():119-130 | Samples not community dwelling |
| 113 | Feldman, Fabio; Chaudhury, Habib Falls and the physical environment: a review and a new multifactorial falls-risk conceptual framework Canadian Journal of Occupational Therapy,2008;75(2):82-95 | Study designs not prospective |
| 114 | Fernandez-Arguelles, Esther Lopez; Rodriguez-Mansilla, Juan; Antunez, Luis Espejo; Garrido-Ardila, Elisa Maria; Munoz, Rafael Perez Effects of dancing on the risk of falling related factors of healthy older adults: a systematic review. Archives of gerontology and geriatrics,2015;60(1):45299 | Review purpose not looking at falls |
| 115 | Fhon, J. R. S.; Silva, A. R. F.; Lima, E. F. C.; Santos Neto, A. P. D.; Henao-Castano, A. M.; Fajardo-Ramos, E.; Puschel, V. A. A. Association between Sarcopenia, Falls, and Cognitive Impairment in Older People: A Systematic Review with Meta-Analysis International Journal of Environmental Research and Public Health,2023;20(5):4156 | Study designs not prospective |
| 116 | Fhon, Jack Roberto Silva; Rodrigues, Rosalina Aparecida Partezani; Neira, Wilmer Fuentes; Huayta, Violeta Magdalena Rojas; Robazzi, Maria Lucia do Carmo Cruz Fall and its association with the frailty syndrome in the elderly: systematic review with meta-analysis. Revista da Escola de Enfermagem da U S P,2016;50(6):1005-1013 | Study designs not prospective |
| 117 | Finco, M. G.; Sumien, Nathalie; Moudy, Sarah C. Clinical evaluation of fall risk in older adults who use lower-limb prostheses: A scoping review Journal of the American Geriatrics Society,2023;71(3):959-967 | Study designs not prospective |
| 118 | Fosnight, S. M.; Zafirau, W. J.; Hazelett, S. E. Vitamin D supplementation to prevent falls in the elderly: Evidence and practical considerations Pharmacotherapy,2008;28(2):225-234 | Review design |
| 119 | Fratangelo, L.; Nguyen, S.; D'Amelio, P. Hyponatremia and aging-related diseases: Key player or innocent bystander? European Geriatric Medicine,2022;13(Supplement 1):S234-S235 | Other |
| 120 | Fratangelo, Luigia; Nguyen, Sylvain; D'Amelio, Patrizia Hyponatremia and aging-related diseases: key player or innocent bystander? A systematic review Systematic reviews,2023;12(1):84 | Other |
| 121 | Fried, Terri R; O'Leary, John; Towle, Virginia; Goldstein, Mary K; Trentalange, Mark; Martin, Deanna K Health outcomes associated with polypharmacy in community-dwelling older adults: a systematic review. Journal of the American Geriatrics Society,2014;62(12):2261-72 | Review purpose not looking at falls |
| 122 | G R Neri, Silvia; S Oliveira, Juliana; B Dario, Amabile; M Lima, Ricardo; Tiedemann, Anne Does Obesity Increase the Risk and Severity of Falls in People Aged 60 Years and Older? A Systematic Review and Meta-analysis of Observational Studies. The journals of gerontology. Series A, Biological sciences and medical sciences,2020;75(5):952-960 | Samples not community dwelling |
| 123 | Gade, G. V.; Jorgensen, M. G.; Ryg, J.; Riis, J.; Thomsen, K.; Masud, T.; Andersen, S. Predicting falls in community-dwelling older adults: A systematic review of prognostic models BMJ Open,2021;11(5):e044170 | Review purpose not looking at falls |
| 124 | GallagheRandom, Celine; Nyfort-Hansen, Karin; Rowett, Debra; Wong, Christopher X; Middeldorp, Melissa E; Mahajan, Rajiv; Lau, Dennis H; Sanders, Prashanthan; Hendriks, Jeroen M Polypharmacy and health outcomes in atrial fibrillation: a systematic review and meta-analysis. Open heart,2020;7(1):e001257 | Review purpose not looking at falls |
| 125 | GallagheRandom, Celine; Nyfort-Hansen, Karin; Rowett, Debra; Wong, Christopher X; Middeldorp, Melissa E; Mahajan, Rajiv; Lau, Dennis H; Sanders, Prashanthan; Hendriks, Jeroen M Polypharmacy and health outcomes in atrial fibrillation: a systematic review and meta-analysis Open Heart,2020;7(1):e001257 | Review purpose not looking at falls |
| 126 | Gama, Zenewton Andre da Silva; Gomez-Conesa, Antonia [Risk factors for falls in the elderly: systematic review]. Revista de saude publica,2008;42(5):946-56 | Language |
| 127 | Gandham, A.; Mesinovic, J.; Jansons, P.; Zengin, A.; Bonham, M. P.; Ebeling, P. R.; Scott, D. Falls, fractures, and areal bone mineral density in older adults with sarcopenic obesity: A systematic review and meta-analysis Obesity Reviews,2021;22(5):e13187 | Age not mean of 60 years + |
| 128 | Gao, Harrison; Yous, Marie-Lee; Connelly, Denise; Hung, Lillian; Garnett, Anna; Hay, Melissa; Snobelen, Nancy Implementation and impacts of virtual team-based care planning for older persons in formal care settings: A scoping review Digital health,2023;9():20552076231151500 | Review purpose not looking at falls |
| 129 | Garrigan, Hannah; Hamati, Jacquelyn; Lalakia, Parth; Frasso, Rosemary; Salzman, Brooke; Hyman, Leslie Does age-related macular degeneration (AMD) treatment influence patient falls and mobility? A systematic review Ophthalmic epidemiology,2022;29(2):128-138 | Age not mean of 60 years + |
| 130 | Gavrila Laic, R. A.; Bogaert, L.; Vander Sloten, J.; Depreitere, B. Functional outcome, dependency and well-being after traumatic brain injury in the elderly population: A systematic review and meta-analysis Brain and Spine,2021;1():100849 | Review purpose not looking at falls |
| 131 | Gawronska, K.; Lorkowski, J. Falls as One of the Atypical Presentations of COVID-19 in Older Population Geriatric Orthopaedic Surgery and Rehabilitation,2021;12(): | Review design |
| 132 | Gebara, Marie Anne; Lipsey, Kim L; Karp, Jordan F; Nash, Maureen C; Iaboni, Andrea; Lenze, Eric J Cause or Effect? Selective Serotonin Reuptake Inhibitors and Falls in Older Adults: A Systematic Review. The American journal of geriatric psychiatry : official journal of the American Association for Geriatric Psychiatry,2015;23(10):1016-28 | Samples not community dwelling |
| 133 | Gerbasi, M. E.; NambiaRandom, S.; Reed, S.; Hennegan, K.; HadkeRandom, N.; Eldar-Lissai, A.; Cosentino, S. Essential tremor patients experience significant burden beyond tremor: A systematic literature review Frontiers in Neurology,2022;13():891446 | Age not mean of 60 years + |
| 134 | Gianni, C.; Prosperini, L.; JonsdottiRandom, J.; Cattaneo, D. A systematic review of factors associated with accidental falls in people with multiple sclerosis: a meta-analytic approach Clinical Rehabilitation,2014;28(7):704-716 | Study designs not prospective |
| 135 | Goto, N. A.; Weststrate, A. C. G.; Oosterlaan, F. M.; VerhaaRandom, M. C.; Willems, H. C.; Emmelot-Vonk, M. H.; HamakeRandom, M. E. The association between chronic kidney disease, falls, and fractures: a systematic review and meta-analysis. Osteoporosis International,2020;31(1):13-29 | Review purpose not looking at falls |
| 136 | Grabowska, Weronika; Burton, Wren; Kowalski, Matthew H.; Vining, Robert; Long, Cynthia R.; Lisi, Anthony; Hausdorff, Jeffrey M.; ManoRandom, Brad; Mu√±oz-Vergara, Dennis; Wayne, Peter M. A systematic review of chiropractic care for fall prevention: rationale, state of the evidence, and recommendations for future research BMC Musculoskeletal Disorders,2022;23(1):45322 | Age not mean of 60 years + |
| 137 | GranacheRandom, Urs; GollhofeRandom, Albert; Hortobagyi, Tibor; Kressig, Reto W; MuehlbaueRandom, Thomas The importance of trunk muscle strength for balance, functional performance, and fall prevention in seniors: a systematic review. Sports medicine (Auckland, N.Z.),2013;43(7):627-41 | Study designs not prospective |
| 138 | Graveson, Jack; BauermeisteRandom, Sarah; McKeown, Denis; Bunce, David Intraindividual Reaction Time Variability, Falls, and Gait in Old Age: A Systematic Review. The journals of gerontology. Series B, Psychological sciences and social sciences,2016;71(5):857-64 | Study designs not prospective |
| 139 | Gregg, Edward W.; Pereira, Mark A.; Caspersen, Carl J. Physical activity, falls, and fractures among older adults: a review of the epidemiologic evidence Journal of the American Geriatrics Society,2000;48(8):883-893 | Samples not community dwelling |
| 140 | Guelich, Marsha M. Prevention of falls in the elderly: a literature review Topics in Geriatric Rehabilitation,1999;15(1):15-25 | Review design |
| 141 | Gunn, H. J.; Newell, P.; Haas, B.; Marsden, J. F.; Freeman, J. A. Identification of Risk Factors for Falls in Multiple Sclerosis: A Systematic Review and Meta-Analysis Physical Therapy,2013;93(4):504-513 | Age not mean of 60 years + |
| 142 | Guo, X.; Pei, J.; Ma, Y.; Cui, Y.; Guo, J.; Wei, Y.; Han, L. Cognitive Frailty as a Predictor of Future Falls in Older Adults: A Systematic Review and Meta-analysis Journal of the American Medical Directors Association,2022;(): | Samples not community dwelling |
| 143 | Haasum, Ylva; Johnell, Kristina Use of antiepileptic drugs and risk of falls in old age: A systematic review. Epilepsy research,2017;138(ema, 8703089):98-104 | Study designs not prospective |
| 144 | Hacƒ±dursunoƒülu Erba≈ü, Dilay; √áƒ±naRandom, Fadime; Eti Aslan, Fatma Elderly patients and falls: a systematic review and meta-analysis Aging Clinical and Experimental Research,2021;33(11):2953-2966 | Samples not community dwelling |
| 145 | Hallford, David John; Nicholson, Geoff; Sanders, Kerrie; McCabe, Marita P The Association Between Anxiety and Falls: A Meta-Analysis. The journals of gerontology. Series B, Psychological sciences and social sciences,2017;72(5):729-741 | Study designs not prospective |
| 146 | HamacheRandom, D; Singh, N B; Van Dieen, J H; HelleRandom, M O; TayloRandom, W R Kinematic measures for assessing gait stability in elderly individuals: a systematic review. Journal of the Royal Society, Interface,2011;8(65):1682-98 | Study designs not prospective |
| 147 | Hanlon, Joseph T.; Cutson, Toni; Ruby, Christine M. Drug-related falls in the older adult Topics in Geriatric Rehabilitation,1996;11(3):38-54 | Samples not community dwelling |
| 148 | Harlein, Jurgen; Dassen, Theo; Halfens, Ruud J G; Heinze, Cornelia Fall risk factors in older people with dementia or cognitive impairment: a systematic review. Journal of advanced nursing,2009;65(5):922-33 | Samples not community dwelling |
| 149 | Hartikainen, Sirpa; L√∂nnroos, Eija; Louhivuori, Kirsti Medication as a Risk Factor for Falls: Systematic Review Journals of Gerontology: Series A: Biological Sciences and Medical Sciences,2007;62(10):1172-1181 | Samples not community dwelling |
| 150 | Hartikainen, Sirpa; Lonnroos, Eija; Louhivuori, Kirsti Medication as a risk factor for falls: critical systematic review. The journals of gerontology. Series A, Biological sciences and medical sciences,2007;62(10):1172-81 | Other |
| 151 | Hartikainen, Sirpa; Lonnroos, Eija; Louhivuori, Lirsti Medication as a risk factor for falls: critical systematic review Journals of Gerontology: Series A: Biological Sciences and Medical Sciences,2007;62A(10):1172-1181 | Samples not community dwelling |
| 152 | Hartog, Laura Caroline; Schrijnders, Dennis; Landman, G W D; GroenieRandom, Klaas; Kleefstra, Nanne; Bilo, Henk J G; van Hateren, Kornelis Johannes Jongers Is orthostatic hypotension related to falling? A meta-analysis of individual patient data of prospective observational studies. Age and ageing,2017;46(4):568-575 | Other |
| 153 | Harvey, N. C.; Oden, A.; Orwoll, E.; Kwok, T.; Karlsson, M. K.; Rosengren, B. E.; Ribom, E.; Cawthon, P. M.; Ensrud, K.; CoopeRandom, C.; Kanis, J. A.; Lorentzon, M.; Ohlsson, C.; Mellstrom, D.; Johansson, H.; McCloskey, E. V. Sarcopenia definitions as predictors of fracture risk independent of frax, falls andbmd in the osteoporotic fractures in men (MROS) study: A meta-analysis Osteoporosis International,2019;30(SUPPL 2):S176 | Other |
| 154 | Harvey, Nicholas C; Orwoll, Eric; Kwok, Timothy; Karlsson, Magnus K; Rosengren, Bj√∂rn E; Ribom, Eva; Cauley, Jane A; Cawthon, Peggy M; Ensrud, Kristine; Liu, Enwu Sarcopenia definitions as predictors of fracture risk independent of FRAX¬Æ, falls, and BMD in the osteoporotic fractures in men (MrOS) study: a meta‚Äêanalysis Journal of Bone and Mineral Research,2021;36(7):1235-1244 | Review design |
| 155 | Healey, Frances; OliveRandom, David; Milne, Alisoun; Connelly, James B. Effect of bedrails on falls and injury: a systematic review of clinical studies Age and Ageing,2008;37(4):368-378 | Samples not community dwelling |
| 156 | Hegeman, Judith; van den Bemt, Bart J F; Duysens, Jacques; van Limbeek, Jacques NSAIDs and the risk of accidental falls in the elderly: a systematic review. Drug safety,2009;32(6):489-98 | Study designs not prospective |
| 157 | Hoang, Ptsb; Mol, A.; Reijnierse, E. M.; Meskers, C. G. M.; MaieRandom, A. B. Orthostatic hypotension and falls in older adults: A systematic review and meta-analysis Australasian Journal on Ageing,2018;37():45-45 | Samples not community dwelling |
| 158 | Hodge, William; Horsley, Tanya; Albiani, David; Baryla, Julia; Belliveau, Michel; Buhrmann, Ralf; O'ConnoRandom, Michael; BlaiRandom, Jason; Lowcock, Elizabeth The consequences of waiting for cataract surgery: a systematic review. CMAJ : Canadian Medical Association journal = journal de l'Association medicale canadienne,2007;176(9):1285-90 | Review purpose not looking at falls |
| 159 | Hsu, C L; Nagamatsu, L S; Davis, J C; Liu-Ambrose, T Examining the relationship between specific cognitive processes and falls risk in older adults: a systematic review. Osteoporosis international : a journal established as result of cooperation between the European Foundation for Osteoporosis and the National Osteoporosis Foundation of the USA,2012;23(10):2409-24 | Study designs not prospective |
| 160 | Hsu, Hui‚ÄêFen; Chen, Kuei‚ÄêMin; Belcastro, Frank; Chen, Yih‚ÄêFung Polypharmacy and pattern of medication use in community‚Äêdwelling older adults: A systematic review Journal of Clinical Nursing (John Wiley & Sons, Inc.),2021;30(45481):918-928 | Review purpose not looking at falls |
| 161 | Hu, K.; Zhou, Q.; Jiang, Y.; Shang, Z.; Mei, F.; Gao, Q.; Chen, F.; Zhao, L.; Jiang, M.; Ma, B. Association between Frailty and Mortality, Falls, and Hospitalization among Patients with Hypertension: A Systematic Review and Meta-Analysis BioMed Research International,2021;2021():2690296 | Samples not community dwelling |
| 162 | Huang, M. H.; Hile, E.; Croarkin, E.; Wampler-Kuhn, M.; Blackwood, J.; Colon, G.; PfalzeRandom, L. A. Academy of Oncologic Physical Therapy EDGE Task Force: A Systematic Review of Measures of Balance in Adult Cancer Survivors Rehabilitation Oncology,2019;37(3):92-103 | Study designs not prospective |
| 163 | HunteRandom, Kathleen F; Wagg, Adrian; Kerridge, Teresa; Chick, Hope; Chambers, Thane Falls risk reduction and treatment of overactive bladder symptoms with antimuscarinic agents: a scoping review. Neurourology and urodynamics,2011;30(4):490-4 | Samples not community dwelling |
| 164 | HunteRandom, Susan W; BatcheloRandom, Frances; Hill, Keith D; Hill, Anne-Marie; Mackintosh, Shylie; Payne, Michael Risk Factors for Falls in People With a Lower Limb Amputation: A Systematic Review. PM & R : the journal of injury, function, and rehabilitation,2017;9(2):170-180.e1 | Age not mean of 60 years + |
| 165 | IQBAL S., MUHAMMAD; BAHMAN J.; ASLINDA C. M. Prevalence of Falls and Its Characteristics among Malaysian Older Adults: A Review. Medicine & Health (Universiti Kebangsaan Malaysia),2020;15(1):18-33 | Samples not community dwelling |
| 166 | IyeRandom, Shoba; Naganathan, Vasi; McLachlan, Andrew J; Le CouteuRandom, David G Medication withdrawal trials in people aged 65 years and older: a systematic review. Drugs & aging,2008;25(12):1021-31 | Review purpose not looking at falls |
| 167 | Jahantabi-Nejad, Seifollah; Azad, Akram Predictive accuracy of performance oriented mobility assessment for falls in older adults: A systematic review. Medical journal of the Islamic Republic of Iran,2019;33(8910777):38 | Samples not community dwelling |
| 168 | Jansen, Sofie; Bhangu, Jaspreet; de Rooij, Sophia; Daams, Joost; Kenny, Rose Anne; van der Velde, Nathalie The Association of Cardiovascular Disorders and Falls: A Systematic Review. Journal of the American Medical Directors Association,2016;17(3):193-9 | Samples not community dwelling |
| 169 | Jiam, Nicole Tin-Lok; Li, Carol; Agrawal, Yuri Hearing loss and falls: A systematic review and meta-analysis. The Laryngoscope,2016;126(11):2587-2596 | Study designs not prospective |
| 170 | Jiang, Y.; Wang, M.; Liu, S.; Ya, X.; Duan, G.; Wang, Z. The association between sedentary behavior and falls in older adults: A systematic review and meta-analysis Frontiers in public health,2022;10():1019551 | Samples not community dwelling |
| 171 | Job, Mirko; DottoRandom, Alberto; Viceconti, Antonello; Testa, Marco Ecological Gait as a Fall Indicator in Older Adults: A Systematic Review. The Gerontologist,2020;60(5):e395-e412 | Study designs not prospective |
| 172 | Johnson, R L; Kopp, S L; Hebl, J R; Erwin, P J; Mantilla, C B Falls and major orthopaedic surgery with peripheral nerve blockade: a systematic review and meta-analysis. British journal of anaesthesia,2013;110(4):518-28 | Age not mean of 60 years + |
| 173 | Johnston, Kylie N; PotteRandom, Adrian J; Phillips, Anna Measurement Properties of Short Lower Extremity Functional Exercise Tests in People With Chronic Obstructive Pulmonary Disease: Systematic Review. Physical therapy,2017;97(9):926-943 | Review purpose not looking at falls |
| 174 | Joseph, Alex; KumaRandom, Dhasarathi; Bagavandas, M. A Review of Epidemiology of Fall among Elderly in India. Indian Journal of Community Medicine,2019;44(2):166-168 | Study designs not prospective |
| 175 | K. Karlsson, Magnus; Nordqvist, Anders; Karlsson, Caroline Physical activity, muscle function, falls and fractures. Food & Nutrition Research,2008;52():1-N.PAG | Review design |
| 176 | Kahlaee, Hamid Reza; Latt, Mark D; SchneideRandom, Carl R Association Between Chronic or Acute Use of Antihypertensive Class of Medications and Falls in Older Adults. A Systematic Review and Meta-Analysis. American journal of hypertension,2018;31(4):467-479 | Samples not community dwelling |
| 177 | Kalula, Sebastiana Zimba; Scott, Vicky; Dowd, Andrea; Brodrick, Kathleen Falls and fall prevention programmes in developing countries: environmental scan for the adaptation of the Canadian Falls prevention curriculum for developing countries. Journal of safety research,2011;42(6):461-72 | Samples not community dwelling |
| 178 | Kanagaratnam, Lukshe; Drame, Moustapha; Trenque, Thierry; Oubaya, Nadia; Nazeyrollas, Pierre; Novella, Jean-Luc; Jolly, Damien; Mahmoudi, Rachid Adverse drug reactions in elderly patients with cognitive disorders: A systematic review. Maturitas,2016;85(mwn, 7807333):56-63 | Review purpose not looking at falls |
| 179 | Kannenberg, A.; Zacharias, B.; Probsting, E. Benefits of microprocessor-controlled prosthetic knees to limited community ambulators: Systematic review Journal of Rehabilitation Research and Development,2014;51(10):1469-1495 | Age not mean of 60 years + |
| 180 | Kansagara, D.; Freeman, M.; Low, A.; Motu'apuaka, M.; Weiss, J. W.; PaynteRandom, R.; Kondo, K.; Fu, R.; Kerfoot, A. L. Benefits and harms of treating blood pressure in older adults: A systematic review and meta-analysis Journal of General Internal Medicine,2016;31(2 SUPPL. 1):S139 | Samples not community dwelling |
| 181 | Karavatas, Spiridon G.; Eugene, Roxanne; Evans, Bernardine S. The Link Between Geriatric Depression and Functional Mobility in Older Adults: A Systematic Review. Journal of the National Society of Allied Health,2020;17(1):36-45 | Samples not community dwelling |
| 182 | Kay-Rivest, Emily; SchlacteRandom, Jamie; Waltzman, Susan B. Cochlear implantation outcomes in the older adult: a scoping review Cochlear implants international,2022;23(5):280-290 | Study designs not prospective |
| 183 | Keglovits, Marian; Clemson, Lindy; Hu, Yi-Ling; Nguyen, An; Neff, Anna J; Mandelbaum, Caren; Hudson, Margaret; Williams, Rebecca; Silianoff, Tara; Stark, Susan A scoping review of fall hazards in the homes of older adults and development of a framework for assessment and intervention. Australian occupational therapy journal,2020;67(5):470-478 | Study designs not prospective |
| 184 | Kim, D.; Brown, R.; Berry, S. Cognitive enhancers and risk of fall-related adverse events - Systematic review and meta-analysis of randomized controlled trials Journal of the American Geriatrics Society,2010;58(SUPPL. 1):S99 | Other |
| 185 | Kim, Kwang-Il; Jung, Hye-Kyung; Kim, Chang Oh; Kim, Soo-Kyung; Cho, Hyun-Ho; Kim, Dae Yul; Ha, Yong-Chan; Hwang, Sung-Hee; Won, Chang Won; Lim, Jae-Young; Kim, Hyun Jung; Kim, Jae Gyu; Korean Association of Internal Medicine, The Korean Geriatrics Society Evidence-based guidelines for fall prevention in Korea. The Korean journal of internal medicine,2017;32(1):199-210 | Review design |
| 186 | King, Mary B.; Tinetti, Mary E. Falls in community-dwelling older persons Journal of the American Geriatrics Society,1995;43(10):1146-1154 | Review design |
| 187 | Knoop, V.; Cloots, B.; Costenoble, A.; Debain, A.; Vella Azzopardi, R.; Vermeiren, S.; Bautmans, I.; Verte, D.; BeyeRandom, I.; Petrovic, M.; De DondeRandom, L.; Kardol, T.; Rossi, G.; Clarys, P.; Scafoglieri, A.; Cattrysse, E.; de Hert, P.; Jansen, B. Fatigue and the prediction of negative health outcomes: A systematic review with meta-analysis Ageing Research Reviews,2021;67():101261 | Review purpose not looking at falls |
| 188 | Kozinc, Ziga; LofleRandom, Stefan; HofeRandom, Christian; Carraro, Ugo; Sarabon, Nejc Diagnostic Balance Tests for Assessing Risk of Falls and Distinguishing Older Adult Fallers and Non-Fallers: A Systematic Review with Meta-Analysis. Diagnostics (Basel, Switzerland),2020;10(9): | Study designs not prospective |
| 189 | Krishnaswamy, N.; Zullo, A.; Dore, D. D. A systematic review of the adverse effects of atypical antipsychotics in the elderly population Pharmacoepidemiology and Drug Safety,2013;22(SUPPL. 1):242-243 | Other |
| 190 | KronzeRandom, V. L.; Wildes, T. M.; Stark, S. L.; Avidan, M. S. Review of perioperative falls. BJA: The British Journal of Anaesthesia,2016;117(6):720-732 | Samples not community dwelling |
| 191 | Kudlac, Megan; Sabol, Joseph; KaiseRandom, Katelynn; Kane, Cecelia; Phillips, Robert S. Reliability and Validity of the Berg Balance Scale in the Stroke Population: A Systematic Review Physical and Occupational Therapy in Geriatrics,2019;37(3):196-221 | Review purpose not looking at falls |
| 192 | Kuljeerung, Orawan; Lach, Helen W. Extrinsic and Behavioral Fall Risk Factors in People With Parkinson's Disease: An Integrative Review Rehabilitation Nursing,2021;46(1):45361 | Review design |
| 193 | Kulkarni, Snehal; NagarkaRandom, Aarti A Systematic Review and Meta-Analysis of the Prevalence of Falls Among Community-dwelling Older Adults in India Indian Journal of Gerontology,2021;35(3):456-469 | Journal not peer reviewed |
| 194 | Kwan, Marcella Mun-San; Close, Jacqueline C T; Wong, Alfred Kwok Wai; Lord, Stephen R Falls incidence, risk factors, and consequences in Chinese older people: a systematic review. Journal of the American Geriatrics Society,2011;59(3):536-43 | Study designs not prospective |
| 195 | Lachance, Chantelle C; Jurkowski, Michal P; Dymarz, Ania C; Robinovitch, Stephen N; Feldman, Fabio; Laing, Andrew C; Mackey, Dawn C Compliant flooring to prevent fall-related injuries in older adults: A scoping review of biomechanical efficacy, clinical effectiveness, cost-effectiveness, and workplace safety. PloS one,2017;12(2):e0171652 | Review purpose not looking at falls |
| 196 | Lapane, Kate L; Yang, Shibing; Brown, Monique J; JawahaRandom, Rachel; Pagliasotti, Caleb; Rajpathak, Swapnil Sulfonylureas and risk of falls and fractures: a systematic review. Drugs & aging,2013;30(7):527-47 | Study designs not prospective |
| 197 | Le Berre, M.; Dumoulin, C. Strength, balance, mobility and function in elderly women with urinary incontinence: A review of the literature Kinesitherapie,2020;20(226):45371 | Language |
| 198 | Lee, Jacob; GelleRandom, Andrew I; StrasseRandom, Dale C Analytical review: focus on fall screening assessments. PM & R : the journal of injury, function, and rehabilitation,2013;5(7):609-21 | Review design |
| 199 | Lee, Junga The association between physical activity and risk of falling in older adults: A systematic review and meta-analysis of prospective cohort studies. Geriatric nursing (New York, N.Y.),2020;(8309633, FW7): | Samples not community dwelling |
| 200 | Lee, Justin; Negm, Ahmed; Peters, Ryan; Wong, Eric KC; Holbrook, Anne Deprescribing fall-risk increasing drugs (FRIDs) for the prevention of falls and fall-related complications: a systematic review and meta-analysis BMJ open,2021;11(2):e035978 | Study designs not prospective |
| 201 | Lee, Kayoung; PressleRandom, Susan J; TitleRandom, Marita Falls in Patients With Heart Failure: A Systematic Review. The Journal of cardiovascular nursing,2016;31(6):555-561 | Samples not community dwelling |
| 202 | Lee, V.; Balucani, C.; DeLuca, J.; Lederman, Y. S.; Arnedo, V.; Lushbough, C. A.; Levine, S. R. Post-stroke fatigue: A systematic evidence-based critique Stroke,2012;43(2 Meeting Abstracts): | Other |
| 203 | Leipzig, R M; Cumming, R G; Tinetti, M E Drugs and falls in older people: a systematic review and meta-analysis: I. Psychotropic drugs. Journal of the American Geriatrics Society,1999;47(1):45565 | Study designs not prospective |
| 204 | Leipzig, R M; Cumming, R G; Tinetti, M E Drugs and falls in older people: a systematic review and meta-analysis: II. Cardiac and analgesic drugs. Journal of the American Geriatrics Society,1999;47(1):40-50 | Samples not community dwelling |
| 205 | LevingeRandom, P.; Wallman, S.; Hill, K. Balance dysfunction and falls in people with lower limb arthritis: factors contributing to risk and effectiveness of exercise interventions European Review of Aging and Physical Activity,2012;9(1):17-25 | Review purpose not looking at falls |
| 206 | Li, Y.; Hou, L.; Zhao, H.; Xie, R.; Yi, Y.; Ding, X. Risk factors for falls among community-dwelling older adults: A systematic review and meta-analysis Frontiers in Medicine,2023;9():1019094 | Age not mean of 60 years + |
| 207 | Lim, Grace Rui Si; Ng, Caitlin Hsuen; Kwan, Yu Heng; Fong, Warren Prevalence and risk factors for falls in patients with spondyloarthritis: A systematic review International journal of rheumatic diseases,2021;24(5):623-632 | Age not mean of 60 years + |
| 208 | Lisibach, Angela; Benelli, Valerie; Ceppi, Marco Giacomo; Waldner-KnogleRandom, Karin; Csajka, Chantal; Lutters, Monika Quality of anticholinergic burden scales and their impact on clinical outcomes: a systematic review. European journal of clinical pharmacology,2020;(en4, 1256165): | Review purpose not looking at falls |
| 209 | Liu, Hao; Frank, Adam Tai chi as a balance improvement exercise for older adults: a systematic review. Journal of geriatric physical therapy (2001),2010;33(3):103-9 | Review purpose not looking at falls |
| 210 | Liu, Yang; Yang, Yanjiang; Liu, Hao; Wu, Wenyuan; Wu, Xintao; Wang, Tao A systematic review and meta-analysis of fall incidence and risk factors in elderly patients after total joint arthroplasty. Medicine,2020;99(50):e23664 | Samples not community dwelling |
| 211 | Lo, A.; CarpenteRandom, C. R. Predicting geriatric falls following emergency department care: A systematic review Annals of Emergency Medicine,2014;64(4 SUPPL. 1):S78 | Other |
| 212 | Lo, C W T; Tsang, W W N; Yan, C H; Lord, S R; Hill, K D; Wong, A Y L Risk factors for falls in patients with total hip arthroplasty and total knee arthroplasty: a systematic review and meta-analysis. Osteoarthritis and cartilage,2019;27(7):979-993 | Samples not community dwelling |
| 213 | Lo, C. W. T.; Lin, C. Y.; Tsang, W. W. N.; Yan, C. H.; Wong, A. Y. L. Psychometric Properties of Brief-Balance Evaluation Systems Test Among Multiple Populations: A Systematic Review and Meta-analysis Archives of Physical Medicine and Rehabilitation,2022;103(1):155-175.e2 | Review purpose not looking at falls |
| 214 | Lockwood, Kylee J; TayloRandom, Nicholas F; Harding, Katherine E Pre-discharge home assessment visits in assisting patients' return to community living: A systematic review and meta-analysis. Journal of rehabilitation medicine,2015;47(4):289-99 | Samples not community dwelling |
| 215 | Lord, Stephen R. Visual risk factors for falls in older people Age and Ageing,2006;35(Suppl. 2):ii42-ii45 | Review design |
| 216 | Lord, Stephen R.; Menz, Hylton B.; Sherrington, Catherine Home environment risk factors for falls in older people and the efficacy of home modifications Age and Ageing,2006;35(Suppl. 2):ii55-ii59 | Review design |
| 217 | Lu, Feng-Ping; Lin, Kun-Pei; Kuo, Hsu-Ko Diabetes and the risk of multi-system aging phenotypes: a systematic review and meta-analysis. PloS one,2009;4(1):e4144 | Review purpose not looking at falls |
| 218 | Lu, Xiaojie; Park, Nam-Kyu; Ahrentzen, Sherry Lighting Effects on Older Adults' Visual and Nonvisual Performance: A Systematic Review Journal of Housing for the Elderly,2019;33(3):298-324 | Review purpose not looking at falls |
| 219 | Lukaszyk, Caroline; Harvey, Lara; Sherrington, Cathie; Keay, Lisa; Tiedemann, Anne; Coombes, Julieann; Clemson, Lindy; Ivers, Rebecca Risk factors, incidence, consequences and prevention strategies for falls and fall-injury within older indigenous populations: a systematic review. Australian and New Zealand journal of public health,2016;40(6):564-568 | Age not mean of 60 years + |
| 220 | Lunt, Eleanor; Ong, Terence; Gordon, Adam L; Greenhaff, Paul L; Gladman, John R F The clinical usefulness of muscle mass and strength measures in older people: a systematic review. Age and ageing,2020;(0375655, 2xr): | Samples not community dwelling |
| 221 | Lunt, Eleanor; Ong, Terence; Gordon, Adam L.; Greenhaff, Paul L.; Gladman, John R. F. The clinical usefulness of muscle mass and strength measures in older people: a systematic review Age and Ageing,2021;50(1):88-95 | Samples not community dwelling |
| 222 | Lynds, M. E.; Arnold, C. M. Fall Risk Screening and Assessment for People Living With Dementia: A Scoping Review Journal of Applied Gerontology,2023;(): | Review purpose not looking at falls |
| 223 | Macdonald, John B. Role of drugs in falls in the elderly Clinics in Geriatric Medicine,1985;1(3):621-636 | Review design |
| 224 | Maciaszek, Janusz; Osinski, W The effects of Tai Chi on body balance in elderly people--a review of studies from the early 21st century. The American journal of Chinese medicine,2010;38(2):219-29 | Review purpose not looking at falls |
| 225 | MacKay, Scott; Ebert, Patricia; Harbidge, Cathy; Hogan, David B. Fear of Falling in Older Adults: A Scoping Review of Recent Literature Canadian Geriatrics Journal,2021;24(4):379-394 | Review purpose not looking at falls |
| 226 | Malik, V.; GallagheRandom, C.; Linz, D.; Elliott, A.; Agbaedeng, T.; Emami, M.; Mishima, R.; Hendriks, J.; Mahajan, R.; Arnolda, L. F.; Lau, D. H.; Sanders, P. Atrial fibrillation is an independent predictor of falls risk, syncope & orthostatic intolerance in older adults: A systematic review and meta-analysis Europace,2019;21(Supplement 2):ii860 | Other |
| 227 | Malik, V.; GallagheRandom, C.; Linz, D.; Elliott, A.; Emami, M.; Kadhim, K.; Mishima, R.; Hendriks, J.; Mahajan, R.; Arnolda, L.; Sanders, P.; Lau, D. Atrial Fibrillation is Independently Associated with Both Syncope and the Risk of Falling in Older Adults: A Systematic Review and Meta-Analysis. Heart, Lung & Circulation,;28():S202-S203 | Other |
| 228 | Malik, Varun; GallagheRandom, Celine; Linz, Dominik; Elliott, Adrian D; Emami, Mehrdad; Kadhim, Kadhim; Mishima, Ricardo; Hendriks, Jeroen M L; Mahajan, Rajiv; Arnolda, Leonard; Sanders, Prashanthan; Lau, Dennis H Atrial Fibrillation Is Associated With Syncope and Falls in Older Adults: A Systematic Review and Meta-analysis. Mayo Clinic proceedings,2020;95(4):676-687 | Study designs not prospective |
| 229 | Manias, Elizabeth; KabiRandom, Md Zunayed; MaieRandom, Andrea B. Inappropriate medications and physical function: a systematic review Therapeutic advances in drug safety,2021;12():20420986211030300 | Samples not community dwelling |
| 230 | Mansbart, Felix; KienbergeRandom, Gerda; S√∂nnichsen, Andreas; Mann, Eva Efficacy and safety of adrenergic alpha-1 receptor antagonists in older adults: a systematic review and meta-analysis supporting the development of recommendations to reduce potentially inappropriate prescribing BMC Geriatrics,2022;22(1):45320 | Study designs not prospective |
| 231 | Marks, R.; Allegrante, J. P. Falls-prevention programs for older ambulatory community dwellers: From public health research to health promotion policy Sozial- und Praventivmedizin,2004;49(3):171-178 | Review design |
| 232 | Marshall, Samantha; Gabiazon, Raphael; Persaud, Priyanka; Nagamatsu, Lindsay S. What do functional neuroimaging studies tell us about the association between falls and cognition in older adults? A systematic review Ageing research reviews,2023;85():101859 | Review purpose not looking at falls |
| 233 | Masud, Tahir; Morris, Robert O. Epidemiology of falls Age and Ageing,2001;30(Suppl. 4):45358 | Review design |
| 234 | Mattishent, K.; Loke, Y. K. Meta-Analysis: Association Between Hypoglycemia and Serious Adverse Events in Older Patients Treated With Glucose-Lowering Agents Frontiers in Endocrinology,2021;12():571568 | Samples not community dwelling |
| 235 | Mattishent, Katharina; Loke, Yoon Kong Meta-analysis: Association between hypoglycaemia and serious adverse events in older patients. Journal of diabetes and its complications,2016;30(5):811-8 | Study designs not prospective |
| 236 | Maximos, Mira; Chang, Feng; Patel, Tejal Risk of falls associated with antiepileptic drug use in ambulatory elderly populations: A systematic review. Canadian pharmacists journal : CPJ = Revue des pharmaciens du Canada : RPC,2017;150(2):101-111 | Study designs not prospective |
| 237 | Maximos, Mira; Chang, Feng; Patel, Tejal Risk of falls associated with antiepileptic drug use in ambulatory elderly populations. Canadian Pharmacists Journal (Sage Publications Inc.),2017;150(2):101-111 | Other |
| 238 | Mazeaud, S.; Castellana, F.; Coelho-JunioRandom, H. J.; Panza, F.; Rondanelli, M.; Fassio, F.; De Pergola, G.; Zupo, R.; Sardone, R. Coffee Drinking and Adverse Physical Outcomes in the Aging Adult Population: A Systematic Review Metabolites,2022;12(7):654 | Study designs not prospective |
| 239 | McCarthy, F.; Fan, C. W.; Kearney, P. M.; Walsh, C.; Kenny, R. A. What is the evidence for cardiovascular disorders as a risk factor for non-syncopal falls? Scope for future research European Geriatric Medicine,2010;1(4):244-251 | Samples not community dwelling |
| 240 | McKenzie, Katherine; Martin, Lynn; Ouellette-Kuntz, H√©l√®ne Frailty and Intellectual and Developmental Disabilities: a Scoping Review. Canadian Geriatrics Journal,2016;19(3):103-112 | Review purpose not looking at falls |
| 241 | McMillan, J. M.; Michalchuk, Q.; Goodarzi, Z. Frailty in Parkinson's disease: A systematic review and meta-analysis Clinical Parkinsonism and Related Disorders,2021;4():100095 | Review purpose not looking at falls |
| 242 | Mehdizadeh, D.; Hale, M.; Todd, O.; Zaman, H.; Marques, I.; Petty, D.; Alldred, D. P.; Johnson, O.; Faisal, M.; GardneRandom, P.; Clegg, A. Associations Between Anticholinergic Medication Exposure and Adverse Health Outcomes in Older People with Frailty: A Systematic Review and Meta-analysis Drugs - Real World Outcomes,2021;8(4):431-458 | Samples not community dwelling |
| 243 | Mehdizadeh, S.; Van Ooteghem, K.; Gulka, H.; Nabavi, H.; Faieghi, M.; Taati, B.; Iaboni, A. A systematic review of center of pressure measures to quantify gait changes in older adults Experimental Gerontology,2021;143():111170 | Samples not community dwelling |
| 244 | Mei, Fan; Gao, Qianqian; Chen, Fei; Zhao, Li; Shang, Yi; Hu, Kaiyan; Zhang, Weiyi; Zhao, Bing; Ma, Bin Frailty as a Predictor of Negative Health Outcomes in Chronic Kidney Disease: A Systematic Review and Meta-Analysis Journal of the American Medical Directors Association,2021;22(3):535-535 | Samples not community dwelling |
| 245 | Menant, Jasmine C; Schoene, Daniel; Sarofim, Mina; Lord, Stephen R Single and dual task tests of gait speed are equivalent in the prediction of falls in older people: a systematic review and meta-analysis. Ageing research reviews,2014;16(101128963):83-104 | Samples not community dwelling |
| 246 | Menant, Jasmine C; Steele, Julie R; Menz, Hylton B; Munro, Bridget J; Lord, Stephen R Optimizing footwear for older people at risk of falls. Journal of rehabilitation research and development,2008;45(8):1167-81 | Review design |
| 247 | Menz, H. B.; Auhl, M.; Spink, M. J. Foot Problems as a Risk Factor for Falls in Community-Dwelling Older People: A Systematic Review and Meta-Analysis... [including Commentary by Robert A. Eckles]. Foot & Ankle Quarterly--The Seminar Journal,2019;30(3):169-171 | Journal not peer reviewed |
| 248 | Michael, W.; Couture, A. D.; Swedlund, M.; Hampton, A.; Eglash, A.; SchrageRandom, S. An Evidence-Based Review of Vitamin D for Common and High-Mortality Conditions Journal of the American Board of Family Medicine,2022;35(6):1217-1229 | Review design |
| 249 | Min, Yaena; Slattum, Patricia W Poor Sleep and Risk of Falls in Community-Dwelling Older Adults: A Systematic Review. Journal of applied gerontology : the official journal of the Southern Gerontological Society,2018;37(9):1059-1084 | Study designs not prospective |
| 250 | Moayyeri, Alireza The association between physical activity and osteoporotic fractures: a review of the evidence and implications for future research. Annals of epidemiology,2008;18(11):827-35 | Age not mean of 60 years + |
| 251 | Modaberi, Shaghayegh; Saemi, Esmaeel; Federolf, Peter A.; van Andel, Steven A Systematic Review on Detraining Effects after Balance and Fall Prevention Interventions Journal of clinical medicine,2021;10(20): | Review purpose not looking at falls |
| 252 | Modarresi, Shirin; Divine, Alison; Grahn, Jessica A.; Overend, Tom J.; HunteRandom, Susan W. Gait parameters and characteristics associated with increased risk of falls in people with dementia: a systematic review International Psychogeriatrics,2019;31(9):1287-1303 | Other |
| 253 | Mohamed, M. R.; Ramsdale, E.; Loh, K. P.; Arastu, A.; Xu, H.; Obrecht, S.; Castillo, D.; Sharma, M.; Holmes, H. M.; Nightingale, G.; Juba, K. M.; Mohile, S. G. Associations of Polypharmacy and Inappropriate Medications with Adverse Outcomes in Older Adults with Cancer: A Systematic Review and Meta-Analysis The oncologist,2019;(): | Review purpose not looking at falls |
| 254 | Mohamed, Mostafa R; Ramsdale, Erika; Loh, Kah Poh; Arastu, Asad; Xu, Huiwen; Obrecht, Spencer; Castillo, Daniel; Sharma, Manvi; Holmes, Holly M; Nightingale, Ginah; Juba, Katherine M; Mohile, Supriya G Associations of Polypharmacy and Inappropriate Medications with Adverse Outcomes in Older Adults with Cancer: A Systematic Review and Meta-Analysis. The oncologist,2020;25(1):e94-e108 | Review purpose not looking at falls |
| 255 | Mol, Arjen; Bui Hoang, Phuong Thanh Silvie; Sharmin, Sifat; Reijnierse, Esmee M; van Wezel, Richard J A; Meskers, Carel G M; MaieRandom, Andrea B Orthostatic Hypotension and Falls in Older Adults: A Systematic Review and Meta-analysis. Journal of the American Medical Directors Association,2019;20(5):589-597.e5 | Study designs not prospective |
| 256 | Mollinedo-Cardalda, I.; Carral, J. M. C.; Rodriguez-Fuentes, G. Timed up and go and its application in older adults with Parkinson's disease. systematic review Parkinsonism and Related Disorders,2016;22(SUPPL. 2):e53 | Other |
| 257 | Montero‚ÄêOdasso, Manuel; Speechley, Mark Falls in Cognitively Impaired Older Adults: Implications for Risk Assessment And Prevention. Journal of the American Geriatrics Society,2018;66(2):367-375 | Review design |
| 258 | Moon, Shinje; Chung, Hye Soo; Kim, Yoon Jung; Kim, Sung Jin; Kwon, Ohseong; Lee, Young Goo; Yu, Jae Myung; Cho, Sung Tae The impact of urinary incontinence on falls: A systematic review and meta-analysis PloS one,2021;16(5):e0251711 | Study designs not prospective |
| 259 | Moore, Heather; Boltong, Anna Don't fall for weight: A systematic review of weight status and falls. Nutrition & Dietetics,2011;68(4):273-279 | Samples not community dwelling |
| 260 | Moore, Martha; BarkeRandom, Karen The validity and reliability of the four square step test in different adult populations: a systematic review. Systematic reviews,2017;6(1):187 | Review purpose not looking at falls |
| 261 | Moreland, Julie; Richardson, Julie; Chan, David H; O'Neill, John; Bellissimo, Agostino; Grum, Rosa Maria; Shanks, Lynne Evidence-based guidelines for the secondary prevention of falls in older adults. Gerontology,2003;49(2):93-116 | Review design |
| 262 | Morfis, Pavlos; Gkaraveli, Maria Effects of aging on biomechanical gait parameters in the healthy elderly and the risk of falling Journal of Research & Practice on the Musculoskeletal System (JRPMS),2021;5(2):59-64 | Review purpose not looking at falls |
| 263 | Mortaza, N; Abu Osman, N A; Mehdikhani, N Are the spatio-temporal parameters of gait capable of distinguishing a faller from a non-faller elderly?. European journal of physical and rehabilitation medicine,2014;50(6):677-91 | Study designs not prospective |
| 264 | Mossey, Jana M. Social and psychologic factors related to falls among the elderly Clinics in Geriatric Medicine,1985;1(3):541-553 | Samples not community dwelling |
| 265 | Mota Sousa, Lu√≠s Manuel; Alves Marques-Vieira, Cristina Maria; Nogueira de Caldevilla, Maria Nilza Guimar√£es; Alves Dias Henriques, Cristina Maria; Pedro Severino, Sandy Silva; Alves Caldeira, S√≠lvia Maria Risk for falls among community-dwelling older people: systematic literature review. Revista Gaucha de Enfermagem,2016;37(4):45300 | Study designs not prospective |
| 266 | Moutzouri, M.; Gleeson, N.; Billis, E.; Tsepis, E.; Panoutsopoulou, I.; Gliatis, J. The effect of total knee arthroplasty on patients' balance and incidence of falls: a systematic review Knee Surgery Sports Traumatology Arthroscopy,2017;25(11):3439-3451 | Samples not community dwelling |
| 267 | MuhlbaueRandom, Viktoria; DallmeieRandom, Dhayana; Brefka, Simone; Bollig, Claudia; Voigt-Radloff, Sebastian; DenkingeRandom, Michael The Pharmacological Treatment of Arterial Hypertension in Frail, Older Patients-a Systematic Review. Deutsches Arzteblatt international,2019;116(3):23-30 | Review purpose not looking at falls |
| 268 | Myint, Zin W.; Momo, Harry D.; Otto, Danielle E.; Yan, Donglin; Wang, Peng; KolesaRandom, Jill M. Evaluation of Fall and Fracture Risk Among Men With Prostate Cancer Treated With Androgen Receptor Inhibitors: A Systematic Review and Meta-analysis. JAMA Network Open,2020;3(11):e2025826-e2025826 | Samples not community dwelling |
| 269 | N√≥brega Medeiros, Emilene; Lima da N√≥brega, Maria Miriam; de Lourdes de Farias Pontes, Maria; de Fran√ßa Vasconcelos, Marileuza Maria; do Socorro Guedes de Paiva, Maria; Silva Paredes Moreira, Maria Adelaide Determinants of risk of falls among elderly: a systematic study... content seems to change in results sections to different topic. Revista de Pesquisa: Cuidado e Fundamental,;6(5):111-120 | Language |
| 270 | Neuls, Patrick D; Clark, Tammie L; Van Heuklon, Nicole C; ProctoRandom, Joy E; KilkeRandom, Barbra J; BiebeRandom, Mallory E; Donlan, Alice V; Carr-Jules, Suchitha A; Neidel, William H; Newton, Roberta A Usefulness of the Berg Balance Scale to predict falls in the elderly. Journal of geriatric physical therapy (2001),2011;34(1):45361 | Samples not community dwelling |
| 271 | Neville, Christopher; Nguyen, Hung; Ross, Kim; Wingood, Mariana; Peterson, Elizabeth Walker; Dewitt, James E; Moore, Jonathan; King, Michael J; Atanelov, Levan; White, Josh; Najafi, Bijan Lower-Limb Factors Associated with Balance and Falls in Older Adults: A Systematic Review and Clinical Synthesis. Journal of the American Podiatric Medical Association,2020;110(5): | Age not mean of 60 years + |
| 272 | Ng, Chin Teck; Tan, Maw Pin Osteoarthritis and falls in the older person. Age & Ageing,2013;42(5):561-566 | Review design |
| 273 | Nicklett, Emily J; Kadell, Andria R Fruit and vegetable intake among older adults: a scoping review. Maturitas,2013;75(4):305-12 | Review purpose not looking at falls |
| 274 | Nilsagard, Y; Gunn, H; Freeman, J; Hoang, P; Lord, S; MazumdeRandom, Rajarshi; Cameron, Michelle Falls in people with MS--an individual data meta-analysis from studies from Australia, Sweden, United Kingdom and the United States. Multiple sclerosis (Houndmills, Basingstoke, England),2015;21(1):92-100 | Age not mean of 60 years + |
| 275 | Noguchi, Naomi; Chan, Lewis; Cumming, Robert G; Blyth, Fiona M; Naganathan, Vasi A systematic review of the association between lower urinary tract symptoms and falls, injuries, and fractures in community-dwelling older men. The aging male : the official journal of the International Society for the Study of the Aging Male,2016;19(3):168-174 | Study designs not prospective |
| 276 | Nothern, A. C.; Berry, S. Hemophilia: Are we missing frailty? Journal of the American Geriatrics Society,2017;65(Supplement 1):S75 | Other |
| 277 | Nowson, C A; Service, C; Appleton, J; GriegeRandom, J A The Impact of Dietary Factors on Indices of Chronic Disease in Older People: A Systematic Review. The journal of nutrition, health & aging,2018;22(2):282-296 | Review purpose not looking at falls |
| 278 | O'Hare, Margaret P; Pryde, Shona J; Gracey, Jacqueline H A systematic review of the evidence for the provision of walking frames for older people. Physical Therapy Reviews,2013;18(1):45619 | Review purpose not looking at falls |
| 279 | O'Neil, Christine K; Hanlon, Joseph T; Marcum, Zachary A Adverse effects of analgesics commonly used by older adults with osteoarthritis: focus on non-opioid and opioid analgesics. The American journal of geriatric pharmacotherapy,2012;10(6):331-42 | Review purpose not looking at falls |
| 280 | Obradoviƒá, Nata≈°a; Lagueux, √âmilie; Michaud, Fr√©d√©ric; ProvencheRandom, V√©ronique Pros and cons of pet ownership in sustaining independence in community-dwelling older adults: a scoping review Ageing and Society,2020;40(9):2061-2076 | Review purpose not looking at falls |
| 281 | Ohikere, K.; Veracruz, N.; Wong, R. J. Cognitive Impairment and Cirrhosis in Older Patients: A Systematic Review Gerontology and Geriatric Medicine,2022;8(): | Samples not community dwelling |
| 282 | Okubo, Yoshiro; Schoene, Daniel; Caetano, Maria Jd; PlineRandom, Erika M; Osuka, Yosuke; Toson, Barbara; Lord, Stephen R Stepping impairment and falls in older adults: A systematic review and meta-analysis of volitional and reactive step tests. Ageing research reviews,2020;(101128963):101238 | Samples not community dwelling |
| 283 | Oliveira, C. C.; Annoni, R.; Lee, A. L.; McGinley, J.; Irving, L. B.; Denehy, L. Falls prevalence and risk factors in people with chronic obstructive pulmonary disease: A systematic review Respiratory Medicine,2021;176():106284 | Samples not community dwelling |
| 284 | Oliveira, Cristino C; Annoni, Raquel; Lee, Annemarie L; McGinley, Jennifer; Irving, Louis B; Denehy, Linda Falls prevalence and risk factors in people with chronic obstructive pulmonary disease: A systematic review. Respiratory medicine,2020;176(8908438, rme):106284 | Samples not community dwelling |
| 285 | Ortega-Bastidas, Paulina; Gomez, Britam; Aqueveque, Pablo; Luarte-Martinez, Soledad; Cano-de-la-Cuerda, Roberto Instrumented Timed Up and Go Test (iTUG)-More Than Assessing Time to Predict Falls: A Systematic Review Sensors (Basel, Switzerland),2023;23(7): | Review purpose not looking at falls |
| 286 | Pahlevanian, Ali Akbar; Najarian, Reyhaneh; Adabi, Sadegh; Mirshoja, Mina Sadat The Prevalence of Fall and Related Factors in Iranian Elderly: A Systematic Review Archives of Rehabilitation,2020;21(3):286-303 | Language |
| 287 | Pamoukdjian, Frederic; Paillaud, Elena; Zelek, Laurent; Laurent, Marie; Levy, Vincent; Landre, Thierry; Sebbane, Georges Measurement of gait speed in older adults to identify complications associated with frailty: A systematic review. Journal of geriatric oncology,2015;6(6):484-96 | Review purpose not looking at falls |
| 288 | Pana, Anastasia; Sourtzi, Panayota; Kalokairinou, Athina; Pastroudis, Alexandros; Chatzopoulos, Stamatios-Theodoros; Velonaki, Venetia Sofia Association between SRed or perceived fatigue and falls among older people: A systematic review International Journal of Orthopaedic & Trauma Nursing,2021;43():N.PAG-N.PAG | Study designs not prospective |
| 289 | Park, Hyerim; Satoh, Hiroki; Miki, Akiko; Urushihara, Hisashi; Sawada, Yasufumi Medications associated with falls in older people: systematic review of publications from a recent 5-year period. European journal of clinical pharmacology,2015;71(12):1429-40 | Samples not community dwelling |
| 290 | Park, M. K.; Kim, G. S.; Kim, N.; Shim, M. S.; Lee, J. J. The home environment as fall risk factors among community-dwelling frail older people: A systematic review...International Society for Gerontechnology 13th World Conference, October 22-26, 2022, Daegu, South Korea Gerontechnology,2022;21():45292 | Other |
| 291 | Park, Seong-Hi Tools for assessing fall risk in the elderly: a systematic review and meta-analysis. Aging clinical and experimental research,2018;30(1):45307 | Samples not community dwelling |
| 292 | Pearson, Ella; Siskind, Dan; Hubbard, Ruth E.; Gordon, Emily H.; Coulson, Elizabeth J.; Warren, Nicola Frailty and severe mental illness: A systematic review and narrative synthesis Journal of psychiatric research,2022;147():166-175 | Review purpose not looking at falls |
| 293 | Peel, Nancye May Epidemiology of falls in older age Canadian Journal on Aging/La Revue canadienne du vieillissement,2011;30(1):45492 | Review design |
| 294 | PELLICER GARCIA, BEGO√ëA; JU√ÅREZ VELA, RA√öL; GRACIA CARRASCO, EL√çAS; GUERRERO PORTILLO, SANDRA; GARC√çA MOYANO, LORETO MAR√çA; AZ√ìN BELARRE, JOS√â CARLOS Epidemiolog√≠a de ca√≠das en la poblaci√≥n anciana espa√±ola no institucionalizada. Revista Rol de Enfermer√≠a,2015;38(11):40-45 | Other |
| 295 | Pellicer Garcia, Begona; Juarez Vela, Raul; Gracia Carrasco, Elias; Guerrero Portillo, Sandra; Garcia Moyano, Loreto Maria; Azon Belarre, Jose Carlos [EPIDEMIOLOGY OF FALLS IN THE NON-INSTITUTIONALIZED SPANISH ELDERLY POPULATION, SYSTEMATIC REVIEW 2014]. Revista de enfermeria (Barcelona, Spain),2015;38(11):14732 | Language |
| 296 | Pena, Silvana Barbosa; Guimar√£es, Helo√≠sa Cristina Quatrini Carvalho Passos; Lopes, Juliana Lima; Guandalini, Lidia Santiago; Taminato, M√¥nica; Barbosa, Dulce Aparecida; de Barros, Alba L√∫cia Bottura Leite Fear of falling and risk of falling: a systematic review and meta-analysis. Acta Paulista de Enfermagem,2019;32(4):456-463 | Study designs not prospective |
| 297 | Peng, Ke; Tian, Maoyi; Andersen, Melanie; Zhang, Jing; Liu, Yishu; Wang, Qilong; Lindley, Richard; Ivers, Rebecca Incidence, risk factors and economic burden of fall-related injuries in older Chinese people: a systematic review. Injury prevention : journal of the International Society for Child and Adolescent Injury Prevention,2019;25(1):45394 | Study designs not prospective |
| 298 | Perell, Karen L.; Nelson, Audrey; Goldman, Ronald L.; LutheRandom, Stephen L.; Prieto-Lewis, Nicole; Rubenstein, Laurence Z. Fall risk assessment measures: an analytic review Journals of Gerontology: Series A: Biological Sciences and Medical Sciences,2001;56A(12):M761-M766 | Review design |
| 299 | Perry, Bruce C. Falls among the elderly: a review of the methods and conclusions of epidemiologic studies Journal of the American Geriatrics Society,1982;30(6):367-371 | Review design |
| 300 | PfortmuelleRandom, C A; LindneRandom, G; Exadaktylos, A K Reducing fall risk in the elderly: risk factors and fall prevention, a systematic review. Minerva medica,2014;105(4):275-81 | Review design |
| 301 | Pham, V. K.; Yeung, S. S. Y.; Reijnierse, E. M.; Trappenburg, M. C.; Meskers, C. G. M.; MaieRandom, A. B. Sarcopenia and its association with falls and fractures in older adults: A systematic review and meta-analysis Australasian Journal on Ageing,2018;37():47-47 | Samples not community dwelling |
| 302 | Pires, R. E.; Reis, I. G. N.; Waldolato, G. S.; Pires, D. D.; Bidolegui, F.; Giordano, V. What Do We Need to Know about Musculoskeletal Manifestations of COVID-19?: A Systematic Review JBJS Reviews,2022;10(6):e22.00013 | Review purpose not looking at falls |
| 303 | Quicke, J. G.; FosteRandom, N. E.; Thomas, M. J.; Holden, M. A. Is long-term physical activity safe for older adults with knee pain?: A systematic review Osteoarthritis and Cartilage,2015;23(9):1445-1456 | Review purpose not looking at falls |
| 304 | Quinn, G.; CombeRandom, L.; Galvin, R.; Coote, S. The ability of clinical balance measures to identify falls risk in multiple sclerosis: a systematic review and meta-analysis Clinical Rehabilitation,2018;32(5):571-582 | Age not mean of 60 years + |
| 305 | Randolph, Saungaylia; Dalal, Rajiv A.; Asumu, Eevi; Colbert, Vontreia; Isom, Alex T.; Mayfield, Stephanie R. THE EFFECT OF INAPPROPRIATE FOOTWEAR ON GAIT AND FALLS IN OLDER ADULTS: A LITERATURE REVIEW. Journal of the National Society of Allied Health,2017;14(1):13-17 | Review design |
| 306 | Rao, Wen-Wang; Zeng, Liang-Nan; Zhang, Ji-Wen; Zong, Qian-Qian; An, Feng-Rong; Ng, Chee H; Ungvari, Gabor S; Yang, Fang-Yu; Zhang, Juan; Peng, Kelly Z; Xiang, Yu-Tao Worldwide prevalence of falls in older adults with psychiatric disorders: A meta-analysis of observational studies. Psychiatry research,2019;273(qc4, 7911385):114-120 | Samples not community dwelling |
| 307 | Rasmussen, N. H.; Dal, J. Falls and Fractures in DiabetesMore than Bone Fragility Current Osteoporosis Reports,2019;17(3):147-156 | Review design |
| 308 | Rawsky, Elaine Review of the literature on falls among the elderly Image: The Journal of Nursing Scholarship,1998;30(1):47-52 | Age not mean of 60 years + |
| 309 | Redsell S; Cheater F; Juby L Management and prevention of falls in the elderly. Journal of Clinical Governance,2002;10(4):215-222 | Review design |
| 310 | Reid, M Carrington; Boutros, Nashaat N; O'ConnoRandom, Patrick G; Cadariu, Arina; Concato, John The health-related effects of alcohol use in older persons: a systematic review. Substance abuse,2002;23(3):149-64 | Review purpose not looking at falls |
| 311 | Remelli, Francesca; Ceresini, Maria Giorgia; Trevisan, Caterina; Noale, Marianna; Volpato, Stefano Prevalence and impact of polypharmacy in older patients with type 2 diabetes Aging Clinical and Experimental Research,2022;34(9):1969-1983 | Review purpose not looking at falls |
| 312 | Rensink, M; Schuurmans, M; Lindeman, E; HafsteinsdottiRandom, T B [Falls: incidence and risk factors after stroke. A systematic literature review]. Tijdschrift voor gerontologie en geriatrie,2009;40(4):156-67 | Language |
| 313 | Rice, Laura A; Ousley, Cherita; Sosnoff, Jacob J A systematic review of risk factors associated with accidental falls, outcome measures and interventions to manage fall risk in non-ambulatory adults Disability and rehabilitation,2015;37(19):1697-1705 | Samples not community dwelling |
| 314 | Roeing, Kathleen L; Hsieh, Katherine L; Sosnoff, Jacob J A systematic review of balance and fall risk assessments with mobile phone technology. Archives of gerontology and geriatrics,2017;73(8214379, 7ax):222-226 | Review purpose not looking at falls |
| 315 | Romli, M H; Tan, M P; Mackenzie, L; Lovarini, M; Suttanon, P; Clemson, L Falls amongst older people in Southeast Asia: a scoping review. Public health,2017;145(qi7, 0376507):96-112 | Study designs not prospective |
| 316 | Romli, Muhammad Hibatullah; Mackenzie, Lynette; Lovarini, Meryl; Tan, Maw Pin; Clemson, Lindy The Clinimetric Properties of Instruments Measuring Home Hazards for Older People at Risk of Falling: A Systematic Review. Evaluation & the health professions,2018;41(1):82-128 | Study designs not prospective |
| 317 | Rosedale, Mary Catching falls: a synthesis of recent research Caring,2001;20(1):14-19 | Review design |
| 318 | Rossignaud, R.; Oliveira, A. C. P.; Lara, J. P. R.; MayoRandom, J. J. V.; Rodacki, A. L. F. Methodological tools used for tripping gait analysis of elderly and prosthetic limb users: a systematic review Aging Clinical and Experimental Research,2019;(): | Review purpose not looking at falls |
| 319 | Rubenstein, Laurence Z Falls in older people: epidemiology, risk factors and strategies for prevention Age and ageing,2006;35(suppl_2):ii37-ii41 | Review design |
| 320 | Ruggieri, Marcello; Palmisano, Biagio; Fratocchi, Giancarlo; Santilli, Valter; Mollica, Roberta; Berardi, Anna; Galeoto, Giovanni Validated Fall Risk Assessment Tools for Use with Older Adults: A Systematic Review Physical and Occupational Therapy in Geriatrics,2018;36(4):331-353 | Study designs not prospective |
| 321 | Ruxton, Drugs with anticholinergic effects and cognitive impairment, falls and all-cause mortality in older adults: a systematic review and meta-analysis (vol 80, pg 209, 2015) British Journal of Clinical Pharmacology,2015;80(4):921-926 | Study designs not prospective |
| 322 | Ruxton, Kimberley; Woodman, Richard J; Mangoni, Arduino A Drugs with anticholinergic effects and cognitive impairment, falls and all-cause mortality in older adults: A systematic review and meta-analysis. British journal of clinical pharmacology,2015;80(2):209-20 | Other |
| 323 | Rydwik, Elisabeth; Bergland, Astrid; Fors√©n, Lisa; Fr√§ndin, Kerstin Psychometric Properties of Timed Up and Go in Elderly People: A Systematic Review Physical and Occupational Therapy in Geriatrics,2011;29(2):102-125 | Review purpose not looking at falls |
| 324 | Salari, Nader; Darvishi, Niloofar; Ahmadipanah, Melika; Shohaimi, Shamarina; Mohammadi, Masoud Global prevalence of falls in the older adults: a comprehensive systematic review and meta-analysis Journal of Orthopaedic Surgery and Research,2022;17(1):334 | Study designs not prospective |
| 325 | Sanchez-Riera, L; Carnahan, E; Vos, T; Veerman, L; Norman, R; Lim, S S; Hoy, D; Smith, E; Wilson, N; Nolla, J M; Chen, J S; Macara, M; Kamalaraj, N; Li, Y; Kok, C; Santos-Hernandez, C; March, L The global burden attributable to low bone mineral density. Annals of the rheumatic diseases,2014;73(9):1635-45 | Review purpose not looking at falls |
| 326 | Santos, Silvana Sidney Costa; da Silva, Mar√≠lia Egues; de Pinho, Leandro Barbosa; Gaut√©rio, Daiane Porto; PelzeRandom, Marlene Teda; da Silveira, Rosemary Silva Risk of falls in the elderly: an integrative review based on the North American Nursing Diagnosis Association. Revista da Escola de Enfermagem da USP,2012;46(5):1227-1236 | Review design |
| 327 | SattaRandom, S; Haase, K; KusteRandom, S; Puts, M; Spoelstra, S; Bradley, C; Wildes, T M; Alibhai, S Falls in older adults with cancer: an updated systematic review of prevalence, injurious falls, and impact on cancer treatment. Supportive care in cancer : official journal of the Multinational Association of Supportive Care in CanceRandom,2021;29(1):21-33 | Samples not community dwelling |
| 328 | SattaRandom, Schroder; Alibhai, Shabbir M H; Spoelstra, Sandra L; Fazelzad, Rouhi; Puts, Martine T E Falls in older adults with cancer: a systematic review of prevalence, injurious falls, and impact on cancer treatment. Supportive care in cancer : official journal of the Multinational Association of Supportive Care in CanceRandom,2016;24(10):4459-69 | Samples not community dwelling |
| 329 | Sattin, Richard W Falls among older persons: a public health perspective Annual review of public health,1992;13(1):489-508 | Review design |
| 330 | ScharneRandom, Vincenz; HasiebeRandom, Lukas; S√∂nnichsen, Andreas; Mann, Eva Efficacy and safety of Z-substances in the management of insomnia in older adults: a systematic review for the development of recommendations to reduce potentially inappropriate prescribing BMC Geriatrics,2022;22(1):87-87 | Samples not community dwelling |
| 331 | ScheffeRandom, Alice C; Schuurmans, Marieke J; van Dijk, Nynke; van der Hooft, Truus; de Rooij, Sophia E Fear of falling: measurement strategy, prevalence, risk factors and consequences among older persons. Age and ageing,2008;37(1):19-24 | Review purpose not looking at falls |
| 332 | Schepers, Paul; den BrinkeRandom, Berry; Methorst, Rob; Helbich, Marco Pedestrian falls: A review of the literature and future research directions. Journal of safety research,2017;62(1264241):227-234 | Samples not community dwelling |
| 333 | Schoene, Daniel; Wu, Sandy M-S; Mikolaizak, A Stefanie; Menant, Jasmine C; Smith, Stuart T; Delbaere, Kim; Lord, Stephen R Discriminative ability and predictive validity of the timed up and go test in identifying older people who fall: systematic review and meta-analysis. Journal of the American Geriatrics Society,2013;61(2):202-8 | Study designs not prospective |
| 334 | Schoot, Tessa S.; Goto, Namiko A.; van Marum, Rob J.; Hilbrands, Luuk B.; Kerckhoffs, Angele P. M. Dialysis or kidney transplantation in older adults? A systematic review summarizing functional, psychological, and quality of life-related outcomes after start of kidney replacement therapy International urology and nephrology,2022;54(11):2891-2900 | Study designs not prospective |
| 335 | Schulein, S [Comparison of the performance-oriented mobility assessment and the Berg balance scale. Assessment tools in geriatrics and geriatric rehabilitation]. Zeitschrift fur Gerontologie und Geriatrie,2014;47(2):153-64 | Language |
| 336 | Scott, V.; Metcalfe, S.; Yassin, Y. FALL AND FALL-RELATED INJURY STUDIES AMONG OLDER ABORIGINAL PEOPLE IN AUSTRALIA, CANADA, NEW ZEALAND AND THE USA: A SYSTEMATIC REVIEW Injury Prevention,2015;21():A123-A123 | Other |
| 337 | Scott, Vicky; Votova, Kristine; Scanlan, Andria; Close, Jacqueline Multifactorial and functional mobility assessment tools for fall risk among older adults in community, home-support, long-term and acute care settings. Age and ageing,2007;36(2):130-9 | Review purpose not looking at falls |
| 338 | Seaman, Karla; Ludlow, Kristiana; Wabe, Nasir; Dodds, Laura; Siette, Joyce; Nguyen, Amy; Jorgensen, Mikaela; Lord, Stephen R.; Close, Jacqueline C. T.; O'Toole, Libby; Lin, Caroline; Eymael, Annaliese; Westbrook, Johanna The use of predictive fall models for older adults receiving aged care, using routinely collected electronic health record data: a systematic review BMC Geriatrics,2022;22(1):45304 | Samples not community dwelling |
| 339 | Seidu, Samuel; KunutsoRandom, Setor K; TopseveRandom, Pinar; Hambling, Clare E; Cos, Francesc X; Khunti, Kamlesh Deintensification in older patients with type 2 diabetes: A systematic review of approaches, rates and outcomes. Diabetes, obesity & metabolism,2019;21(7):1668-1679 | Review purpose not looking at falls |
| 340 | Seppala, Lotta J; Wermelink, Anne M A T; de Vries, Max; Ploegmakers, Kimberley J; van de Glind, Esther M M; Daams, Joost G; van der Velde, Nathalie; EUGMS task and Finish group on fall-risk-increasing drugs Fall-Risk-Increasing Drugs: A Systematic Review and Meta-Analysis: II. Psychotropics. Journal of the American Medical Directors Association,2018;19(4):371.e11-371.e17 | Samples not community dwelling |
| 341 | Shafizadeh, Mohsen; Manson, Jane; Fowler-Davis, Sally; Ali, Khalid; Lowe, Anna C; Stevenson, Judy; ParvinpouRandom, Shahab; Davids, Keith Effects of Enriched Physical Activity Environments on Balance and Fall Prevention in Older Adults: A Scoping Review. Journal of aging and physical activity,2020;(9415639):45305 | Review purpose not looking at falls |
| 342 | Silpakampises, Kanokwan; Suwanwaha, Sakkarin Risk Factors Affecting Falls among Old Adults in Community: A Systematic Review KASEM BUNDIT JOURNAL,2019;20(Feburary):23-38 | Study designs not prospective |
| 343 | Silva de Souza Moreira, Ana Carolina; Zarpellon Mazo, Giovana; HauseRandom, Eduardo; Bertolini de Paiva, Paula; Menezes, Enaiane; Cardoso, Fernando Luiz Do functional mobility tests predict the risk of falls in community-dwelling elderly? Manual Therapy, Posturology & Rehabilitation Journal,2016;14():45305 | Study designs not prospective |
| 344 | Silva Fernandes, Viviane Lemos; Martins Ribeiro, Darlan; Caetano Fernandes, Luciana; Losada de Menezes, Ruth Postural changes versus balance control and falls in community-living older adults: a systematic review. Fisioterapia em Movimento,2018;31(1):45306 | Study designs not prospective |
| 345 | Silva Gama, Zenewton Andre da; Gomez Conesa, Antonia; Sobral Ferreira, Marta [Epidemiology of falls in the elderly in Spain: a systematic review, 2007]. Revista espanola de salud publica,2008;82(1):43-55 | Language |
| 346 | Sirkin, Amy J; RosneRandom, Noel G Hypertensive management in the elderly patient at risk for falls. Journal of the American Academy of Nurse Practitioners,2009;21(7):402-8 | Review design |
| 347 | Snowden, Mark B; Steinman, Lesley E; Bryant, Lucinda L; CherrieRandom, Monique M; Greenlund, Kurt J; Leith, Katherine H; Levy, Cari; Logsdon, Rebecca G; Copeland, Catherine; Vogel, Mia; Anderson, Lynda A; Atkins, David C; Bell, Janice F; Fitzpatrick, Annette L Dementia and co-occurring chronic conditions: a systematic literature review to identify what is known and where are the gaps in the evidence?. International journal of geriatric psychiatry,2017;32(4):357-371 | Review purpose not looking at falls |
| 348 | Soares, Wuber J S; Lopes, Alexandre D; Nogueira, Eduardo; Candido, Victor; de Moraes, Suzana A; Perracini, Monica R Physical Activity Level and Risk of Falling in Community-Dwelling Older Adults: Systematic Review and Meta-Analysis. Journal of aging and physical activity,2018;(9415639):45301 | Other |
| 349 | Sobieraj, Diana M; Martinez, Brandon K; Hernandez, Adrian V; Coleman, Craig I; Ross, Joseph S; Berg, Karina M; Steffens, David C; BakeRandom, William L Adverse Effects of Pharmacologic Treatments of Major Depression in Older Adults. Journal of the American Geriatrics Society,2019;67(8):1571-1581 | Study designs not prospective |
| 350 | Sousa, Luis Manuel Mota; Marques-Vieira, Cristina Maria Alves; Caldevilla, Maria Nilza Guimaraes Nogueira de; Henriques, Cristina Maria Alves Dias; Severino, Sandy Silva Pedro; Caldeira, Silvia Maria Alves Risk for falls among community-dwelling older people: systematic literature review. Revista gaucha de enfermagem,2017;37(4):e55030 | Other |
| 351 | Stalenhoef, Paul A.; CreboldeRandom, Harry F.J.M.; Knottnerus, J. Andre; Van Der Horst, Frans G.E.M. Incidence, risk factors and consequences of falls among elderly subjects living in the community: a criteria-based analysis European Journal of Public Health,1997;7():328-334 | Study designs not prospective |
| 352 | Stasny, Bernadette Marie; Newton, Roberta A.; Viggiano LoCascio, Lara; Bedio, Natalie; Lauke, Christine; Conroy, Margaret; Thompson, Allison; Vakhnenko, Lyudmila; Polidoro, Christine The ABC Scale and Fall Risk: A Systematic Review Physical and Occupational Therapy in Geriatrics,2011;29(3):233-242 | Study designs not prospective |
| 353 | Stewart, C.; Taylor-Rowan, M.; Soiza, R. L.; Quinn, T. J.; Loke, Y. K.; Myint, P. K. Anticholinergic burden measures and older people's falls risk: a systematic prognostic review Therapeutic Advances in Drug Safety,2021;12(): | Samples not community dwelling |
| 354 | Strini, Veronica; Schiavolin, Roberta; Prendin, Angela Fall Risk Assessment Scales: A Systematic Literature Review Nursing Reports,2021;11(2):430-443 | Review purpose not looking at falls |
| 355 | Stubbs, Brendon; Stubbs, Jean; Gnanaraj, Solomon Donald; Soundy, Andrew Falls in older adults with major depressive disorder (MDD): a systematic review and exploratory meta-analysis of prospective studies. International psychogeriatrics,2016;28(1):45558 | Samples not community dwelling |
| 356 | Stuck, A. K.; Basile, G.; FreystaetteRandom, G.; de Godoi Rezende Costa Molino, C.; Lang, W.; Bischoff-Ferrari, H. A. Predictive validity of current sarcopenia definitions (EWGSOP2, SDOC, and AWGS2) for clinical outcomes: A scoping review Journal of Cachexia, Sarcopenia and Muscle,2023;14(1):71-83 | Review purpose not looking at falls |
| 357 | Su, Y. C.; Chang, S. F.; Tsai, H. C. The Relationship between Sarcopenia and Injury Events: A Systematic Review and Meta-Analysis of 98,754 Older Adults Journal of Clinical Medicine,2022;11(21):6474 | Samples not community dwelling |
| 358 | Suh, Kevin; Beck, Jordan; Katzman, Wendy; Allen, Diane D Homelessness and rates of physical dysfunctions characteristic of premature geriatric syndromes: systematic review and meta-analysis. Physiotherapy theory and practice,2020;(9015520):45301 | Age not mean of 60 years + |
| 359 | Suh, Kevin; Beck, Jordan; Katzman, Wendy; Allen, Diane D. Homelessness and rates of physical dysfunctions characteristic of premature geriatric syndromes: systematic review and meta-analysis Physiotherapy Theory & Practice,2022;38(7):858-867 | Age not mean of 60 years + |
| 360 | SunYoung Hong; Heeok Park A Meta-analysis of the Risk Factors related to Falls among Elderly Patients with Dementia. Korean Journal of Adult Nursing,2017;29(1):51-62 | Language |
| 361 | Szabo, Shelagh M; Gooch, Katherine L; WalkeRandom, David R; Johnston, Karissa M; Wagg, Adrian S The Association Between Overactive Bladder and Falls and Fractures: A Systematic Review. Advances in therapy,2018;35(11):1831-1841 | Study designs not prospective |
| 362 | TanwaRandom, Rohit; Nandal, Neha; Zamani, Mazdak; Manaf, Azizah Abdul Pathway of Trends and Technologies in Fall Detection: A Systematic Review Healthcare (Basel, Switzerland),2022;10(1): | Review purpose not looking at falls |
| 363 | Teng, Z.; Zhu, Y.; Teng, Y.; Long, Q.; Hao, Q.; Yu, X.; Yang, L.; Lv, Y.; Liu, J.; Zeng, Y.; Lu, S. The analysis of osteosarcopenia as a risk factor for fractures, mortality, and falls Osteoporosis International,2021;32(11):2173-2183 | Samples not community dwelling |
| 364 | Thibaud, M.; Bloch, F.; Tournoux-Facon, C.; Breque, C.; Rigaud, A. S.; Dugue, B.; Kemoun, G. Impact of physical activity and sedentary behaviour on fall risks in older people: a systematic review and meta-analysis of observational studies European Review of Aging and Physical Activity,2012;9(1):45427 | Study designs not prospective |
| 365 | Thornby, Maureen A. Balance and falls in the frail older person: a review of the literature Topics in Geriatric Rehabilitation,1995;11(2):35-43 | Review design |
| 366 | Tolentino, Arthur; Amaral, Savio; S Souza, Lucca; Zeballos, Diana; Brites, Carlos Frequency of Falls and Associated Risk Factors in People Living With HIV: A Systematic Review With Meta-Analysis and Meta-Regression Journal of acquired immune deficiency syndromes (1999),2021;86(5):616-625 | Age not mean of 60 years + |
| 367 | Tolentino, Arthur; Amaral, Savio; Souza, Lucca S; Zeballos, Diana; Brites, Carlos Frequency of falls and associated risk factors in people living with HIV: a systematic review with meta-analysis and meta-regression. Journal of acquired immune deficiency syndromes (1999),2020;Publish Ahead of Print(100892005): | Age not mean of 60 years + |
| 368 | Torbahn, G; Strauss, T; SiebeRandom, C C; KiesswetteRandom, E; Volkert, D Nutritional status according to the mini nutritional assessment (MNA) R as potential prognostic factor for health and treatment outcomes in patients with cancer - a systematic review. BMC canceRandom,2020;20(1):594 | Review purpose not looking at falls |
| 369 | Torbahn, G.; Strauss, T.; SiebeRandom, C. C.; KiesswetteRandom, E.; Volkert, D. Nutritional status according to the Mini Nutritional Assessment (MNA) as potential prognostic factor for health and treatment outcomes in cancer patients: A systematic review European Geriatric Medicine,2019;10(Supplement 1):S294 | Review purpose not looking at falls |
| 370 | Torbahn, G.; Strauss, T.; SiebeRandom, C. C.; KiesswetteRandom, E.; Volkert, D. Nutritional status according to the mini nutritional assessment (MNA)¬Æ as potential prognostic factor for health and treatment outcomes in patients with cancer - a systematic review. BMC CanceRandom,2020;20(1):45309 | Other |
| 371 | Treves, Nir; Perlman, Amichai; Kolenberg Geron, Lital; Asaly, Angham; Matok, Ilan Z-drugs and risk for falls and fractures in older adults-a systematic review and meta-analysis. Age and ageing,2018;47(2):201-208 | Samples not community dwelling |
| 372 | Tuena, C.; Borghesi, F.; Bruni, F.; Cavedoni, S.; Maestri, S.; Riva, G.; Tettamanti, M.; Liperoti, R.; Rossi, L.; Ferrarin, M.; Stramba-Badiale, M. Technology-Assisted Cognitive Motor Dual-Task Rehabilitation in Chronic Age-Related Conditions: Systematic Review Journal of Medical Internet Research,2023;25():e44484 | Review purpose not looking at falls |
| 373 | Tuira, M.; Martins, L.; Barroso, M. P.; Arezas, P.; Baptista, J. S.; Carneiro, P.; Cordeiro, P.; Costa, N.; Melo, R.; Miguel, A. S.; Perestrelo, G. Environmental factors and household accidents in the elderly: a systematic review Sho2019: International Symposium on Occupational Safety and Hygiene,2019;():92-94 | Other |
| 374 | Tully-Wilson, C.; Bojack, R.; MilleaRandom, P. M.; Stallman, H. M.; Allen, A.; Mason, J. Self-Perceptions of Aging: A Systematic Review of Longitudinal Studies Psychology and Aging,2021;36(7):773-789 | Review purpose not looking at falls |
| 375 | Vaishya, Raju; Vaish, Abhishek Falls in Older Adults are Serious. Indian Journal of Orthopaedics,2020;54(1):69-74 | Review design |
| 376 | ValipooRandom, S.; Pati, D.; Kazem-Zadeh, M.; Mihandoust, S.; Mohammadigorji, S. Falls in Older Adults: A Systematic Review of Literature on Interior-Scale Elements of the Built Environment Journal of Housing for the Elderly,2020;34(4):351-374 | Study designs not prospective |
| 377 | Van Camp, L; DejaegeRandom, M; Tournoy, J; Gielen, E; Laurent, M R Association of orthogeriatric care models with evaluation and treatment of osteoporosis: a systematic review and meta-analysis. Osteoporosis international : a journal established as result of cooperation between the European Foundation for Osteoporosis and the National Osteoporosis Foundation of the USA,2020;31(11):2083-2092 | Review purpose not looking at falls |
| 378 | Varela-Burstein, Ellyn; MilleRandom, Patricia A. Is chronic pain a risk factor for falls among community dwelling elders? Topics in Geriatric Rehabilitation,2003;19(2):145-159 | Review design |
| 379 | Venegas Sanabria, Luis Carlos; Barbosa Balaquera, Stephany; Suarez Acosta, Ana Maria; Garcia Pena, Angel Alberto; Cano Gutierrez, Carlos Alberto [Statin and risk of falls in the elderly: A sytematic review of the literature]. Revista espanola de geriatria y gerontologia,2017;52(6):317-321 | Language |
| 380 | Verma, Sarita; Pickett, William Falls in the elderly Geriatrics Today,2001;41(1):28-31 | Review design |
| 381 | Vermeiren, Sofie; Vella-Azzopardi, Roberta; Beckwee, David; Habbig, Ann-Katrin; Scafoglieri, Aldo; Jansen, Bart; Bautmans, Ivan; Gerontopole Brussels Study group Frailty and the Prediction of Negative Health Outcomes: A Meta-Analysis. Journal of the American Medical Directors Association,2016;17(12):1163.e1-1163.e17 | Review purpose not looking at falls |
| 382 | Vernet, N.; Mouchoux, C. Impact of the potentially inappropriate prescribing among the elderly: A literature review Journal de Pharmacie Clinique,2019;38(1):45496 | Language |
| 383 | Vieira, E. R.; PalmeRandom, R. C.; Chaves, P. H. M. Prevention of falls in older people living in the community Bmj-British Medical Journal,2016;353(): | Review design |
| 384 | Virnes, Roosa-Emilia; Tiihonen, Miia; Karttunen, Niina; van Poelgeest, Eveline P.; van der Velde, Natalie; Hartikainen, Sirpa Opioids and Falls Risk in Older Adults: A Narrative Review Drugs & Aging,2022;39(3):199-207 | Review design |
| 385 | Visschedijk, Jan; Achterberg, Wilco; van Balen, Romke; Hertogh, Cees Fear of Falling After Hip Fracture: A Systematic Review of Measurement Instruments, Prevalence, Interventions, and Related Factors Journal of the American Geriatrics Society,2010;58(9):1739-1748 | Review purpose not looking at falls |
| 386 | WANG Han; WANG Shasha; HU Shujing; GUO Xuejie; HAN Mengxi; LIU Hongxia Association between failty and adverse outcomes in patients with chronic kidney disease: a systematic review. Chinese Nursing Research,2020;34(9):1497-1505 | Age not mean of 60 years + |
| 387 | Watt, Andrew A.; Clark, Carol; Williams, Jonathan M. Differences in sit-to-stand, standing sway and stairs between community-dwelling fallers and non-fallers: a review of the literature. Physical Therapy Reviews,2018;23(45387):273-290 | Study designs not prospective |
| 388 | WechsleRandom, Stephen; Wood, Lisa The Effect of Chemotherapy on Balance, Gait, and Falls Among Cancer Survivors: A Scoping Review. Rehabilitation Oncology,2021;39(1):45465 | Age not mean of 60 years + |
| 389 | Wehner-Hewson, N.; Watts, P.; Buscombe, R.; Bourne, N.; Hewson, D. Racial and Ethnic Differences in Falls Among Older Adults: a Systematic Review and Meta-analysis Journal of racial and ethnic health disparities,2022;9(6):2427-2440 | Study designs not prospective |
| 390 | Weiss, Jessica; Freeman, Michele; Low, Allison; Fu, Rochelle; Kerfoot, Amy; PaynteRandom, Robin; Motu'apuaka, Makalapua; Kondo, Karli; Kansagara, Devan Benefits and Harms of Intensive Blood Pressure Treatment in Adults Aged 60 Years or Older: A Systematic Review and Meta-analysis. Annals of internal medicine,2017;166(6):419-429 | Study designs not prospective |
| 391 | Welsh, Victoria K; Clarson, Lorna E; Mallen, Christian D; McBeth, John Multisite pain and SRed falls in older people: systematic review and meta-analysis. Arthritis research & therapy,2019;21(1):67 | Study designs not prospective |
| 392 | Whelan, Greg Alcohol: a much neglected risk factor in elderly mental disorders Current Opinion in Psychiatry,2003;16(6):609-614 | Review design |
| 393 | Wildes, Tanya M; Dua, Priya; FowleRandom, Susan A; MilleRandom, J Philip; CarpenteRandom, Christopher R; Avidan, Michael S; Stark, Susan Systematic review of falls in older adults with cancer. Journal of geriatric oncology,2015;6(1):70-83 | Study designs not prospective |
| 394 | Willgoss, Thomas G; Yohannes, Abebaw M; Mitchell, Duncan Review of risk factors and preventative strategies for fall-related injuries in people with intellectual disabilities. Journal of clinical nursing,2010;19(15-16):73294 | Age not mean of 60 years + |
| 395 | Wiseman, Taneal; Betihavas, Vasiliki The association between unexplained falls and cardiac arrhythmias: A scoping literature review. Australian critical care : official journal of the Confederation of Australian Critical Care Nurses,2019;32(5):434-441 | Age not mean of 60 years + |
| 396 | Wong, A. Y.; Lo, C.; Tsang, W.; Yan, C.; Stephen, L.; Hill, K. A systematic review on risk factors for falls in patients after total hip/knee arthroplasty Osteoarthritis and Cartilage,2019;27(Supplement 1):S450-S451 | Other |
| 397 | Wong, M. Y. C.; Ou, K. L.; Chung, P. K.; Chui, K. Y. K.; Zhang, C. Q. The relationship between physical activity, physical health, and mental health among older Chinese adults: A scoping review Frontiers in public health,2022;10():914548 | Review purpose not looking at falls |
| 398 | Woolcott, John C; Richardson, Kathryn J; Wiens, Matthew O; Patel, Bhavini; Marin, Judith; Khan, Karim M; Marra, Carlo A Meta-analysis of the impact of 9 medication classes on falls in elderly persons. Archives of internal medicine,2009;169(21):1952-60 | Study designs not prospective |
| 399 | Xie, C.; Zhu, M.; Hu, Y. Risk stratification for REM sleep behavior disorder in patients with Parkinson's disease: A PRISMA-compliant meta-analysis and systematic review Clinical Neurology and Neurosurgery,2021;202():106484 | Samples not community dwelling |
| 400 | Xu, Q.; Ou, X.; Li, J. The risk of falls among the aging population: A systematic review and meta-analysis Frontiers in public health,2022;10():902599 | Samples not community dwelling |
| 401 | Xu, X. J.; Tan, M. P. Anticholinergics and falls in older adults Expert Review of Clinical Pharmacology,2022;15(3):285-294 | Review design |
| 402 | Yadav, K.; Eagles, D.; Perry, J. J.; Emond, M. Mobility assessments of geriatric emergency department patients: A systematic review Academic Emergency Medicine,2017;24(Supplement 1):S275 | Other |
| 403 | Yang, L; Liao, L R; Lam, F M H; He, C Q; Pang, M Y C Psychometric properties of dual-task balance assessments for older adults: a systematic review. Maturitas,2015;80(4):359-69 | Review purpose not looking at falls |
| 404 | Yang, Yi; Wang, Kun; Liu, Hengxu; Qu, Jiawei; Wang, Yan; Chen, Peijie; Zhang, TingRan; Luo, Jiong The impact of Otago exercise programme on the prevention of falls in older adult: A systematic review Frontiers in public health,2022;10():953593 | Review purpose not looking at falls |
| 405 | Yeung, S.; Reijnierse, E.; Trappenburg, M.; Meskers, C.; MaieRandom, A. B. Sarcopenia and its association with falls and fractures in older adults: A systematic review and meta-analysis European Geriatric Medicine,2018;9(Supplement 1):S97 | Other |
| 406 | Yeung, Suey S Y; Reijnierse, Esmee M; Pham, Vivien K; Trappenburg, Marijke C; Lim, Wen Kwang; Meskers, Carel G M; MaieRandom, Andrea B Sarcopenia and its association with falls and fractures in older adults: A systematic review and meta-analysis. Journal of cachexia, sarcopenia and muscle,2019;10(3):485-500 | Samples not community dwelling |
| 407 | Yoshikawa, Aya; Ramirez, Gilbert; Smith, Matthew Lee; FosteRandom, Margaret; Nabil, Anas K; Jani, Sagar N; Ory, Marcia G Opioid Use and the Risk of Falls, Fall Injuries and Fractures among Older Adults: A Systematic Review and Meta-Analysis. The journals of gerontology. Series A, Biological sciences and medical sciences,2020;75(10):1989-1995 | Samples not community dwelling |
| 408 | YU YANG; XINHUA HU; QIANG ZHANG; RUI ZOU Diabetes mellitus and risk of falls in older adults: a systematic review and meta-analysis Age and Ageing,2016;45(6):761-767 | Other |
| 409 | Zang, Guiming Antihypertensive drugs and the risk of fall injuries: a systematic review and meta-analysis. The Journal of international medical research,2013;41(5):1408-17 | Samples not community dwelling |
| 410 | Zhang, Jun; Wu, Yifei; Sharma, Bhavna; Gupta, Ritu; Jawla, Shantanu; Bullimore, Mark A Epidemiology and burden of astigmatism: a systematic literature review Optometry and Vision Science,2023;100(3):218 | Age not mean of 60 years + |
| 411 | Zhang, Sulin; Xu, Wenchao; Zhu, Yuting; Tian, E; Kong, Weijia Impaired Multisensory Integration Predisposes the Elderly People to Fall: A Systematic Review. Frontiers in neuroscience,2020;14(101478481):411 | Review purpose not looking at falls |
| 412 | Zhang, Xiao Ming; Wu, Xin Juan; Cao, J.; Jiao, J.; Chen, W. Association between Cognitive Frailty and Adverse Outcomes among Older Adults: A Meta-Analysis Journal of Nutrition, Health & Aging,2022;26(9):817-825 | Samples not community dwelling |
| 413 | Zheng, Jacqueline J J; Delbaere, Kim; Close, Jacqueline C T; Sachdev, Perminder S; Lord, Stephen R Impact of white matter lesions on physical functioning and fall risk in older people: a systematic review. Stroke,2011;42(7):2086-90 | Study designs not prospective |
| 414 | Zhong, Runting; Rau, Pei-Luen Patrick Are cost-effective technologies feasible to measure gait in older adults? A systematic review of evidence-based literature. Archives of gerontology and geriatrics,2020;87(8214379, 7ax):103970 | Review purpose not looking at falls |
| 415 | Zhou, Jia; Chang, Hui; Leng, Minmin; Wang, Zhiwen Intrinsic Capacity to Predict Future Adverse Health Outcomes in Older Adults: A Scoping Review Healthcare (2227-9032),2023;11(4):450 | Samples not community dwelling |
| 416 | Zhou, Q.; He, J.; Yang, X.; Yin, H.; Zhang, Z.; He, N. The association between physical frailty and injurious falls and all-cause mortality as negative health outcomes in people living with HIV: A systematic review and meta-analysis International Journal of Infectious Diseases,2023;126():193-199 | Age not mean of 60 years + |
| 417 | Zhou, Yuanjin; StrayeRandom, Alisa T.; Phelan, Elizabeth A.; Sadak, Tatiana; Hooyman, Nancy R. A mixed methods systematic review of informal caregivers' experiences of fall risk among community‚Äêdwelling elders with dementia Health & Social Care in the Community,2021;29(4):1126-1144 | Review purpose not looking at falls |
| 418 | Zhu, Hongli; Hu, Kun; Liu, Shiyong; Kim, Ho-Cheol; Wang, Youfa; Xue, Qianli Systematic causality mapping of factors leading to accidental falls of older adults Public health in practice (Oxford, England),2020;1():100045 | Study designs not prospective |
| 419 | Zijlstra, A; Ufkes, T; Skelton, Dawn A; Lundin-Olsson, Lillemor; Zijlstra, W Do dual tasks have an added value over single tasks for balance assessment in fall prevention programs? A mini-review Gerontology,2008;54(1):40-49 | Samples not community dwelling |
| 420 | Gates S, Smith LA, Fisher JD, Lamb SE. Systematic review of accuracy of screening instruments for predicting fall risk among independently living older adults. Database of Abstracts of Reviews of Effects (DARE): Quality-assessed Reviews [Internet]. 2008. | Study designs not prospective |

#### Table S1.2. Risk of Bias – ROBIS Assessment

| **Review Author** | **Phase 2: By Domains** | | | | **Phase 3: Judging Risk of Bias** |
| --- | --- | --- | --- | --- | --- |
|  | **Domain 1: Study Eligibility** | **Domain 2: Identification & Selection of Studies** | **Domain 3: Data Collection & Study Appraisal** | **Domain 4: Synthesis & Findings** |  |
| Ang et al., (2018) | low | low | low | unclear | low |
| Barry et al., (2014) | low | low | low | low | low |
| Blodgett et al., (2022); | high | low | low | high | low |
| Chantanachai et al., (2021) | low | low | low | low | low |
| Cheng et al., (2017) | high | high | high | low | high |
| Chu et al., (2013) | high | high | low | high | high |
| Deandrea et al., (2010) | high | Low | high | Low | low |
| Dibello et al., (2023) | high | high | low | high | high |
| Fernando et al., (2017) | low | low | high | high | High |
| Gambaro et al., (2022) | low | low | low | low | low |
| Gafner et al., (2021) | low | low | low | high | low |
| Ganz et al., (2007) | high | high | high | low | high |
| Gillain et al., (2018) | high | low | low | high | high |
| Gravesande et al., (2017) | low | unclear | low | high | low |
| Hartog et al., (2017) | low | high | high | low | low |
| Iseli et al., (2020) | low | high | high | high | high |
| Jehu et al., (2020) | low | low | low | low | low |
| Kearney et al., (2013) | low | high | high | high | high |
| Kojima (2015) | low | high | high | low | low |
| kvelde et al., (2013) | high | high | high | high | high |
| Launay et al., (2012) | high | high | high | high | High |
| Leroy et al., (2023) | low | high | low | high | low |
| Letts et al., (2010) | high | high | low | high | high |
| Lima et al., (2018) | low | low | low | low | low |
| Lusardi et al., (2017) | low | high | low | high | high |
| Malik et al., (2020) | high | low | low | low | low |
| Manlapaz et al., (2019) | low | low | low | unclear | High |
| Marin et al., (2022) | low | high | low | low | high |
| Meekes et al., (2021) | high | high | low | high | high |
| Menz et al., (2018) | high | high | high | high | high |
| Ming et al., (2018) | high | high | low | low | high |
| Modaressi et al., (2019) | low | low | low | low | low |
| Moreland et al., (2004) | high | low | low | high | low |
| Muir et al., (2010) | low | high | low | low | low |
| Muir et al., (2012) | low | low | low | unclear | low |
| Muir-Hunter et al., (2016) | low | low | low | high | high |
| Mullin et al., (2021) | low | high | low | low | low |
| Okubo et al., (2021) | low | high | low | low | low |
| Õmana et al., (2021 | high | high | low | low | low |
| Pesonen et al., (2020) | low | low | high | high | low |
| Petersen et al., (2020) | high | high | high | high | high |
| Piirtola et al., (2006) | low | high | high | high | high |
| Power et al., (2014) | high | high | low | high | high |
| Qian et al., (2021) | low | high | low | low | low |
| Quijoux et al., (2020) | low | low | high | low | low |
| Ramsey et al., (2022) | high | low | low | low | low |
| Rosa et al., (2019) | low | low | high | high | high |
| Salonen et al., (2012) | high | high | high | high | high |
| Seppala et al., (2018) | low | low | low | high | low |
| Sharma et al., (2023) | low | high | low | low | low |
| Soares et al., (2019) | low | low | low | low | low |
| Stubbs (2014b) | low | high | low | low | high |
| Stubbs et al., (2014a) | low | high | low | low | low |
| Trevisan et al., (2019) | low | high | low | low | low |
| Xu et al., (2018) | low | low | low | low | low |
| Yang et al., (2016) | low | high | high | high | high |
| Zhang et al., (2020) | low | high | low | low | high |

#### Table S1.3. Summary of All Relationships Predicting Any Fall Risk

| **Factor** | **# of Reviews** | **Total # of Relationships** | **# of Relationships Significant for Predicting Fall Risk (reduced or increased risk)** | **% Relationships that were Significant** |
| --- | --- | --- | --- | --- |
| Age | 6 | 8 | 4 | 50% |
| Balance | 14 | 142 | 65 | 46% |
| Body composition | 3 | 13 | 4 | 31% |
| Chronic conditions | 16 | 42 | 35 | 83% |
| Cognition | 12 | 80 | 46 | 58% |
| Disability | 8 | 11 | 9 | 82% |
| Dual task ability | 6 | 19 | 15 | 79% |
| Education | 1 | 1 | 1 | 100% |
| Environment | 1 | 2 | 2 | 100% |
| Fall-specific factors | 6 | 9 | 9 | 100% |
| Foot problems | 3 | 3 | 2 | 67% |
| Frailty | 3 | 17 | 17 | 100% |
| Gait | 10 | 57 | 39 | 68% |
| Gender | 5 | 5 | 3 | 60% |
| Health status | 3 | 3 | 3 | 100% |
| Medications | 9 | 26 | 15 | 58% |
| Physical function | 11 | 38 | 31 | 82% |
| Nutrition | 1 | 2 | 2 | 100% |
| Oral health | 1 | 5 | 5 | 100% |
| Pain | 4 | 5 | 3 | 60% |
| Physical activity | 8 | 32 | 14 | 44% |
| Psychosocial | 9 | 13 | 12 | 92% |
| Range of Motion | 1 | 4 | 2 | 50% |
| Sedentary Time | 1 | 3 | 1 | 33% |
| Sensorimotor | 5 | 13 | 10 | 77% |
| Sensory | 3 | 3 | 3 | 100% |
| Strength | 3 | 15 | 8 | 53% |
| Vision | 5 | 7 | 1 | 14% |
| *Notes.*  This table summarizes the relationships from both the meta-analysis and the narratively reported results via counts and proportions  This table does not include the relationships examining predictive validity  Data does not take into consideration magnitude, quality, or sample sizes | | | | |

#### Table S1.4. Summary of Meta-Analysis Relationships Predicting Any Fall Risk

| **Factor** | **# of Reviews** | **Total # of Relationships** | **# of Relationships predicting increased fall risk** | **# of Relationships predicting null fall risk** | **% of Relationships that were Significant** |
| --- | --- | --- | --- | --- | --- |
| Age | 2 | 2 | 1 | 1 | 50% |
| Balance | 5 | 12 | 11 | 1 | 92% |
| Body composition | 2 | 12 | 3 | 9 | 25% |
| Chronic conditions | 8 | 18 | 11 | 7 | 61% |
| Cognition | 4 | 4 | 3 | 1 | 75% |
| Disability | 3 | 3 | 3 | 0 | 100% |
| Dual task ability | 1 | 1 | 1 | 0 | 100% |
| Education | 1 | 1 | 0 | 1 | 0% |
| Fall-specific factors | 2 | 3 | 3 | 0 | 100% |
| Foot problems | 1 | 1 | 1 | 0 | 100% |
| Frailty | 3 | 17 | 17 | 0 | 100% |
| Gait | 2 | 2 | 1 | 1 | 50% |
| Gender | 2 | 2 | 1 | 1 | 50% |
| Health status | 1 | 1 | 1 | 0 | 100% |
| Medications | 4 | 9 | 5 | 4 | 56% |
| Physical function | 4 | 4 | 3 | 1 | 75% |
| Nutrition | 1 | 2 | 2 | 0 | 100% |
| Pain | 3 | 4 | 2 | 2 | 50% |
| Physical activity | 2 | 2 | 1 | 1 | 50% |
| Psychosocial | 5 | 6 | 5 | 1 | 83% |
| Strength | 2 | 3 | 2 | 1 | 67% |
| Vision | 3 | 3 | 1 | 2 | 33% |
| *Notes.*  This table summarizes the relationships from the meta-analysis results via counts and proportions  This table does not include the relationships examining predictive validity  Data does not take into consideration magnitude, quality, or sample sizes | | | | | |

#### Table S1.5. Summary of Single Relationships Predicting Any Fall

| **Risk Factor** | **# of Reviews** | **Total # of relationships** | **# of Relationships Predicting Increased Risk** | **# of Relationships Predicting Null Risk** | **# of Relationships Predicting Reduced Risk** | **% Relationships that were Significant** |
| --- | --- | --- | --- | --- | --- | --- |
| Age | 4 | 6 | 3 | 3 | 0 | 50% |
| Balance | 9 | 130 | 52 | 76 | 2 | 42% |
| Body composition-skeletal muscle | 1 | 1 | 0 | 0 | 1 | 100% |
| Chronic conditions | 8 | 24 | 23 | 1 | 0 | 96% |
| Cognition | 8 | 76 | 38 | 33 | 5 | 57% |
| Disability | 5 | 8 | 5 | 2 | 1 | 75% |
| Dual task ability | 2 | 18 | 14 | 4 | 0 | 78% |
| Environment | 1 | 2 | 2 | 0 | 0 | 100% |
| Fall-specific factors | 4 | 6 | 6 | 0 | 0 | 100% |
| Foot problems | 2 | 2 | 1 | 1 | 0 | 50% |
| Gait | 8 | 55 | 36 | 17 | 2 | 69% |
| Gender | 3 | 3 | 2 | 1 | 0 | 67% |
| Health status | 2 | 2 | 2 | 0 | 0 | 100% |
| Medications | 5 | 17 | 10 | 7 | 0 | 59% |
| Physical function | 7 | 34 | 26 | 6 | 2 | 82% |
| Oral health | 1 | 5 | 2 | 0 | 3 | 100% |
| Pain | 1 | 1 | 1 | 0 | 0 | 100% |
| Physical activity | 6 | 30 | 3 | 17 | 10 | 43% |
| Psychosocial | 4 | 7 | 6 | 0 | 1 | 100% |
| Range of motion | 1 | 4 | 0 | 2 | 2 | 50% |
| Sedentary time | 1 | 3 | 0 | 2 | 1 | 33% |
| Sensorimotor | 5 | 13 | 10 | 3 | 0 | 77% |
| Sensory | 1 | 3 | 3 | 0 | 0 | 100% |
| Strength | 3 | 12 | 4 | 6 | 2 | 50% |
| Vision | 2 | 4 | 0 | 4 | 0 | 0% |
| *Notes.*  This table summarizes the relationships from the narratively reported results via counts and proportions  This table does not include the relationships examining predictive validity  Data does not take into consideration magnitude, quality, or sample sizes | | | | | | |

#### Table S1.6. Predictive Validity of Tests for Any Falls

| **Tool** | **Review Author** | **Cut point** | **# of primary studies** | **AUC (95%CI)** | **Sensitivity % (95% CI)** | **Specificity % (95% CI)** | **Positive Likelihood Ratio (95% CI)** | **Negative Likelihood Ratio (95% CI)** |
| --- | --- | --- | --- | --- | --- | --- | --- | --- |
| **Berg Balance Scale** | Lima et al., (2018) | <40 score | 1 | ·· | ·· | ·· | 5.19 (NR to NR) | ·· |
|  | Lima et al., (2018) | <40 score | 1 | ·· | ·· | ·· | 2.29 (NR to NR) | ·· |
|  | Lima et al., (2018) | 40-44 score | 1 | ·· | ·· | ·· | 1.56 (NR to NR) | ·· |
|  | Lima et al., (2018) | 40-44 score | 1 | ·· | ·· | ·· | 2.07 (NR to NR) | ·· |
|  | Meekes et al., (2021) | $\leq$ 45 score | 1 | ·· | 25 (16 to 36) | 87 (79 to 92) | ·· | ·· |
|  | Lima et al., (2018) | $\leq$45 score | 1 | ·· | 67 (NR to NR) | 68 (NR to NR) | ·· | ·· |
|  | Lima et al., (2018) | $\leq$ 45 score | 1 | ·· | 25 (NR to NR) | 87 (NR to NR) | ·· | ·· |
|  | Lima et al., (2018) | 45-49 score | 1 | ·· | ·· | ·· | 1.34 (NR to NR) | ·· |
|  | Lima et al., (2018) | 45-49 score | 1 | ·· | ·· | ·· | 0.58 (NR to NR) | ·· |
|  | Meekes et al., (2021) | $\leq$ 48 score | 1 | ·· | 69 (NR to NR) | 0.76 (NR to NR) | ·· | ·· |
|  | Lima et al., (2018) | 50-54 score | 1 | ·· | ·· | ·· | 1.003 (NR to NR) | ·· |
|  | Lima et al., (2018) | 50-54 score | 1 | ·· | ·· | ·· | 0.91 (NR to NR) | ·· |
|  | Meekes et al., (2021) | $\leq$52 score | 1 | 0.47 (NR to NR) | ·· | ·· | ·· | ·· |
|  | Meekes et al., (2021) | $\leq$54 score | 1 | 0.59 (NR to NR | 61 (50 to 72) | 53 (43 to 63) | ·· | ·· |
|  | Lima et al., (2018) | $\leq$54 score | 1 | ·· | 61 (NR to NR) | 53 (NR to NR) | ·· | ·· |
|  | Lima et al., (2018) | $\geq$55 score | 1 | ·· | ·· | ·· | 0.73 (NR to NR) | ·· |
|  | Lima et al., (2018) | $\geq$55 score | 1 | ·· | ·· | ·· | 0.53 (NR to NR) | ·· |
|  | Lima et al., (2018) | NR | 1 | NR but insignificant for predicting falls | ·· | ·· | ·· | ·· |
| **Depression - self report** | Lusardi et al., (2017) | Depression present | 1 | ·· | 27 (16 to 40) | 83 (71 to 91) | 1.6 (0.8 to 3.2) | 0.9 (0.7 to 1.1) |
| **Dynamic Gait Index** | Power et al., (2014) | $\leq$20 score | 1 | ·· | 100 (NR to NR) | 76 (NR to NR) | ·· | ·· |
| **Fall History** | Meekes et al., (2021) | $\geq$1 fall in past year | 1 | ·· | 60 (NR to NR) | 65 (NR to NR) | ·· | ·· |
|  | Meekes et al., (2021) | $\geq$1 fall in past year | 1 | ·· | 39 (NR to NR) | 82 (NR to NR) | ·· | ·· |
|  | Meekes et al., (2021) | $\geq$1 fall in past year | 1 | ·· | 63 (NR to NR) | 77 (NR to NR) | ·· | ·· |
|  | Meekes et al., (2021) | $\geq$1 fall in past year | 1 | ·· | 69 (57 to 78) | 63 (57 to 69) | ·· | ·· |
| **Falls Efficacy Scale International** | Lusardi et al., (2017) | $\geq$24 score | 2 | ·· | 6 (6 to 60) | 60 (56 to 65) | 1.7 (1 to 2.4) | 0.6 (0.1 to 0.2) |
| **Functional Gait Assessment** | Power et al., (2014) | $\leq$20 score | 1 | ·· | 1 (NR to NR) | 83 (NR to NR) | ·· | ·· |
| **Functional Reach Test** | Õmana et al., (2021) | $\leq$14.5 cm | 1 | ·· | 68 (NR to NR) | 79 (NR to NR) | ·· | ·· |
|  | Õmana et al., (2021) | $\leq$18 cm | 1 | ·· | 47 (NR to NR) | 59 (NR to NR) | ·· | ·· |
|  | Õmana et al., (2021) | 8 inches | 1 | ·· | 73 (NR to NR) | 88 (NR to NR) | ·· | ·· |
|  | Õmana et al., (2021) | $\leq$20.32 cm | 1 | ·· | 73 (NR to NR) | 88 (NR to NR) | ·· | ·· |
| **Gait Speed (4m)** | Meekes et al., (2021) | 0.67 m/s | 1 | ·· | 82 (NR to NR) | 71 (NR to NR) | ·· | ·· |
|  | Meekes et al., (2021) | $\geq$12 seconds | 1 | ·· | 1 (NR to NR) | 24 (NR to NR) | ·· | ·· |
|  | Meekes et al., (2021) | $\geq$14 seconds | 1 | ·· | 77 (NR to NR) | 57 (NR to NR) | ·· | ·· |
|  | Meekes et al., (2021) | $\geq$18 seconds | 1 | ·· | 38 (NR to NR) | 85 (NR to NR) | ·· | ·· |
| **Gait variabilit**y | Gillain et al., (2018) | NR | 1 | ·· | ·· | ·· | 0.92 (NR to NR) | ·· |
| **Geriatric Depression Scale** | Lusardi et al., (2017) | $\geq$7 score | 3 | ·· | 27 (22 to 31) | 86 (83 to 88) | 1.9 (1.5 to 2.4) | 0.9 (0.8 to 0.9) |
|  | Lusardi et al., (2017) | $\geq$1 Score | 2 | ·· | 37 (34 to 41) | 72 (70 to 75) | 1.3 (1.2 to 1.5) | 0.9 (0.8 to 0.9) |
| **Pain** | Lusardi et al., (2017) | “significant” | 1 | ·· | 47 (36 to 58) | 74 (67 to 80) | 1.8 (1.3 to 2.5) | 0.7 (0.6 to 0.9) |
| **POMA**-**balance** | Õmana et al., (2021) | $\leq$8 score | 1 | ·· | 8 (NR to NR) | 91 (NR to NR) | ·· | ·· |
|  | Meekes et al., (2021) | $\leq$8 score | 1 | ·· | 8 (NR to NR) | 91 (NR to NR) | ·· | ·· |
|  | Õmana et al., (2021) | $\leq$9 score | 1 | ·· | 23 (NR to NR) | 80 (NR to NR) | ·· | ·· |
|  | Meekes et al., (2021) | $\leq$9 score | 1 | ·· | 23 (NR to NR) | 80 (NR to NR) | ·· | ·· |
|  | Õmana et al., (2021) | $\leq$10 score | 1 | ·· | 62 (NR to NR) | 70 (NR to NR) | ·· | ·· |
|  | Meekes et al., (2021) | $\leq$10 score | 1 | ·· | 62 (NR to NR) | 70 (NR to NR) | ·· | ·· |
|  | Meekes et al., (2021) | 10 score | 1 | ·· | 24 (NR to NR) | 91 (NR to NR) | ·· | ·· |
|  | Meekes et al., (2021) | 10 score | 1 | ·· | 64 (45 to 80) | 66 (53 to 77) | ·· | ·· |
|  | Õmana et al., (2021) | $\leq$12 score | 1 | ·· | 24 (NR to NR) | 91 (NR to NR) | ·· | ·· |
|  | Õmana et al., (2021) | $\leq$12 score | 1 | ·· | 55 (NR to NR) | 97 (NR to NR) | ·· | ·· |
|  | Gafner et al., (2021) | 12 score | 1 | ·· | 21 (NR to NR) | 95 (NR to NR) | 4.2 (NR to NR) | 0.83 (NR to NR) |
|  | Power et al., (2014) | NR | 1 | ·· | 51 (NR to NR) | 1 (NR to NR) | ·· | ·· |
|  | Õmana et al., (2021) | NR | 1 | ·· | 95 (NR to NR) | 16 (NR to NR) | ·· | ·· |
| **POMA-GAIT** | Õmana et al., (2021) | $\leq$8 score | 1 | ·· | 21 (NR to NR) | 95 (NR to NR) | ·· | ·· |
|  | Meekes et al., (2021) | 9 score | 1 | ·· | 21 (NR to NR) | 95 (NR to NR) | ·· | ·· |
|  | Meekes et al., (2021) | 9 score | 1 | ·· | 64 (45 to 80) | 63 (50 to 74) | ·· | ·· |
|  | Õmana et al., (2021) | NR | 1 | ·· | 1 (NR to NR) | 0 (NR to NR) | ·· | ·· |
| **POMA-total** | Õmana et al., (2021) | <30 score | 1 | ·· | 27 (NR to NR) | 83 (NR to NR) | ·· | ·· |
|  | Õmana et al., (2021) | $\leq$33 score | 1 | ·· | 51 (NR to NR) | 74 (NR to NR) | ·· | ·· |
|  | Õmana et al., (2021) | $\leq$36 score | 1 | ·· | 70 (NR to NR) | 52 (NR to NR) | ·· | ·· |
|  | Õmana et al., (2021) | NR | 1 | ·· | 93 (NR to NR) | 11 (NR to NR) | ·· | ·· |
| **Reaction time** | Power et al., (2014) | >6 score | 1 | ·· | 75 (22 to 99) | 89 (51 to 99) | ·· | ·· |
| **Self-Percieved Health Status** | Lusardi et al., (2017) | Fair or Poor | 3 | ·· | 31 (27 to 36) | 64 (60 to 68) | 0.9 (0.7 to 1) | 1.1 (1 to 1.2) |
| **Sensory Organization Test** | Power et al., (2014) | NR | 1 | ·· | 32 (NR to NR) | 93 (NR to NR) | ·· | ·· |
| **Sickness Impact Profile** | Lusardi et al., (2017) | $\geq$8 score | 1 | ·· | 13 (5 to 26) | 77 (70 to 83) | 0.6 (0.3 to 1.3) | 1.1 (1 to 1.3) |
| **Single Leg Stance** | Power et al., (2014) | <5 seconds | 1 | ·· | 33 (NR to NR) | 58 (NR to NR) | ·· | ·· |
|  | Lusardi et al., (2017) | <7.6 seconds | 1 | ·· | 46 (42 to 50) | 65 (63 to 68) | 1.3 (1.2 to 1.5) | 0.8 (0.8 to 0.9) |
|  | Power et al., (2014) | change in arm position during <5s | 1 | ·· | 56 (NR to NR) | 71 (NR to NR) | ·· | ·· |
| **Tandem Walk** | Lusardi et al., (2017) | Unable to complete | 1 | ·· | 96 (90 to 99) | 26 (14 to 42) | 1.3 (1.1 to 1.6) | 0.2 (0.1 to 0.5) |
| **Trail-Walking Test** | Power et al., (2014) | NR | 1 | ·· | 66 (NR to NR) | 84 (NR to NR) | ·· | ·· |
| **TUG (3m)** | Power et al., (2014) | >9 seconds | 1 | ·· | 50 (9 to 91) | 56 (40 to 96) | ·· | ·· |
|  | Meekes et al., (2021) | 12 seconds | 1 | ·· | 71 (56 to 83) | 28 (21 to 37) | ·· | ·· |
|  | Power et al., (2014) | $\leq$12.3 seconds | 1 | ·· | 83 (NR to NR) | 97 (NR to NR) | ·· | ·· |
|  | Meekes et al., (2021) | 12.34 seconds | 1 | ·· | 83 (NR to NR) | 97 (NR to NR) | ·· | ·· |
|  | Power et al., (2014) | 12.47 seconds | 1 | ·· | 74 (NR to NR) | 66 (NR to NR) | ·· | ·· |
|  | Meekes et al., (2021) | 12.47 seconds | 1 | ·· | 74 (NR to NR) | 66 (NR to NR) | ·· | ·· |
|  | Meekes et al., (2021) | 12.6 seconds | 1 | ·· | 31 (NR to NR) | 90 (NR to NR) | ·· | ·· |
|  | Power et al., (2014) | $\geq$12-13 seconds | 1 | ·· | 67 (NR to NR) | 50 (NR to NR) | ·· | ·· |
|  | Power et al., (2014) | $\geq$12-13 seconds | 1 | ·· | 78 (NR to NR) | 37 (NR to NR) | ·· | ·· |
|  | Meekes et al., (2021) | $\geq$12-13 seconds | 1 | ·· | 67 (NR to NR) | 50 (NR to NR) | ·· | ·· |
|  | Meekes et al., (2021) | $\geq$12-13 seconds | 1 | ·· | 78 (nr to nr) | 37 (NR to NR) | ·· | ·· |
|  | Barry et al., (2014) | $\geq$ 13.5 seconds | 10 | ·· | 32 (14 to 57) | 73 (51 to 88) | 1.2 (0.82 to 1.75) | 0.93 (0.78 to 1.1) |
|  | Meekes et al., (2021) | NR | 1 | ·· | 50 (9 to 91) | 56 (40 to 96) | ·· | ·· |
|  | Meekes et al., (2021) | NR | 1 | ·· | 10 (NR to NR) | 95 (NR to NR) | ·· | ·· |
| *Notes*.  NR – not reported  TUG – Timed Up and Go | | | | | | | | |

### ***Appendix 2 – Recurrent Falls Results***

#### Table S2.1. Summary of Meta-Analysis Relationships Predicting Recurrent Falls

| **Factor** | **# of Reviews** | **Total # of relationships** | **# of Relationships predicting increased fall risk** | **# of Relationships predicting null fall risk** | **% of Relationships that were Significant** |
| --- | --- | --- | --- | --- | --- |
| Age | 1 | 1 | 1 | 0 | 100% |
| Balance | 3 | 3 | 1 | 2 | 33% |
| Body composition | 2 | 12 | 0 | 12 | 0% |
| Chronic conditions | 3 | 11 | 9 | 2 | 82% |
| Cognition | 1 | 1 | 1 | 0 | 100% |
| Disability | 1 | 2 | 2 | 0 | 100% |
| Dual task ability | 1 | 1 | 1 | 0 | 100% |
| Education | 1 | 1 | 0 | 1 | 0% |
| Environment | 1 | 1 | 0 | 1 | 100% |
| Fall-specific factors | 2 | 3 | 3 | 0 | 100% |
| Frailty | 2 | 9 | 9 | 0 | 100% |
| Gait | 1 | 1 | 1 | 0 | 100% |
| Gender | 1 | 1 | 1 | 0 | 100% |
| Health status | 1 | 1 | 1 | 0 | 100% |
| Medications | 5 | 13 | 7 | 6 | 54% |
| Physical function | 1 | 1 | 1 | 0 | 100% |
| Nutrition | **1** | 1 | 0 | 1 | 0% |
| Pain | 2 | 2 | 2 | 0 | 100% |
| Physical activity | 2 | 2 | 1 | 1 | 50% |
| Psychosocial | 2 | 3 | 3 | 0 | 100% |
| Sensorimotor | 1 | 1 | 1 | 0 | 100% |
| Strength | 2 | 3 | 2 | 1 | 67% |
| Vision | 1 | 1 | 1 | 0 | 100% |
| *Notes.*  This table summarizes the relationships from the meta-analysis results via counts and proportions  This table does not include the relationships examining predictive validity  Data does not take into consideration magnitude, quality, or sample sizes | | | | | |

#### Table S2.2. Description of Meta-Analysis Relationships Predicting Recurrent Falls

| **Review author** | **Population** | **Measurement of Factor** | **Construct Assessed** | **# of Primary Studies** | **Pooled Sample Size** | **Model / Statistics** | **Effect Sizes (95% CI)** | **I2 (%)** |
| --- | --- | --- | --- | --- | --- | --- | --- | --- |
| **Age** |  |  |  |  |  |  |  |  |
| Deandrea et al., (2010) | General older adult | NS | Older age | 15 | 14495 | RANDOM,OR | 1.12 (1.07 to 1.18)* | 53%* |
| **Balance** |  |  |  |  |  |  |  |  |
| Jehu et al., (2020) | General older adult | ClinTest | General | 13 | NR | RANDOM,RR | 1.33 (1.11 to 1.6)* | NR |
| Muir et al., (2010) | General older adult | ClinTest | Impairment | 4 | 3071 | RANDOM,OR | 1.31 (0.98 to 1.77) | NR |
| Xu et al., (2018) | Stroke | NS | Impairment | 3 | 264 | RANDOM,OR | 2.4 (0.68 to 8.49) | 81% |
| **Body composition (BMI; kg/m^2^)** |  |  |  |  |  |  |  |  |
| Deandrea et al., (2010) | General older adult | NS | Low vs Intermediate/High | 6 | 6985 | RANDOM,OR | 1.03 (0.86 to 1.23) | 0% |
| Trevisan et al., (2019) | General older adult | NS | 17 vs 23.5 | 23 | 120185 | RANDOM,RR | 1.07 (0.98 to 1.16) | NR |
| Trevisan et al., (2019) | General older adult | NS | 18.5 vs 23.5 | 23 | 120185 | RANDOM,RR | 1.05 (0.99 to 1.12) | NR |
| Trevisan et al., (2019) | General older adult | NS | 20 vs 23.5 | 23 | 120185 | RANDOM,RR | 1.03 (0.99 to 1.08) | NR |
| Trevisan et al., (2019) | General older adult | NS | 25 vs 23.5 | 23 | 120185 | RANDOM,RR | 0.99 (0.97 to 1.01) | NR |
| Trevisan et al., (2019) | General older adult | NS | 27.5 vs 23.5 | 23 | 120185 | RANDOM,RR | 0.98 (0.94 to 1.02) | NR |
| Trevisan et al., (2019) | General older adult | NS | 30 vs 23.5 | 23 | 120185 | RANDOM,RR | 0.97 (0.91 to 1.04) | NR |
| Trevisan et al., (2019) | General older adult | NS | 35 vs 23.5 | 23 | 120185 | RANDOM,RR | 0.97 (0.84 to 1.12) | NR |
| Trevisan et al., (2019) | General older adult | NS | 37.5 vs 23.5 | 23 | 120185 | RANDOM,RR | 0.97 (0.81 to 1.16) | NR |
| Trevisan et al., (2019) | General older adult | NS | <20 | 15 | NR | RANDOM,RR | 1.06 (0.98 to 1.13) | NR |
| Trevisan et al., (2019) | General older adult | NS | 25-29.9 | 15 | NR | RANDOM,RR | 0.99 (0.86 to 1.13) | NR |
| Trevisan et al., (2019) | General older adult | NS | $\geq$30 | 15 | NR | RANDOM,RR | 0.94 (0.83 to 1.08) | NR |
| **Chronic conditions** |  |  |  |  |  |  |  |  |
| Deandrea et al., (2010) | General older adult | NS | Number of conditions | 8 | 9875 | RANDOM,OR | 1.40 (1.22 to 1.6)* | 54.4%* |
| Jehu et al., (2021) | General older adult | PROM | Comorbidity | 7 | NR | RANDOM,RR | 1.26 (NR to NR) | NR |
| Deandrea et al., (2010) | General older adult | NS | Diabetes | 7 | 18511 | RANDOM,OR | 1.28 (1.09 to 1.5)* | 34% |
| Deandrea et al., (2010) | General older adult | NS | Dizziness/Vertigo | 8 | Unclear | RANDOM,OR | 2.28 (1.9 to 2.75)* | 28% |
| Deandrea et al., (2010) | General older adult | NS | Hearing impairment | 8 | Unclear | RANDOM,OR | 1.53 (1.33 to 1.76)* | 93% |
| Deandrea et al., (2010) | General older adult | NS | Stroke | 7 | 9469 | RANDOM,OR | 1.79 (1.51 to 2.13)* | NR |
| Deandrea et al., (2010) | General older adult | NS | Hypotension | 6 | Unclear | RANDOM,OR | 1.31 (0.95 to 1.81) | 55.7%* |
| Pesonen et al., (2020) | General older adult | NS | Nocturia | 3 | NR | RANDOM,RR | 1.38 (1.11 to 1.71)* | 55% |
| Deandrea et al., (2010) | General older adult | NS | Parkinson's disease | 5 | 11308 | RANDOM,OR | 2.84 (1.77 to 4.58)* | NR |
| Deandrea et al., (2010) | General older adult | NS | Rheumatic disease | 10 | Unclear | RANDOM,OR | 1.57 (1.42 to 1.73)* | NR |
| Deandrea et al., (2010) | General older adult | NS | Urinary incontinence | 11 | Unclear | RANDOM,OR | 1.67 (1.45 to 1.92)* | 38% |
| **Disability** |  |  |  |  |  |  |  |  |
| Deandrea et al., (2010) | General older adult | NS | Instrumental disability (yes vs no) | 4 | 5000 | RANDOM,OR | 2.04 (1.41 to 2.95)* | 79.5%* |
| Deandrea et al., (2010) | General older adult | NS | Physical disability (yes vs no) | 8 | 5284 | RANDOM,OR | 2.42 (1.8 to 3.26)* | 77.4%* |
| **Cognition** |  |  |  |  |  |  |  |  |
| Deandrea et al., (2010) | General older adult | NS | Cognitive impairment | 12 | Unclear | RANDOM,OR | 1.56 (1.26 to 1.94)* | 53%* |
| **Dual task ability** |  |  |  |  |  |  |  |  |
| Mullin et al., (2021) | General older adult | NS | Motoric Cognitive Risk | 1 | 5958 | RANDOM,HR | 1.46 (1.04 to 2.05)* | NA |
| **Education** |  |  |  |  |  |  |  |  |
| Deandrea et al., (2010) | General older adult | NS | Low vs Intermediate/High) | 8 | 9655 | RANDOM,OR | 0.81 (0.62 to 1.05) | 70.3%* |
| **Fall-specific factors** |  |  |  |  |  |  |  |  |
| Deandrea et al., (2010) | General older adult | NS | Fear of falling | 7 | 6208 | RANDOM,OR | 2.51 (1.78 to 3.54)* | 70.9%* |
| Deandrea et al., (2010) | General older adult | NS | History of fall | 12 | Unclear | RANDOM,OR | 3.46 (2.85 to 4.22)* | 46%* |
| Xu et al., (2018) | Stroke | NS | History of fall | 4 | 786 | RANDOM,OR | 4.19 (2.5 to 7.01)* | 13% |
| **Frailty** |  |  |  |  |  |  |  |  |
| Cheng et al., (2017) | General older adult | PROM | Frail vs prefrail | 2 | NR | M,OR | 1.92 (1.71 to 2.15)* | NR |
| Chu et al., (2013) | General older adult | NS | Frail vs prefrail | 1 | NR | RANDOM,HR | 1.56 (1.15 to 2.13)* | 0% |
| Cheng et al., (2017) | General older adult | PROM | Frail vs prefrail | 1 | NR | M,HR | 1.56 (1.15 to 2.13)* | NR |
| Cheng et al., (2017) | General older adult | PROM | Frail vs robust | 2 | NR | M,OR | 2.77 (2.06 to 3.72)* | NR |
| Chu et al., (2013) | General older adult | NS | Frail vs robust | 1 | NR | RANDOM,HR | 2.01 (1.37 to 2.95)* | 0% |
| Cheng et al., (2017) | General older adult | PROM | Frail vs robust | 1 | NR | M,HR | 2.01 (1.37 to 2.94)* | NR |
| Cheng et al., (2017) | General older adult | PROM | Prefrail vs robust | 2 | NR | M,OR | 1.42 (1.17 to 1.73)* | NR |
| Chu et al., (2013) | General older adult | NS | Prefrail vs robust | 1 | NR | RANDOM,HR | 1.3 (1.03 to 1.65)* | 0% |
| Cheng et al., (2017) | General older adult | PROM | Prefrail vs robust | 1 | NR | M,HR | 1.3 (1.03 to 1.65)* | NR |
| **Gait** |  |  |  |  |  |  |  |  |
| Deandrea et al., (2010) | General older adult | NS | Impairment | 6 | 3646 | RANDOM,OR | 2.16 (1.47 to 3.19)* | 86%* |
| **Gender** |  |  |  |  |  |  |  |  |
| Deandrea et al., (2010) | General older adult | NS | Female gender | 18 | Unclear | RANDOM,OR | 1.34 (1.12 to 1.60)* | 66.1%* |
| **Health status** |  |  |  |  |  |  |  |  |
| Deandrea et al., (2010) | General older adult | SR | Self-perceived health status | 8 | Unclear | RANDOM,OR | 1.82 (1.26 to 2.61)* | 87.2%* |
| **Medications** |  |  |  |  |  |  |  |  |
| Jehu et al., (2021) | General older adult | NS | Number of medications | 9 | NR | RANDOM,RR | 1.51 (1.07 to 2.11)* | NR |
| Deandrea et al., (2010) | General older adult | NS | Number of medications | 12 | NR | RANDOM,OR | 1.05 (1.04 to 1.07)* | 0% |
| Ming et al., (2018) | General older adult | NS | Antidepressants | 2 | 11149 | RANDOM,OR | 2.37 (1.95 to 2.88)* | 0% |
| Deandrea et al., (2010) | General older adult | NS | Antiepileptics | 5 | 14660 | RANDOM,OR | 2.68 (1.83 to 3.92)* | 0% |
| Ang et al., (2018) | General older adult | NS | Antihypertensives-angiotensin receptor blockers | 1 | 2948 | RANDOM,OR | 1.17 (0.8 to 1.71) | NA |
| Ang et al., (2018) | General older adult | NS | Antihypertensives-calcium channel blockers | 1 | 2948 | RANDOM,OR | 1.06 (0.84 to 1.34) | NA |
| Ang et al., (2018) | General older adult | NS | Antihypertensives-beta blockers | 1 | 2948 | RANDOM,OR | 1.06 (0.82 to 1.36) | NA |
| Ang et al., (2018) | General older adult | NS | Antihypertensives-alpha blockers | 1 | 2948 | RANDOM,OR | 1.25 (0.94 to 1.66) | NA |
| Ang et al., (2018) | General older adult | NS | Antihypertensives- angiotensin converting enzyme inhibitors | 1 | 2948 | RANDOM,OR | 1.10 (0.85 to 1.42) | NA |
| Deandrea et al., (2010) | General older adult | NS | Antihypertensives | 7 | 17754 | RANDOM,OR | 1.23 (1.04 to 1.45)* | 51% |
| Ang et al., (2018) | General older adult | NS | Diuretic | 5 | 19573 | RANDOM,OR | 1.12 (0.91 to 138) | 53% |
| Deandrea et al., (2010) | General older adult | NS | Sedatives | 10 | 21861 | RANDOM,OR | 1.53 (1.34 to 1.75)* | 0% |
| Xu et al., (2018) | Stroke | NS | Sedatives & psychotropics | 3 | 671 | RANDOM,OR | 2.23 (1.18 to 4.23)* | 94% |
| **Physical function** |  |  |  |  |  |  |  |  |
| Deandrea et al., (2010) | General older adult | SR | Gait aid use | 6 | Unclear | RANDOM,OR | 3.09 (2.1 to 4.53)* | 68%* |
| **Nutrition** |  |  |  |  |  |  |  |  |
| Trevisan et al., (2019) | General older adult | PROM | Nourishment | 2 | 970 | RANDOM,RR | 1.33 (0.85 to 2.09) | 36% |
| **Pain** |  |  |  |  |  |  |  |  |
| Deandrea et al., (2010) | General older adult | NS | Present | 6 | Unclear | RANDOM,OR | 1.6 (1.44 to 1.78)* | 0% |
| Stubbs et al., (2014b) | General older adult | SR | Present | 3 | 2646 | RANDOM,OR | 1.79 (1.44 to 2.21)* | 0% |
| **Physical activity** |  |  |  | 2 |  |  |  |  |
| Deandrea et al., (2010) | General older adult | SR | Limitation | 9 | Unclear | RANDOM,OR | 1.13 (0.94 to 1.34) | 50%* |
| Soares et al., (2019) | General older adult | PROM | Level | 2 | 2420 | RANDOM,RR | 1.39 (1.17 to 1.65)* | 0% |
| **Psychosocial** |  |  |  |  |  |  |  |  |
| Deandrea et al., (2010) | General older adult | NS | Depression | 15 | Unclear | RANDOM,OR | 1.86 (1.45 to 2.38)* | 86% |
| Deandrea et al., (2010) | General older adult | NS | Living situation | 9 | Unclear | RANDOM,OR | 1.25 (1.1 to 1.43)* | 60% |
| Jehu et al., (2021) | General older adult | NS | Psychological wellbeing | 9 | NR | RANDOM,RR | 1.39 (1.08 to 1.79)* | NR |
| **Sensorimotor** |  |  |  |  |  |  |  |  |
| Jehu et al., (2021) | General older adult | NS | Sensorimotor ability | 11 | NR | RANDOM,RR | 1.51 (1.29 to 1.78)* | NR |
| **Strength** |  |  |  |  |  |  |  |  |
| Moreland et al., (2004) | General older adult | ClinTest | Weakness-lower extremity | 3 | NR | RANDOM,OR | 2.7 (1.96 to 3.72)* | NR |
| Xu et al., (2018) | Stroke | NS | Impairment-lower extremity | 3 | 264 | RANDOM,OR | 0.88 (0.3 to 2.53) | 72% |
| Moreland et al., (2004) | General older adult | ClinTest | Weakness-upper extremity | 3 | 2713 | RANDOM,OR | 1.41 (1.25 to 1.59)* | NR |
| **Vision** |  |  |  |  |  |  |  |  |
| Deandrea et al., (2010) | General older adult | NS | Impairment | 13 | Unclear | RANDOM,OR | 1.6 (1.28 to 2.00)* | 74%* |
| *Notes.*  *Indicates significance  NR – not reported  Sample characteristics: CI – cognitive impairment, OA - osteoarthritis  Measurement of factor: PROM – Patient Reported Outcome Measure;  Models: R – Random Model; F – Fixed Model; OR – Odds Ratio; RR –Relative Risk; SMD – Standard Mean Difference; MD – Mean Difference | | | | | | | | |

#### Table S2.3. Summary of Single Relationships Predicting Recurrent Falls

| **Risk Factor** | **Total # of relationships** | **# of Relationships with Increased Risk** | **# of Relationships with Null Risk** | **# of Relationships with Reduced Risk** | **% of Significant Relationships** |
| --- | --- | --- | --- | --- | --- |
| Balance | 26 | 20 | 6 | 0 | 77% |
| Body composition | 2* | 1 | 0 | 0 | 50% |
| Cognition | 11 | 9 | 2 | 0 | 82% |
| Chronic conditions | 6 | 6 | 0 | 0 | 100% |
| Dual task ability | 3 | 2 | 1 | 0 | 67% |
| Fall-specific factors | 3 | 3 | 0 | 0 | 100% |
| Gait | 31 | 22 | 9 | 0 | 71% |
| Gender | 1 | 1 | 0 | 0 | 100% |
| Medications | 1 | 0 | 1 | 0 | 0% |
| Mobility | 11 | 10 | 1 | 0 | 91% |
| Pain | 2 | 2 | 0 | 0 | 100% |
| Physical activity | 1 | 1 | 0 | 0 | 100% |
| Psychosocial | 2 | 2 | 0 | 0 | 100% |
| Sensorimotor | 8 | 7 | 1 | 0 | 88% |
| Sensory | 2 | 2 | 0 | 0 | 100% |
| Strength | 3 | 2 | 1 | 0 | 67% |
| Vision | 30 | 17 | 13 | 0 | 57% |
| *Notes.*  This table summarizes the relationships from the narratively reported results via counts and proportions  This table does not include the relationships examining predictive validity  Data does not take into consideration magnitude, quality, or sample sizes  *1 study reported BMI as being evaluated but did not report results | | | | | |

#### Table S2.4. Description of Single Relationships Predicting Recurrent Falls

| **Review author** | **Population** | **Measurement of Factor** | **Construct Assessed** | **# of Relationships Predicting Increased Risk** | **# of Relationships Predicting Null Risk** | **# of Relationships Predicting Reduced Risk** | **Total # of Relationships** | **% of Relationships that were Significant** |
| --- | --- | --- | --- | --- | --- | --- | --- | --- |
| **Balance** |  |  |  |  |  |  |  |  |
| Lima et al., (2018); Power et al., (2014) | General older adult | ClinTest | Berg Balance Scale-reduced | 1 | 3 | 0 | 4 | 25% |
| Piirtola et al. (2006); Power et al., (2014); | General older adult | ClinTest | Tandem Stance Test | 3 | 0 | 0 | 3 | 100% |
| Marin et al., (2022); Power et al., (2014); Piirtola et al., (2006) | General older adult | ClinTest | POMA Balance Test-reduced | 3 | 0 | 0 | 3 | 100% |
| Power et al., (2014) | General older adult | ClinTest | Functional Reach Test-increased distance | 1 | 0 | 0 | 1 | 100% |
| Power et al., (2014) | General older adult | ClinTest | Functional Reach Test-reduced distance | 1 | 0 | 0 | 1 | 100% |
| Power et al., (2014) | General older adult | ClinTest | Romberg Test-reduced | 1 | 0 | 0 | 1 | 100% |
| Power et al., (2014) | General older adult | ClinTest | Sensory Organization Test-reduced | 1 | 0 | 0 | 1 | 100% |
| Fernando et al., (2017) | Dementia | ClinTest | Coordinated Stability Test | 1 | 0 | 0 | 1 | 100% |
| Fernando et al., (2017); Marin et al., (2022); Piirtola et al. (2006); Power et al., (2014); | General older adult | Post. | Sway amplitude-increasing | 6 | 1 | 0 | 7 | 86% |
| Piirtola et al. (2006) | General older adult | Post. | Sway area-increased | 1 | 0 | 0 | 1 | 100% |
| Power et al., (2014) | General older adult | Post. | Sway position-increased | 1 | 0 | 0 | 1 | 100% |
| Piirtola et al. (2006) | General older adult | Post. | Sway velocity-increased | 0 | 1 | 0 | 1 | 0% |
| Fernando et al., (2017) | Dementia | NS | Proprioception-reduced | 0 | 1 | 0 | 1 | 0% |
| **Body composition** |  |  |  |  |  |  |  |  |
| Gravesande et al., (2017) | Diabetes Mellitus | NS | Overweight | 1 | 0 | 0 | 1 | 100% |
| Gravesande et al., (2017) | Diabetes Mellitus | NS | BMI | 0 | 0 | 0 | 1^*^ | 0% |
| **Chronic conditions** |  |  |  |  |  |  |  |  |
| Sharma et al., (2023) | General older adult | Imaging | Cerebral small vessel disease | 3 | 0 | 0 | 3 | 100% |
| Gravesande et al., (2017) | Diabetes Mellitus | NS | Number of conditions | 1 | 0 | 0 | 1 | 100% |
| Ganz et al., (2007) | General older adult | NS | Dizziness | 1 | 0 | 0 | 1 | 100% |
| Ganz et al., (2007) | General older adult | ClinTest | Orthostatic hypotension | 1 | 0 | 0 | 1 | 100% |
| **Cognition** |  |  |  |  |  |  |  |  |
| Kearney et al., (2013) | General older adult | NS | Executive function-reduced | 6 | 0 | 0 | 6 | 100% |
| Fernando et al., (2017); Muir et al., (2010) | Dementia | NS | Dementia severity | 1 | 2 | 0 | 3 | 33% |
| Kearney et al., (2013) | General older adult | PROM | Memory-reduced | 1 | 0 | 0 | 1 | 100% |
| Kearney et al., (2013) | General older adult | PROM | Impairment | 1 | 0 | 0 | 1 | 100% |
| **Dual task ability** |  |  |  |  |  |  |  |  |
| Muir-Hunter et al., (2016) | General older adult | ClinTest | Dual task | 2 | 1 | 0 | 3 | 67% |
| **Fall-specific factors** |  |  |  |  |  |  |  |  |
| Ganz et al., (2007) | General older adult | SR | Fall history | 2 | 0 | 0 | 2 | 100% |
| Ganz et al., (2007) | General older adult | SR | Fear of falling | 1 | 0 | 0 | 1 | 100% |
| **Gait** |  |  |  |  |  |  |  |  |
| Fernando et al., (2017); Ganz et al., (2007); Marin et al., (2022); Modaressi et al., (2019); Piirtola et al., (2006); Power et al., (2014); | Dementia | ClinTest, Post. | Gait velocity-reduced | 9 | 2 | 0 | 11 | 82% |
| Modaressi et al., (2019); Power et al., (2014) | Dementia | ClinTest, Post. | Cadence speed-reduced | 4 | 1 | 0 | 5 | 80% |
| Fernando et al., (2017); Modaressi et al., (2019); Power et al., (2014); | Dementia | ClinTest, Post., NS | Double support time-increased | 3 | 1 | 0 | 4 | 75% |
| Fernando et al., (2017); Modaressi et al., (2019) | General older adult | Post. | Step length variability-increased | 1 | 1 | 0 | 2 | 50% |
| Modaressi et al., (2019); Power et al., (2014) | Dementia | ClinTest, Post. | Stride length variability-increased | 1 | 1 | 0 | 2 | 50% |
| Fernando et al., (2017); Modaressi et al., (2019) | General older adult | Post. | Stride length-reduced | 0 | 2 | 0 | 2 | 0% |
| Power et al., (2014) | General older adult | ClinTest | Trendelenburg pattern | 1 | 0 | 0 | 1 | 100% |
| Power et al., (2014) | General older adult | ClinTest | Double support time variability-increased | 1 | 0 | 0 | 1 | 100% |
| Power et al., (2014) | General older adult | NS | Step time variability-increased | 1 | 0 | 0 | 1 | 100% |
| Power et al., (2014) | General older adult | ClinTest | Gait analyses stability-reduced | 1 | 0 | 0 | 1 | 100% |
| Power et al., (2014) | General older adult | ClinTest | Dynamic Gait Index-reduced | 0 | 1 | 0 | 1 | 0% |
| **Gender** |  |  |  |  |  |  |  |  |
| Ganz et al., (2007) | General older adult | SR | Female gender | 1 | 0 | 0 | 1 | 100% |
| **Medications** |  |  |  |  |  |  |  |  |
| Seppala et al., (2018) | General older adult | NS | Antiepileptics | 0 | 1 | 0 | 1 | 0% |
| **Mobility** |  |  |  |  |  |  |  |  |
| Piirtola et al. (2006); Power et al., (2014); | General older adult | ClinTest | Sit to Stand Test | 4 | 0 | 0 | 4 | 100% |
| Marin et al., (2022); Power et al., (2014) | General older adult | ClinTest | TUG Test | 4 | 0 | 0 | 4 | 100% |
| Power et al., (2014) | General older adult | ClinTest | Bending Down | 1 | 0 | 0 | 1 | 100% |
| Modaressi et al., (2019) | General older adult | Post. | Gait aid use | 0 | 1 | 0 | 1 | 0% |
| Ganz et al., (2007) | General older adult | SR | Impairment | 1 | 0 | 0 | 1 | 100% |
| **Pain** |  |  |  |  |  |  |  |  |
| Gravesande et al., (2017) | Diabetes Mellitus | NS | Number of sites-lower extremities | 2 | 0 | 0 | 2 | 100% |
| **Physical activity** |  |  |  |  |  |  |  |  |
| Piirtola et al. (2006) | General older adult | NS | Volume of physical activity-increase | 1 | 0 | 0 | 1 | 100% |
| **Psychosocial** |  |  |  |  |  |  |  |  |
| Launay et al., (2013) | General older adult | ClinTest | Depressive symptoms | 2 | 0 | 0 | 2 | 100% |
| **Sensorimotor** |  |  |  |  |  |  |  |  |
| Fernando et al., (2017); Piirtola et al. (2006) | Dementia | ClinTest | Reaction time-reduced | 7 | 1 | 0 | 8 | 88% |
| **Strength** |  |  |  |  |  |  |  |  |
| Fernando et al., (2017); Piirtola et al. (2006) | Dementia | ClinTest | Lower extremity strength-increase | 1 | 1 | 0 | 2 | 50% |
| Gravesande et al., (2017) | Diabetes Mellitus | ClinTest | Lower extremity strength-decreased | 1 | 0 | 0 | 1 | 100% |
| **Vision** |  |  |  |  |  |  |  |  |
| Salonen et al., (2012) | General older adult | NS | Acuity | 6 | 3 | 0 | 9 | 67% |
| Salonen et al., (2012) | General older adult | NS | Cataract | 0 | 1 | 0 | 1 | 0% |
| Fernando et al., (2017); Salonen et al., (2012) | Dementia | NS | Contrast | 4 | 5 | 0 | 9 | 44% |
| Salonen et al., (2012) | General older adult | NS | Depth | 2 | 0 | 0 | 2 | 100% |
| Salonen et al., (2012) | General older adult | NS | Discrepant | 1 | 0 | 0 | 1 | 100% |
| Salonen et al., (2012) | General older adult | NS | Field | 3 | 2 | 0 | 5 | 60% |
| Salonen et al., (2012) | General older adult | NS | Glaucoma | 0 | 1 | 0 | 1 | 0% |
| Salonen et al., (2012) | General older adult | SR | Ophthalmic disease | 1 | 0 | 0 | 1 | 100% |
| Salonen et al., (2012) | General older adult | NS | Retinal disease | 0 | 1 | 0 | 1 | 0% |
| *Notes.*  ^*^study did not report results  Measurement of factor: PROM – Patient Reported Outcome Measure; SR – Self Report; NS – Not Specified; ClinTest – Clinical Test, Post. – Posturography  Constructs: TUG – Timed up and Go | | | | | | | | |

#### Table S2.5. Predictive Validity of Tests for Recurrent Falls

| **Tool** | **Review Author** | **Cut point** | **# of primary studies** | **AUC (95%CI)** | **Sensitivity % (95% CI)** | **Specificity % (95% CI)** | **Positive Likelihood Ratio (95% CI)** | **Negative Likelihood Ratio (95% CI)** |
| --- | --- | --- | --- | --- | --- | --- | --- | --- |
| **Ability to pick up 5lbs** | Lusardi et al., (2017) | Unable to complete | 1 | ·· | 11 (5 to 20) | 93 (89 to 96) | 1.6 (0.8 to 2.4) | 1 (0.9 to 1) |
| **Berg Balance Scale** | Lima et al., (2018) | $\leq$45 points | 1 | ·· | 42 (NR to NR) | 87 (NR to NR) | ·· | ·· |
|  | Lima et al., (2018) | $\leq$53 points | 1 | ·· | 69 (NR to NR) | 57 (NR to NR) | ·· | ·· |
|  | Meekes et al., (2021) | $\leq$53 points | 1 | 0.68 (NR to NR) | 69 (50 to 83) | 57 (47 to 66) | ·· | ·· |
|  | Meekes et al., (2021) | $\leq$45 points | 1 | ·· | 42 (26 to 61) | 87 (79 to 92) | ·· | ·· |
| **Counting Steps** | Lusardi et al., (2017) | 4+ | 1 | ·· | 78 (67 to 86) | 28 (23 to 34) | 1.1 (0.9 to 1.2) | 0.08 (0.5 to 1.3) |
| **Fall history** | Meekes et al., (2021) | $\geq$2 falls in past year | 1 | ·· | 46 (NR to NR) | 80 (NR to NR) | ·· | ·· |
|  | Meekes et al., (2021) | Multiple falls | 1 | 0.64 (NR to NR) | NR (NR to NR) | NR (NR to NR) | ·· | ·· |
| **Functional Gait Assessment** | Õmana et al., (2021) | $\leq$17 score | 1 | 0.74 (0.67 to 0.79) | 56 (45 to 67) | 82 (75 to 88) | 0.53 (0.51 to 0.57) | 3.16 (2.82 to 3.53) |
| **Functional Reach Test** | Õmana et al., (2021) | NR | 1 | 0.62 (0.55 to 0.68) | NR (NR to NR) | NR (NR to NR) | ·· | ·· |
| **Gait Speed (4m)** | Meekes et al., (2021) | Recurrent falls | 1 | 0.54 (0.45 to 0.64) | NR (NR to NR) | NR (NR to NR) | ·· | ·· |
|  | Meekes et al., (2021) | Recurrent falls | 1 | 0.68 (0.59 to 0.77) | NR (NR to NR) | NR (NR to NR) | ·· | ·· |
| **POMA-Balance** | Meekes et al., (2021) | Multiple fallers | 1 | 0.66 (NR to NR) | 89 (NR to NR) | 47 (NR to NR) | ·· | ·· |
|  | Õmana et al., (2021) | NR | 1 | 0.66 (NR to NR) | 47 (NR to NR) | 89 (NR to NR) | ·· | ·· |
| **POMA-Total** | Õmana et al., (2021) | NR | 1 | 0.76 (NR to NR) | 67 (NR to NR) | 83 (NR to NR) | ·· | ·· |
| **Single Leg Stance** | Lusardi et al., (2017) | Upper extremity movement | 1 | ·· | 51 (46 to 55) | 60 (57 to 63) | 1.3 (1.1 to 1.4) | 0.8 (0.8 to 0.9) |
|  | Õmana et al., (2021) | <5 seconds | 1 | ·· | 33 (NR to NR) | 71.2 (NR to NR) |  |  |
| **Sit to Stand Test** | Lusardi et al., (2017) | $\geq$1 | 1 | ·· | 49 (34 to 64) | 52 (45 to 60) | 1 (7 to 1.4) | 1 (0.7 to 1.3) |
| **Step Test** | Lusardi et al., (2017) | $\geq$10 | 1 | ·· | 69 (57 to 79) | 64 (58 to 70) | 1.9 (1.5 to 2.4) | 0.5 (0.3 to 0.7) |
|  | Lusardi et al., (2017) | $\geq$5 | 1 | ·· | 54 (42 to 65) | 58 (52 to 64) | 1.3 (1 to 1.6) | 0.8 (0.6 to 1) |
|  | Lusardi et al., (2017) | $\geq$5 | 1 | ·· | 63 (51 to 73) | 55 (49 to 61) | 1.4 (1.1 to 1.7) | 0.7 (0.5 to 0.8) |
| **Sway** | Piirtola et al., (2006) | NR | 1 | ·· | 45 (NR to NR) | 83 (NR to NR) | ·· | ·· |
| **Sway** | Piirtola et al., (2006) | NR | 1 | ·· | 74 (NR to NR) | 61 (NR to NR) | ·· | ·· |
| **Tug (3m)** | Meekes et al., (2021) | Recurrent falls | 1 | 0.69 (0.60 to 0.77) | NR (NR to NR) | NR (NR to NR) | ·· | ·· |
|  | Meekes et al., (2021) | $\geq$10.15 seconds | 1 | 0.73 (0.65 to 0.82) | 67 (NR to NR) | 56 (NR to NR) | ·· | ·· |
| *Notes*.  NR – not reported  TUG – Timed Up and Go | | | | | | | | |

### ***Appendix 3 – Injurious Falls Results***

#### Table S3.1. Summary of Meta-Analysis Relationships Predicting Injurious Falls

| **Factor** | **# of Reviews** | **Total # of relationships** | **# of Relationships Predicting Increased Risk** | **# of Relationships Predicting Null Risk** | **% Relationships that were Significant** |
| --- | --- | --- | --- | --- | --- |
| Balance | 1 | 2 | 2 | 0 | 100% |
| Cognition | 1 | 2 | 2 | 0 | 100% |
| Dual task ability | 1 | 1 | 1 | 0 | 100% |
| Strength | 1 | 2 | 2 | 0 | 100% |
| Medications | 1 | 3 | 1 | 2 | 33% |
| *Notes.*  This table summarizes the relationships from the meta-analysis results via counts and proportions  This table does not include the relationships examining predictive validity  Data does not take into consideration magnitude, quality, or sample sizes | | | | | |

#### Table S3.2. Description of Meta-Analysis Relationships Predicting Injurious Falls

| **Review author** | **Population** | **Measurement of Factor** | **Construct Assessed** | **# of Primary Studies** | **Pooled Sample Size** | **Model / Statistics** | **Effect Sizes (95% CI)** | **I2 (%)** |
| --- | --- | --- | --- | --- | --- | --- | --- | --- |
| **Balance** |  |  |  |  |  |  |  |  |
| Muir et al., (2010) | General older adult | ClinTest | Impairment | 2 | 17279 | RANDOM,RR | 1.25 (1.02 to 1.54)* | NR |
| Muir et al., (2010) | General older adult | ClinTest | Impairment | 3 | 2948 | RANDOM,RR | 1.42 (1 to 2.02)* | NR |
| **Cognition** |  |  |  |  |  |  |  |  |
| Muir et al., (2012) | General older adult | NS | Impairment | NR | NR | RANDOM,OR | 2.33 (1.61 to 3.36)* | 6% |
| Muir et al., (2012) | General older adult | NS | Impairment | NR | NR | RANDOM,RR | 1.78 (1.34 to 2.37)* | 0% |
| **Dual task ability** |  |  |  |  |  |  |  |  |
| Mullin et al., (2021) | General older adult | NS | Motoric Cognitive Risk | 1 | 5958 | RANDOM,HR | 2.54 (1.78 to 3.63)* | NR |
| **Strength** |  |  |  |  |  |  |  |  |
| Moreland et al., (2004) | General older adult | ClinTest | Lower extremity weakness | 2 | 1428 | RANDOM,OR | 1.52 (1.05 to 2.2)* | NR |
| Moreland et al., (2004) | General older adult | ClinTest | Upper extremity weakness | 1 | 1103 | RANDOM,OR | 1.77 (1.13 to 2.77)* | NR |
| **Medications** |  |  |  |  |  |  |  |  |
| Ang et al., (2018) | General older adult | NS | Antihypertensives - alpha blockers | 1 | 598 | RANDOM,OR | 0.61 (0.24 to 1.55) | NR |
| Ang et al., (2018) | General older adult | NS | Antihypertensives- angiotensin converting enzyme inhibitors | 1 | 598 | RANDOM,OR | 0.62 (0.4 to 0.96)* | NR |
| Ang et al., (2018) | General older adult | NS | Antihypertensives - angiotensin receptor blockers | 1 | 598 | RANDOM,OR | 1.01 (0.54 to 1.88) | NR |
| *Notes.*  *Indicates significance  NR – not reported  Sample characteristics: CI – cognitive impairment, OA - osteoarthritis  Measurement of factor: PROM – Patient Reported Outcome Measure;  Models: R – Random Model; F – Fixed Model; OR – Odds Ratio; RR –Relative Risk; SMD – Standard Mean Difference; MD – Mean Difference; | | | | | | | | |

#### Table S3.3. Summary of Single Relationships Predicting Injurious Falls

| **Risk Factor** | **# of Relationships with Increased Risk** | **# of Relationships with Null Risk** | **# of Relationships with Reduced Risk** | **Total # of relationships** | **% of Significant Relationships** |
| --- | --- | --- | --- | --- | --- |
| Balance | 2 | 13 | 8 | 23 | 43% |
| Cognition | 6 | 3 | 2 | 11 | 73% |
| Disability | 1 | 0 | 0 | 1 | 100% |
| Gait | 1 | 0 | 5 | 6 | 100% |
| Medications | 2 | 0 | 3 | 5 | 100% |
| Mobility | 2 | 0 | 0 | 2 | 100% |
| Psychosocial | 7 | 0 | 0 | 7 | 100% |
| *Notes.*  This table summarizes the relationships from the narratively reported results via counts and proportions  This table does not include the relationships examining predictive validity  Data does not take into consideration magnitude, quality, or sample sizes | | | | | |

#### Table S3.4. Description of Single Relationships Predicting Injurious Falls

| **Review author** | **Population** | **Measurement of Factor** | **Construct Assessed** | **# of Relationships Predicting Increased Risk** | **# of Relationships Predicting Null Risk** | **# of Relationships Predicting Reduced Risk** | **Total # of Relationships** | **% of Relationships that were Significant** |
| --- | --- | --- | --- | --- | --- | --- | --- | --- |
| **Balance** |  |  |  |  |  |  |  |  |
| Blodgett et al., (2022); Marin et al., (2022) | General older adult | ClinTest | Single Leg Stance Test | 2 | 8 | 13 | 23 | 65% |
| **Cognition** |  |  |  |  |  |  |  |  |
| Chantanachai et al., (2021) | Dementia | ClinTest, PROM, NS | Behaviour changes | 0 | 1 | 0 | 1 | 0% |
| Kearney et al., (2013); Muir et al., (2012) | General older adult | ClinTest, NS | Executive function | 3 | 1 | 0 | 4 | 75% |
| Kearney et al., (2013) | General older adult | PROM | Impairment | 0 | 0 | 1 | 1 | 100% |
| Kearney et al., (2013); Muir et al., (2012) | General older adult | ClinTest, NS | Processing speed | 1 | 0 | 2 | 3 | 100% |
| Muir et al., (2012) | General older adult | NS | Global cognition | 2 | 0 | 0 | 2 | 100% |
| **Disability** |  |  |  |  |  |  |  |  |
| Chantanachai et al., (2021) | Dementia | PROM | ADL decline | 1 | 0 | 0 | 1 | 100% |
| **Gait** |  |  |  |  |  |  |  |  |
| Chantanachai et al., (2021) | MCI | NS | Cadence speed | 0 | 1 | 0 | 1 | 0% |
| Chantanachai et al., (2021) | MCI | NS | Double support time | 0 | 1 | 0 | 1 | 0% |
| Chantanachai et al., (2021) | MCI | NS | Spatial variability | 0 | 1 | 0 | 1 | 0% |
| Chantanachai et al., (2021) | MCI | NS | Stride length variability | 0 | 1 | 0 | 1 | 0% |
| Chantanachai et al., (2021) | MCI | NS | Swing time variability | 0 | 1 | 0 | 1 | 0% |
| Chantanachai et al., (2021) | MCI | NS | Temporal variability | 1 | 0 | 0 | 1 | 100% |
| **Medications** |  |  |  |  |  |  |  |  |
| Seppala et al., (2018) | General older adult | NS | Antiepileptics | 1 | 1 | 0 | 2 | 50% |
| Seppala et al., (2018) | General older adult | NS | Proton-pump inhibitors | 1 | 0 | 0 | 1 | 100% |
| Seppala et al., (2018) | General older adult | NS | Prostatics | 0 | 2 | 0 | 2 | 0% |
| **Physical function** |  |  |  |  |  |  |  |  |
| Marin et al., (2022) | General older adult | ClinTest | TUG Test | 1 | 0 | 0 | 1 | 100% |
| **Psychosocial** |  |  |  |  |  |  |  |  |
| Fernando et al., (2017) | General older adult | SR | Living alone | 4 | 0 | 0 | 4 | 100% |
| Petersen et al., (2020) | General older adult | PROM | Caregiver distress | 3 | 0 | 0 | 3 | 100% |
| *Notes.*  Sample characteristics: CI – cognitive impairment, MCI – mild cognitive impairment  Measurement of factor: PROM – Patient Reported Outcome Measure; SR – Self Report; NS – Not Specified; ClinTest – Clinical Test, Post. – Posturography  Construct: ADL – activities of daily living, TUG – Timed Up and Go | | | | | | | | |

### ***Appendix 4 – Reference List of Included Studies***

Ang, H. T., Lim, K. K., Kwan, Y. H., Tan, P. S., Yap, K. Z., Banu, Z., . . . Ostbye, T. (2018). A systematic review and meta-analyses of the association between anti-hypertensive classes and the risk of falls among older adults. Drugs & Aging, 35, 625-635.

Barry, E., Galvin, R., Keogh, C., Horgan, F., & Fahey, T. (2014). Is the Timed Up and Go test a useful predictor of risk of falls in community dwelling older adults: a systematic review and meta-analysis. BMC geriatrics, 14, 1-14.

Blodgett, J. M., Ventre, J. P., Mills, R., Hardy, R., & Cooper, R. (2022). A systematic review of one-legged balance performance and falls risk in community-dwelling adults. Ageing Research Reviews, 73, 101501.

Chantanachai, T., Sturnieks, D. L., Lord, S. R., Payne, N., Webster, L., & Taylor, M. E. (2021). Risk factors for falls in older people with cognitive impairment living in the community: Systematic review and meta-analysis. Ageing Research Reviews, 71, 101452.

Cheng, M. H., & Chang, S. F. (2017). Frailty as a risk factor for falls among community dwelling people: Evidence from a meta‐analysis. Journal of nursing scholarship, 49(5), 529-536.

Chu, W., Chang, S. F., & Ho, H. Y. (2021). Adverse health effects of frailty: Systematic review and meta‐analysis of middle‐aged and older adults with implications for evidence‐based practice. Worldviews on Evidence‐Based Nursing, 18(4), 282-289.

Deandrea, S., Lucenteforte, E., Bravi, F., Foschi, R., La Vecchia, C., & Negri, E. (2010). Risk factors for falls in community-dwelling older people: A systematic review and meta-analysis. Epidemiology, 21(5), 658-668. doi:10.1097/EDE.0b013e3181e89905

Dibello, V., Lobbezoo, F., Lozupone, M., Sardone, R., Ballini, A., Berardino, G., . . . Stallone, R. (2023). Oral frailty indicators to target major adverse health-related outcomes in older age: A systematic review. Geroscience, 45(2), 663-706.

Fernando, E., Fraser, M., Hendriksen, J., Kim, C. H., & Muir-Hunter, S. W. (2017). Risk factors associated with falls in older adults with dementia: a systematic review. Physiotherapy Canada, 69(2), 161-170.

Gafner, S. C., Allet, L., Hilfiker, R., & Bastiaenen, C. H. G. (2021). Reliability and diagnostic accuracy of commonly used performance tests relative to fall history in older persons: a systematic review. Clinical interventions in aging, 1591-1616.

Gambaro, E., Gramaglia, C., Azzolina, D., Campani, D., Dal Molin, A., & Zeppegno, P. (2022). The complex associations between late life depression, fear of falling and risk of falls. A systematic review and meta-analysis. Ageing Research Reviews, 73, 101532.

Ganz, D. A., Bao, Y., Shekelle, P. G., & Rubenstein, L. Z. (2007). Will my patient fall? JAMA, 297(1), 77-86.

Gillain, S., Boutaayamou, M., Beaudart, C., Demonceau, M., Bruyere, O., Reginster, J.-Y., . . . Petermans, J. (2018). Assessing gait parameters with accelerometer-based methods to identify older adults at risk of falls: A systematic review. European Geriatric Medicine, 9, 435-448.

Gravesande, J., & Richardson, J. (2017). Identifying non-pharmacological risk factors for falling in older adults with type 2 diabetes mellitus: A systematic review. Disability & Rehabilitation, 39(15), 1459-1465.

Hartog, L. C., Schrijnders, D., Landman, G., Groenier, K., Kleefstra, N., Bilo, H. J., & van Hateren, K. J. J. (2017). Is orthostatic hypotension related to falling? A meta-analysis of individual patient data of prospective observational studies. Age and Ageing, 46(4), 568-575.

Iseli, R. K., Lee, E. K., Lewis, E., Duncan, G., & Maier, A. B. (2021). Foot disease and physical function in older adults: A systematic review and meta‐analysis. Australasian Journal on Ageing, 40(1), 35-47.

Jehu, D., Davis, J., Falck, R., Bennett, K., Tai, D., Souza, M., . . . Liu-Ambrose, T. (2021). Risk factors for recurrent falls in older adults: A systematic review with meta-analysis. Maturitas, 144, 23-28.

Kearney, F. C., Harwood, R. H., Gladman, J. R., Lincoln, N., & Masud, T. (2013). The relationship between executive function and falls and gait abnormalities in older adults: A systematic review. Dementia and Geriatric Cognitive Disorders, 36(1-2), 20-35.

Kojima, G. (2015). Frailty as a predictor of future falls among community-dwelling older people: A systematic review and meta-analysis. Journal of the American Medical Directors Association, 16(12), 1027-1033.

Kvelde, T., McVeigh, C., Toson, B., Greenaway, M., Lord, S. R., Delbaere, K., & Close, J. C. (2013). Depressive symptomatology as a risk factor for falls in older people: Systematic review and meta‐analysis. Journal of the American Geriatrics Society, 61(5), 694-706.

Launay, C., De Decker, L., Annweiler, C., Kabeshova, A., Fantino, B., & Beauchet, O. (2013). Association of depressive symptoms with recurrent falls: A cross-sectional elderly population based study and a systematic review. The Journal of Nutrition, Health and Aging, 17(2), 152-157.

Leroy, V., Martinet, V., Nunkessore, O., Dentel, C., Durand, H., Mockler, D., . . . Chen, Y. (2023). The nebulous association between cognitive impairment and falls in older adults: A systematic review of the literature. International Journal of Environmental Research and Public Health, 20(3), 2628.

Letts, L., Moreland, J., Richardson, J., Coman, L., Edwards, M., Ginis, K. M., . . . Wishart, L. (2010). The physical environment as a fall risk factor in older adults: Systematic review and meta‐analysis of cross‐sectional and cohort studies. Australian Occupational Therapy Journal, 57(1), 51-64.

Lima, C., Ricci, N., Nogueira, E., & Perracini, M. R. (2018). The Berg Balance Scale as a clinical screening tool to predict fall risk in older adults: A systematic review. Physiotherapy, 104(4), 383-394.

Lusardi, M. M., Fritz, S., Middleton, A., Allison, L., Wingood, M., Phillips, E., . . . Chui, K. K. (2017). Determining risk of falls in community dwelling older adults: a systematic review and meta-analysis using posttest probability. Journal of Geriatric Physical Therapy, 40(1), 1-36.

Malik, V., Gallagher, C., Linz, D., Elliott, A. D., Emami, M., Kadhim, K., . . . Arnolda, L. (2020). Atrial fibrillation is associated with syncope and falls in older adults: A systematic review and meta-analysis. Mayo Clinic Proceedings.

Manlapaz, D. G., Sole, G., Jayakaran, P., & Chapple, C. M. (2019). Risk factors for falls in adults with knee osteoarthritis: a systematic review. PM&R, 11(7), 745-757.

Marín-Jiménez, N., Cruz-León, C., Perez-Bey, A., Conde-Caveda, J., Grao-Cruces, A., Aparicio, V. A., . . . Cuenca-García, M. (2022). Predictive validity of motor fitness and flexibility tests in adults and older adults: A systematic review. Journal of Clinical Medicine, 11(2), 328.

Meekes, W. M., Korevaar, J. C., Leemrijse, C. J., & Van de Goor, I. A. (2021). Practical and validated tool to assess falls risk in the primary care setting: A systematic review. BMJ Open, 11(9), e045431.

Menz, H. B., Auhl, M., & Spink, M. J. (2018). Foot problems as a risk factor for falls in community-dwelling older people: A systematic review and meta-analysis. Maturitas, 118, 7-14.

Ming, Y., & Zecevic, A. (2018). Medications & polypharmacy influence on recurrent fallers in community: A systematic review. Canadian Geriatrics Journal, 21(1), 14.

Modarresi, S., Divine, A., Grahn, J. A., Overend, T. J., & Hunter, S. W. (2019). Gait parameters and characteristics associated with increased risk of falls in people with dementia: A systematic review. International Psychogeriatrics, 31(9), 1287-1303.

Moreland, J. D., Richardson, J. A., Goldsmith, C. H., & Clase, C. M. (2004). Muscle weakness and falls in older adults: A systematic review and meta‐analysis. Journal of the American Geriatrics Society, 52(7), 1121-1129.

Muir, S. W., Berg, K., Chesworth, B., Klar, N., & Speechley, M. (2010). Quantifying the magnitude of risk for balance impairment on falls in community-dwelling older adults: A systematic review and meta-analysis. Journal of Clinical Epidemiology, 63(4), 389-406.

Muir, S. W., Gopaul, K., & Montero Odasso, M. M. (2012). The role of cognitive impairment in fall risk among older adults: A systematic review and meta-analysis. Age and Ageing, 41(3), 299-308.

Muir-Hunter, S., & Wittwer, J. (2016). Dual-task testing to predict falls in community-dwelling older adults: A systematic review. Physiotherapy, 102(1), 29-40.

Mullin, D. S., Cockburn, A., Welstead, M., Luciano, M., Russ, T. C., & Muniz‐Terrera, G. (2022). Mechanisms of motoric cognitive risk—hypotheses based on a systematic review and meta‐analysis of longitudinal cohort studies of older adults. Alzheimer's & Dementia, 18(12), 2413-2427.

Okubo, Y., Schoene, D., Caetano, M. J., Pliner, E. M., Osuka, Y., Toson, B., & Lord, S. R. (2021). Stepping impairment and falls in older adults: A systematic review and meta-analysis of volitional and reactive step tests. Ageing Research Reviews, 66, 101238.

Omaña, H., Bezaire, K., Brady, K., Davies, J., Louwagie, N., Power, S., . . . Hunter, S. W. (2021). Functional reach test, single-leg stance test, and tinetti performance-oriented mobility assessment for the prediction of falls in older adults: A systematic review. Physical Therapy, 101(10), pzab173.

Pesonen, J. S., Vernooij, R. W., Cartwright, R., Aoki, Y., Agarwal, A., Mangera, A., . . . Griebling, T. L. (2020). The impact of nocturia on falls and fractures: A systematic review and meta-analysis. The Journal of Urology, 203(4), 674-683.

Petersen, N., König, H.-H., & Hajek, A. (2020). The link between falls, social isolation and loneliness: A systematic review. Archives of Gerontology and Geriatrics, 88, 104020.

Piirtola, M., & Era, P. (2006). Force platform measurements as predictors of falls among older people–A review. Gerontology, 52(1), 1-16.

Power, V., Van De Ven, P., Nelson, J., & Clifford, A. M. (2014). Predicting falls in community-dwelling older adults: A systematic review of task performance-based assessment tools. Physiotherapy Practice and Research, 35(1), 3-15.

Qian, X. X., Chen, Z., Fong, D. Y., Ho, M., & Chau, P. H. (2022). Post-hospital falls incidence and risk factors among older adults: A systematic review and meta-analysis. Age and Ageing, 51(1), afab209.

Quijoux, F., Vienne-Jumeau, A., Bertin-Hugault, F., Zawieja, P., Lefevre, M., Vidal, P.-P., & Ricard, D. (2020). Center of pressure displacement characteristics differentiate fall risk in older people: A systematic review with meta-analysis. Ageing Research Reviews, 62, 101117.

Ramsey, K. A., Zhou, W., Rojer, A. G., Reijnierse, E. M., & Maier, A. B. (2022). Associations of objectively measured physical activity and sedentary behaviour with fall-related outcomes in older adults: A systematic review. Annals of Physical and Rehabilitation Medicine, 65(2), 101571.

Rosa, M. V., Perracini, M. R., & Ricci, N. A. (2019). Usefulness, assessment and normative data of the Functional Reach Test in older adults: A systematic review and meta-analysis. Archives of Gerontology and Geriatrics, 81, 149-170.

Salonen, L., & Kivelä, S.-L. (2012). Eye diseases and impaired vision as possible risk factors for recurrent falls in the aged: A systematic review. Current Gerontology and Geriatrics research, 2012(1), 271481.

Seppala, L. J., van de Glind, E. M., Daams, J. G., Ploegmakers, K. J., de Vries, M., Wermelink, A. M., . . . Bucht, G. (2018). Fall-risk-increasing drugs: A systematic review and meta-analysis: III. Others. Journal of the American Medical Directors Association, 19(4), 372. e371-372. e378.

Sharma, B., Wang, M., McCreary, C. R., Camicioli, R., & Smith, E. E. (2023). Gait and falls in cerebral small vessel disease: A systematic review and meta-analysis. Age and Ggeing, 52(3), afad011.

Soares, W. J., Lopes, A. D., Nogueira, E., Candido, V., de Moraes, S. A., & Perracini, M. R. (2018). Physical activity level and risk of falling in community-dwelling llder adults: Systematic review and meta-analysis. Journal of Aging and Physical Activity, 27(1), 34-43.

Stubbs, B., Binnekade, T., Eggermont, L., Sepehry, A. A., Patchay, S., & Schofield, P. (2014). Pain and the risk for falls in community-dwelling older adults: Systematic review and meta-analysis. Archives of Physical Medicine and Rehabilitation, 95(1), 175-187. e179.

Stubbs, B., Schofield, P., Binnekade, T., Patchay, S., Sepehry, A., & Eggermont, L. (2014). Pain is associated with recurrent falls in community-dwelling older adults: Evidence from a systematic review and meta-analysis. Pain Medicine, 15(7), 1115-1128.

Trevisan, C., Crippa, A., Ek, S., Welmer, A. K., Sergi, G., Maggi, S., . . . Rizzuto, D. (2019). Nutritional Status, Body Mass Index, and the Risk of Falls in Community-Dwelling Older Adults: A Systematic Review and Meta-Analysis. JAMDA, 20(5), 569-582 e567. doi:10.1016/j.jamda.2018.10.027

Xu, T., Clemson, L., O'Loughlin, K., Lannin, N. A., Dean, C., & Koh, G. (2018). Risk factors for falls in community stroke survivors: A systematic review and meta-Analysis. Archives of Physical Medicine and Rehabilitation, 99(3), 563-573 e565. doi:10.1016/j.apmr.2017.06.032

Yang, Y., Hu, X., Zhang, Q., & Zou, R. (2016). Diabetes mellitus and risk of falls in older adults: a systematic review and meta-analysis. Age and Ageing, 45(6), 761-767. doi:10.1093/ageing/afw140

Zhang, X., Huang, P., Dou, Q., Wang, C., Zhang, W., Yang, Y., . . . Zeng, Y. (2020). Falls among older adults with sarcopenia dwelling in nursing home or community: A meta-analysis. Clinical Nutrition, 39(1), 33-39.
